# Large-scale functional annotation establishes a reference framework for human *LRRK2* variants

**DOI:** 10.64898/2026.06.17.26355034

**Authors:** Anthea Cheung, Neringa Pratuseviciute, Kirsten Black, Pawel Lis, Toan Phung, Melanie Cavin, Gabriel Morel, Anna Saari, Vincent Huin, Simone Zittel-Dirks, Francesca Tonelli, Benjamin Riebenbauer, Thomas Gasser, Javier Ruiz-Martinez, Global Parkinson’s Genetics Program (GP2), Huw R. Morris, Lara M Lange, Allison Dilliott, Orly Goldstein, Shachar Shani, Lionel Arnaud, Alexander Zimprich, Walter Pirker, Christine Klein, Roy Alcalay, Katja Lohmann, Dario R. Alessi, Esther Sammler

## Abstract

Pathogenic variants in leucine-rich repeat kinase 2 (*LRRK2*)^1^are among the most frequent monogenic causes of Parkinson’s disease (PD)^2^ and act through a gain-of-function mechanism of increased kinase activity. *LRRK2*-targeted therapies are in clinical development, but interpretation of the rapidly expanding catalogue of rare *LRRK2* variants remains a barrier to translation. Here, we present functionally annotated data on >350 *LRRK2* coding variants using a standardized cellular assay with Rab10 phosphorylation as a readout of kinase activity and integrated these data with curated genetic and clinical annotations from the Movement Disorders Society Genetic Mutation Database (MDSGene). Variants differed in activation magnitude, ranging from modest increases (e.g., p.G2019S) to strongly activating substitutions such as p.Y1699C or p.L1795F. Activating variants occurred across the full length of LRRK2, although the largest effects clustered within the ROC–COR regulatory hub, where structural analysis identified subdomains forming an allosteric scaffold controlling kinase output. All known/established pathogenic variants showed increased activity, whereas benign and likely benign variants remained within the wild-type range. Functional effect sizes correlated with pathway activation in patient-derived immune cells, altogether providing a framework for ACMG-based variant interpretation in which kinase activation can support PS3 functional evidence for reclassification of variants.

## Introduction

Pathogenic variants in the leucine-rich repeat kinase 2 gene (*LRRK2)* represent one of the most frequent monogenic causes of autosomal dominant Parkinson’s disease. ^2,3–5^. The *LRRK2* gene encodes a large, multidomain protein with a catalytic core comprising a ROC–COR GTPase module and a serine/threonine kinase domain (Figure 1A). ^1,6^ Converging genetic, biochemical, and cellular evidence indicates that these variants drive disease through increased LRRK2 kinase pathway activity, establishing kinase activation as a central pathogenic mechanism. ^7,8^

**Figure 1.**
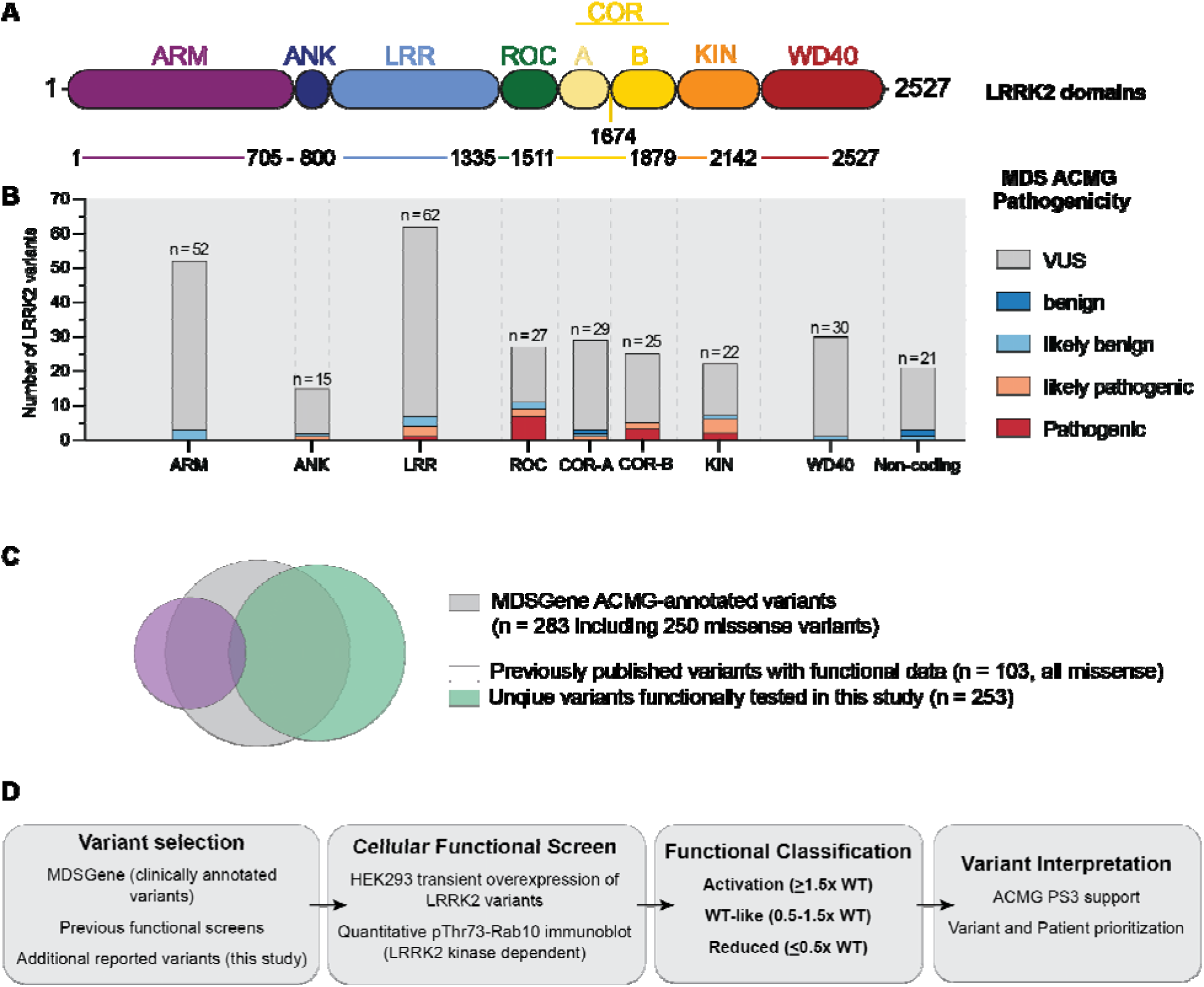
Genetic landscape of reported human *LRRK2* variants and strategy for functional annotation. **(A)** Schematic representation of the multidomain architecture of LRRK2, comprising the armadillo (ARM), ankyrin (ANK), leucine-rich repeat^38^, and atypical Roco family GTPase module consisting of the Ras of complex proteins (ROC) domain with tandem C-terminal of ROC folds (COR-A and COR-B), followed by the catalytic kinase (KIN) domain and the WD40 repeat (tryptophan–aspartic acid repeat) domain, with boundaries as previously defined ^6^. **(B)** Stacked bar plots indicate the number of reported *LRRK2* missense variants per domain, stratified by MDSGene-curated ACMG classification (pathogenic, likely pathogenic, variant of uncertain significance [VUS], likely benign, and benign). Total variant counts per domain are shown above each bar, highlighting that the majority of variants across domains are classified as VUS. Non-missense variants (nonsense, splice-region/splice-site variants, and small deletions) are shown separately as stacked bars to the right. Variants were obtained from genetically annotated *LRRK2* entries curated in MDSGene ^11^. **(C)** Overlap between variant sources and for which functional data is available. Venn diagram illustrating overlap between genetically annotated *LRRK2* variants curated from MDSGene (light grey) ^11^, variants previously assessed in functional screens ^13^ ^15^ ^19^ ^23^)^14^ (purple), and additional variants identified from the literature and other sources (green) that have been included in functional screens in this study. **(D)** Schematic overview of the functional annotation workflow. *LRRK2* variants curated from clinical databases and the literature were assessed using a standardized cellular overexpression assay measuring LRRK2 kinase activity via phosphorylation of endogenous Rab10 at threonine 73 as before ^13^ ^15^ ^14^ ^19^. Functional outcomes were used to classify variants as activating, WT-like, or reduced, providing experimental evidence to support subsequent variant interpretation and prioritization, including application of ACMG PS3/BS3 criteria. PS3 refer to ACMG evidence categories ^16^ indicating strong functional evidence supporting pathogenic variant interpretation.

This mechanistic insight has directly informed the development of LRRK2-targeted therapeutic strategies, including small-molecule kinase inhibitors and LRRK2-lowering approaches, which are now being evaluated in clinical trials. ^9^ These efforts are further supported by the hypothesis that dysregulated LRRK2 kinase pathway activity may also contribute to idiopathic PD, potentially defining a biologically relevant group of PD patients beyond monogenic disease. ^10^

In parallel, next-generation sequencing efforts and large-scale genetic analyses, also in underrepresented populations, have driven a rapid expansion in the number of reported *LRRK2* variants identified in individuals with PD and related phenotypes ^2^ ^11^. However, the vast majority of these variants remain classified as variants of unknown clinical significance (VUS), as highlighted in recent MDSGene analyses. ^11^ Traditional genetic approaches often lack the power to establish pathogenicity because large multigenerational pedigrees are rare, many variants are often identified in only single individuals with uninformative family histories and even established pathogenic *LRRK2* variants such as p.G2019S show incomplete, age-dependent penetrance. ^11,12^ As a result, the key challenge is not the small number of definitively pathogenic *LRRK2* variants per se, but the substantial and growing burden of VUS, underscoring a widening gap between variant discovery and biological or clinical interpretation and translation. ^11^

Here, we address this gap by providing a large-scale functional annotation of human *LRRK2* variants, integrating newly generated and previously published kinase activity data with updated genetic and clinical annotations curated through the Movement Disorders Society Genetic Mutation Database (MDSGene). ^11,13–15^ Using a robust cellular assay that quantifies LRRK2-dependent phosphorylation of Rab10 as a readout of LRRK2’s kinase activity and applying predefined effect-size thresholds for kinase activation established previously, we assessed functional consequences of over 350 *LRRK2* coding variants. Robust kinase activation provides strong functional evidence consistent with the established disease mechanism and can support variant interpretation within current classification frameworks. ^16^ For a subset of strongly activating variants, evidence of LRRK2 pathway activation in clinical samples, including peripheral blood neutrophils and monocytes, offers supportive translational validation of cellular effect sizes, although such evidence is not required for functional interpretation. ^14,15,17–19^ Conversely, the absence of detectable LRRK2 kinase activation does not exclude pathogenicity. Together, this study establishes a reference framework for the functional interpretation of human *LRRK2* variants, intended to complement genetic evidence rather than serve as a definitive classifier alone.

## Materials and methods

### *LRRK2* variant selection

*LRRK2* variants were compiled from multiple sources, including the MDSGene database ^11^, published literature, and personal communications. Full details are provided in the Supplementary materials.

Supplementary Table 1 summarizes all *LRRK2* variants reported in MDSGene ^11^, including variant annotation, ACMG classification, and availability of published functional data. ^13^ Supplementary Table 2 lists all *LRRK2* variants previously evaluated in cellular overexpression assays ^11,13–15,19^, together with functional outcomes and MDSGene annotated ACMG classification where available. Supplementary Table 3 provides a comprehensive overview of *LRRK2* variants with available functional data, including mammalian evolutionary conservation (ConSurf) ^20^ and quantitative measures of kinase activity (mean pRab10/Rab10, SD, and number of replicates) assessed under standardised assay conditions. The dataset combines previously published results ^11,13–15,19^ with newly generated data reported in this manuscript.

### Plasmids

The plasmids were ordered from the MRC PPU Reagents and Services (https://mrcppureagents.dundee.ac.uk). The constructs were all sequenced using whole plasmid sequencing service Plasmidsaurus (https://plasmidsaurus.com/) prior to use. Information about the plasmids used during the duration of this study including their identifiers (DU numbers) can be found summarized in Supplementary Table 4.

### Transformation of competent cells and plasmid purification

2μL of complementary DNA (cDNA) (Supplementary Table 4) was added to competent Escherichia coli (E.coli) DHα - cells and incubated on ice for 20 minutes. Transformation was carried out using the standard heat shock method (42°C, 1 minute) following with 100μL of Luria-Bertani (LB) broth medium without antibiotics was added to each aliquot and incubated for 1 hour at 37°C. After incubation the transformed cell suspension was streaked onto LB agar plates containing 100μg/mL carbenicillin plates were incubated at 30°C for 36-48hrs. Transformed E.coli colonies containing plasmids of interest were amplified by inoculating a single colony into 250mL of LB medium supplemented with 100μg/μL carbenicillin. Cultures were incubated overnight at 30°C with shaking at 180 rpm. After ∼24hrs, bacterial cultures were centrifuged (4000 x g), and the pellets were subjected to plasmid DNA purification using Qiagen Plasmid Maxi Kit according to the manufacturer’s instructions. Purified plasmid concentrations were determined using NanoDrop spectrophotometer.

### Cell culture maintenance

The human embryonic kidney 293 (HEK293) cells, obtained from ATCC, were cultured in Dulbecco’s Modified Eagle Medium (DMEM) supplemented with 10% (v/v) foetal calf serum (FBS), 2 mM L-glutamine, 100 U/mL penicillin-streptomycin, at 37°C in a humidified incubator containing 5% CO_2_.

### Transient cell transfection, cell treatments and lysis

For transient transfections, HEK293 cells were seeded in 6-well plates at a density of 0.5-0.6 x 10^6^ cells per well. The following day, at ∼70-80% confluency the cells were transfected with 2μg total cDNA using polyethylenimine (PEI) as the transfection reagent at a DNA : PEI ratio of 1:3, in a total volume of 300μL Opti-MEMTM Reduced Serum Medium. Prior to being added to the cells the DNA : PEI mixtures were left at room temperature for 20min to allow proper complex formation. Once transfected, cells were incubated for 16-20 hours prior to cell treatments and lysis. Where specified cells were treated with the selective LRRK2 kinase inhibitor, MLi-2, synthesized by Natalia Shpiro (University of Dundee), at a final concentration of 200nM for 90min. Treated and untreated cells were washed and lysed with ice cold 1% Triton X-100 lysis buffer containing 50 mM Tris/HCl (pH 7.4), 1% (v/v) Triton X-100, 1 mM EGTA, 1mM sodium orthovanadate, 50mM sodium fluoride, 10mM 2-glycerophosphate, 5mM sodium pyrophosphate, 270 mM sucrose, 0.1% (v/v) 2-mercapthoethanol, complete EDTA-free protease inhibitor cocktail and freshly supplemented with 1μg/mL microcystin-LR. Post-lysis the cell debris was cleared by centrifuging the samples at 17 000 x g for 15min at 4°C.

### Quantitative immunoblotting

Total protein concentration of all cell lysates was determined using PierceTM BCA Protein Assay Kit, according to the manufacturer’s instructions. Cell lysates were prepared in 4x NuPage LDS Sample Buffer supplemented with 5% β-mercaptoethanol and boiled at 96°C, 10 minutes. Equal amounts of protein (10-20µg per lane) were loaded on NuPAGE 4-12% Bis-Tris gradient gels alongside a molecular weight protein ladder. Electrophoresis was carried out in 1x MOPS SDS running buffer at 90V until samples entered the resolving gel and then at 120V. Proteins were transferred to a 0.45μm nitrocellulose membrane at 90V for 90 minutes in 1x transfer buffer (48mM Tris-HCl, 39mM glycine). Once transferred, the membranes were briefly stained with Ponceau S dye and cut accordingly to molecular weight of the target proteins of interest to visualize. Membranes were washed in TBS-T (20mM Tris-HCl, 150mM NaCl, 0.2% Tween20) to remove residual stain and left to block for 30 minutes at room temperature in 5% non-fat dry milk diluted in TBS-T. The membranes were washed with TBS-T before incubation with appropriate primary antibodies, all prepared in TBS-T containing 5% BSA and 0.02% NaN_3_. The following primary antibodies were used: pRab10-Thr73 (Abcam, #ab241060, 1:1000) multiplexed with total Rab10 (Nanotools (Biomol), #0680-100, 1:500), pSer935-LRRK2 (Abcam, #ab13345, 1:1000) multiplexed with total LRRK2 (C-terminal) (Antibodies inc. /NeuroMab, #75-253, 1:1000) and GAPDH (Santa Cruz Biotechnology, #sc-32233, 1:10 000). The membranes were left to incubate overnight at 4°C. The next day, after wash in TBS-T, the membranes were incubated in the dark (1 hour at room temperature) with multiplexed Goat Anti-Mouse IRDye 680RD (LICOR, #926-68070, 1:10 000) and Goat Anti-Rabbit IRDye 800CW (LICOR, #926-32211, 1:10 000) fluorescent secondary antibodies (in 5% milk, TBS-T). After incubation the membranes were washed in TBS-T and the signals were visualized using a LI-COR Odyssey CLx imaging system and the bands were quantified using Image Studio Lite.

### Patient recruitment and blood sample collection

Patients with Parkinson’s disease and healthy individuals were recruited from multiple clinical sites across Europe (Supplementary Table 5). For each participant relevant demographic and clinical information was collected including age, age at onset and family history of PD where applicable. Blood samples were collected at their respective recruitment sites, with 20-40mL blood collected per participant into EDTA-coated BD Vacutainer tubes by venepuncture. Samples were processed within 2 hours of collection to ensure best sample quality. All participants provided informed written consent prior to enrolment in accordance with Declaration of Helsinki and approved by the appropriate institutional ethics committees.

### Isolation and processing of human neutrophils and monocytes

Neutrophil isolations were performed via immunomagnetic negative selection following previously described method. ^18^ Briefly, fresh blood was transferred into 50mL falcon tubes and EasySepTM direct Human Neutrophil Isolation Cocktail and EasySepTM Direct RapidSpheres magnetic beads were added (50 μL/mL of sample). Blood was subsequently mixed by gentle inversion and incubated for 5 minutes at room temperature. Following incubation, the tubes were topped up to 50mL with Phosphate-buffered saline (PBS) containing 1mM EDTA, mixed again by inversion and placed into the Easy 50 EasySep™ Magnet for 10 minutes. After transferring neutrophil containing cell suspension in to fresh tubes, RapidSpheres beads were added again followed by 5min incubation at RT. Tubes were then returned to the magnet for an additional 5 minutes, after which the cell suspensions were transferred into new 50mL falcon tubes. To ensure complete removal of magnetic beads the tubes were placed back into the magnet for additional 10 minutes. The resulting cell suspensions were transferred into fresh 50mL falcon tubes and centrifuged at 450 x g for 5 minutes. The supernatant was carefully discarded. For samples undergoing treatments, the neutrophil pellets were resuspended in Roswell Park Memorial Institute (RPMI) 1640 media and split equally. Cells were then treated with either vehicle control (DMSO) or LRRK2 kinase inhibitor MLi-2 (200nM) for 30 minutes. Finally, cells were centrifuged at 450 x g for 5 minutes, and the supernatant was carefully discarded and pellets lysed using ice cold Triton X-100 lysis buffer containing 50 mM Tris/HCl (pH 7.4), 1% (v/v) Triton X-100, 1 mM EGTA, 1mM sodium orthovanadate, 50mM sodium fluoride, 10mM 2-glycerophosphate, 5mM sodium pyrophosphate, 270 mM sucrose, 0.1% (v/v) 2-mercapthoethanol, complete EDTA-free protease inhibitor cocktail and freshly supplemented with 1μg/mL microcystin-LR and 0.5mM diisopropylfluorophosphate (DIFP). To remove cell debris the lysates were centrifuged at 17 000 x g for 15 minutes at 4°C.

### Structural modelling

Structural similarities of the four newly proposed COR subdomains were identified using structural alignments performed with the DALI server (http://ekhidna2.biocenter.helsinki.fi/dali/). DALI searches were carried out against both experimentally determined structures deposited in the Protein Data Bank and predicted structures available in the AlphaFold2 database, and the resulting matches were used to define structurally related folds and guide the comparative analysis. PyMOL 3 was used to visualize the structures. Key structures and models discussed here include LRRK2/COR-region views based on PDB 8VH4, the 14-3-3γ-LRRK2 COR interface (PDB 9CI3), the Rab29-LRRK2 cryo-EM structure (PDB 8FO2), LRRK2 dimeric structure (PDB 8U8A), human LRRK1 (PDB 8E04), and the bacterial Chlorobaculum tepidum Roco LRR-Roc-COR structure (PDB 6HLU). Additional comparative structures used for subdomain analysis include RAPGEF3 (PDB 6H7E), DEPTOR (PDB 7DKL), PREX1 (PDB 7RX9), MCU (PDB 5Z2H), Pseudomonas aeruginosa phosphomannomutase (PDB 2FKM), Thermococcus kodakarensis phosphomannomutase/phosphoglucomutase-like protein (PDB 9IX8), Thermotoga maritima S-adenosylmethionine decarboxylase (PDB 1TMI), CHIP U-box (PDB 9DRY), and TRIM2 RING (PDB 7ZJ3). In selected cases, AlphaFold3 (https://alphafoldserver.com) models were additionally used to complement the available cryo-EM structures, which do not fully resolve all regions of the LRRK2 COR domain and therefore lack some local structural details.

### Population frequency analysis

To assess the population occurrence of functionally characterized *LRRK2* variants, allele frequencies were extracted from the Global Parkinson’s Genetics Program (GP2) ^21^ Genome Browser (based on data from Release 10; browser assessed in April 2026; https://gp2.broadinstitute.org/) ^22^, using the European (non-Finnish) dataset. For each variant, allele count (AC), allele number (AN), and allele frequency (AF) were retrieved for Parkinson’s disease ^2^ patients and healthy controls from short-read whole-genome sequencing (WGS). Notably, there is some minor overlap of samples between GP2 and PD GENEration, as a small subset of samples from PD GENEration also underwent short-read WGS through GP2.

PDGENEration was queried for variation at 321 LRRK2 codons corresponding to the 364 amino acid substitutions included in the functional dataset. Genetic data were generated by clinical exome sequencing or whole-genome sequencing at Fulgent Genetics, a CLIA-certified laboratory, as previously described. ^5^ Variant calls were extracted from sample-level VCF files using genomic coordinates corresponding to the 963 interrogated bases (hg19/GRCh37). All variants passed quality control filters applied by Fulgent Genetics but were not independently validated by secondary sequencing.

## Data availability

The authors confirm that the data supporting the findings of this study are available within the article, its supplementary material, and in an openly accessible Zenodo repository [DOI: 10.5281/zenodo.19855030]

Data from the GP2 Genome Browser can be accessed at https://gp2.broadinstitute.org/. Access to the full GP2 dataset can be requested through AMP PDRD (https://amp-pdrd.org), where also the PDGENEration can be accessed.

## Results

### Genetic landscape of reported human *LRRK2* variants

To define the current genetic landscape of human *LRRK2* variation in PD, we used variants curated through MDSGene and mapped them across *LRRK2* domains and current pathogenicity categories according to MDS curated American College of Medical Genetics classification (Figure 1B). ^11,16^ In this dataset, 283 unique *LRRK2* variants were found across 4660 *LRRK2* variant carriers, including 197 unique *LRRK2* in 3387 patients with PD. Of these, only 13 variants were classified as pathogenic and 12 as likely pathogenic, whereas the majority, 241 variants were interpreted as VUS (Figure 1B).

Missense variants constitute the predominant class of reported variation (n = 250) and are distributed across all domains (Figure 1B). In contrast, other variants - including truncating or nonsense (n = 7), synonymous (n = 5), splice-region, or small deletion variants (n = 21) - are comparatively rare (Figure 1B). Across all domains, the majority of reported variants are classified as VUS (n = 241, including 215 missense variants) (Figure 1B). This distribution extends across the full length of the protein; however, pathogenic and likely pathogenic variants cluster predominantly within the ROC, COR-B, and kinase domains and are exclusively missense. A detailed overview of the *LRRK2* structure and variant distribution is provided Figure 1, with interactive versions available via links in the figure legends to facilitate exploration of individual variants.

To assess the extent of available functional evidence for reported variants, we compared variants curated in MDSGene ^11^ with those previously evaluated in cellular functional assays ^13–15,19,23^ as well as with additional variants identified through the literature and other sources (Figure 1C). Functional data exist for a subset of reported variants (n = 103) (Supplementary Tables 1 and 2), with partial overlap with MDSGene-curated variants (n = 283, including 250 missense variants of which 184 variants were found in PD patients). However, a substantial proportion of *LRRK2* variants reported in PD patients and in individuals without PD or other reported conditions has not been evaluated using standardized functional assays (n=210, including 177 missense variants of which 121 are reported in PD patients) (Figure 1C).

### Functional *LRRK2* kinase activity across variants

To systematically assess the functional consequences of human *LRRK2* variation, we established a standardized workflow in which variants are evaluated using a cellular overexpression assay quantifying *LRRK2* kinase–dependent phosphorylation of Rab10 at threonine 73 (pThr73-Rab10) (Figure 1D). To ensure consistency and comparability with prior studies ^13–15,19^, we applied a predefined threshold of ≥1.5-fold increased Rab10 phosphorylation relative to wild-type *LRRK2* as an operational criterion to classify variants as kinase-activating.

Using this approach, kinase activity across 356 variants (Supplementary Table 3), including 103 variants previously characterised under comparable assay conditions, with the remainder representing newly generated data incorporated within the same standardised framework. Variants were ranked according to the mean fold-change in pThr73-Rab10 phosphorylation relative to wild-type *LRRK2* and visualized as a continuous distribution (Figure 2A).

**Figure 2.**
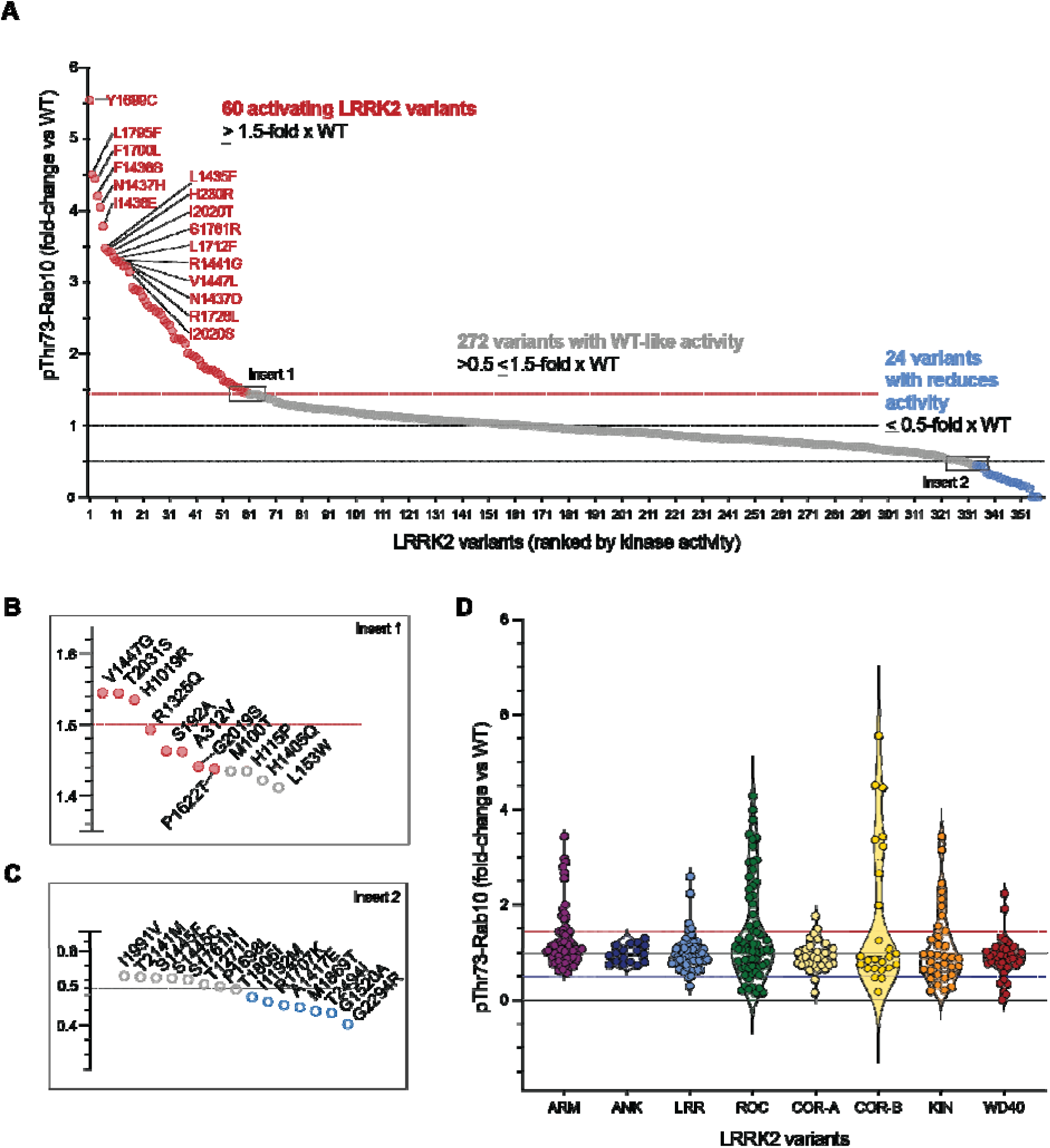
Functional landscape of *LRRK2* kinase activity across human variants. **(A)** Ranked distribution of LRRK2 kinase activity across 359 *LRRK2* variants with available functional data, comprising 354 single missense variants and three nonsense variants. All variants were assessed in a robust HEK293 cellular overexpression assay with at least three independent biological replicates, and data are shown as the mean fold-change in LRRK2 kinase–dependent pThr73-Rab10 phosphorylation relative to wild-type *LRRK2*. Variants are ordered by decreasing kinase activity. Based on these criteria, 60 variants were classified as activating (red), 272 variants as WT-like (grey), and 24 variants as reduced activity (blue). Double variants are plotted separately at the far right of the x-axis. **(B)** Zoomed inset of panel (A) highlighting variants near the activation threshold (1.5-fold), illustrating the distribution of mildly activating variants at the upper boundary of WT-like activity. **(C)** Zoomed inset of panel (A) highlighting variants near the reduced activity threshold (0.5-fold), illustrating variants with reduced kinase activity at the lower boundary of WT-like activity. **(D)** Distribution of LRRK2 kinase activity across protein domains. Variants are grouped by LRRK2 domain (ARM, ANK, LRR, ROC, COR-A, COR-B, kinase, and WD40), and pThr73-Rab10 phosphorylation values are displayed as violin plots, summarizing the range and distribution of functional effects within each domain. This representation reveals domain-specific enrichment and variability of activating and reduced-activity variants within the overall functional landscape.

Kinase activity spanned a broad dynamic range, from strongly increased activity (>5-fold) to marked reduction relative to wild-type *LRRK2* (<0.2-fold for missense and 0 for non-sense variants). Applying the predefined classification criteria, 60 single missense variants exhibited increased kinase activity (≥1.5-fold relative to wild-type), 272 variants showed activity within the wild-type range (>0.5–<1.5-fold), and 24 variants including 3 non-sense variants, demonstrated reduced kinase activity (≤0.5-fold) (Figure 2A and C).

The ranked distribution revealed a gradual transition between wild-type–like and activating, as well as between wild-type like variants and reduced-activity variants (Figure 2A-C).

When stratified by protein domain, functional effects were observed across all *LRRK2* domains (Figure 2D). Variants with increased kinase activity were predominantly located within the enzymatic core, comprising the ROC, COR-B, and kinase domains, but were also identified in the ARM, LRR, and WD40 domains. To date, no activating variants have been observed in the ANK domain. Variants exhibiting reduced kinase activity clustered within the enzymatic core (ROC, COR-B, and kinase domains) as well as the WD40 domain, while variants with wild-type-like activity were distributed throughout the protein. Violin plots further illustrate domain-specific variability in both the magnitude and distribution of kinase activity changes (Figure 2D).

### Functional effects by pathogenicity category

To assess the relationship between functional effects and existing genetic classification, *LRRK2* kinase activity was stratified according to MDSGene-curated ACMG categories (Fig 3A). ^11^ Variants classified as pathogenic consistently exhibited increased kinase activity. Most exceeded the predefined activation threshold, with the exception of p.G2019S, which showed a modest but reproducible increase in activity (1.44-fold; SD 0.45) just below the threshold. In contrast, benign and likely benign variants displayed kinase activity within or below the wild-type range and did not surpass the activation threshold.

**Figure 3.**
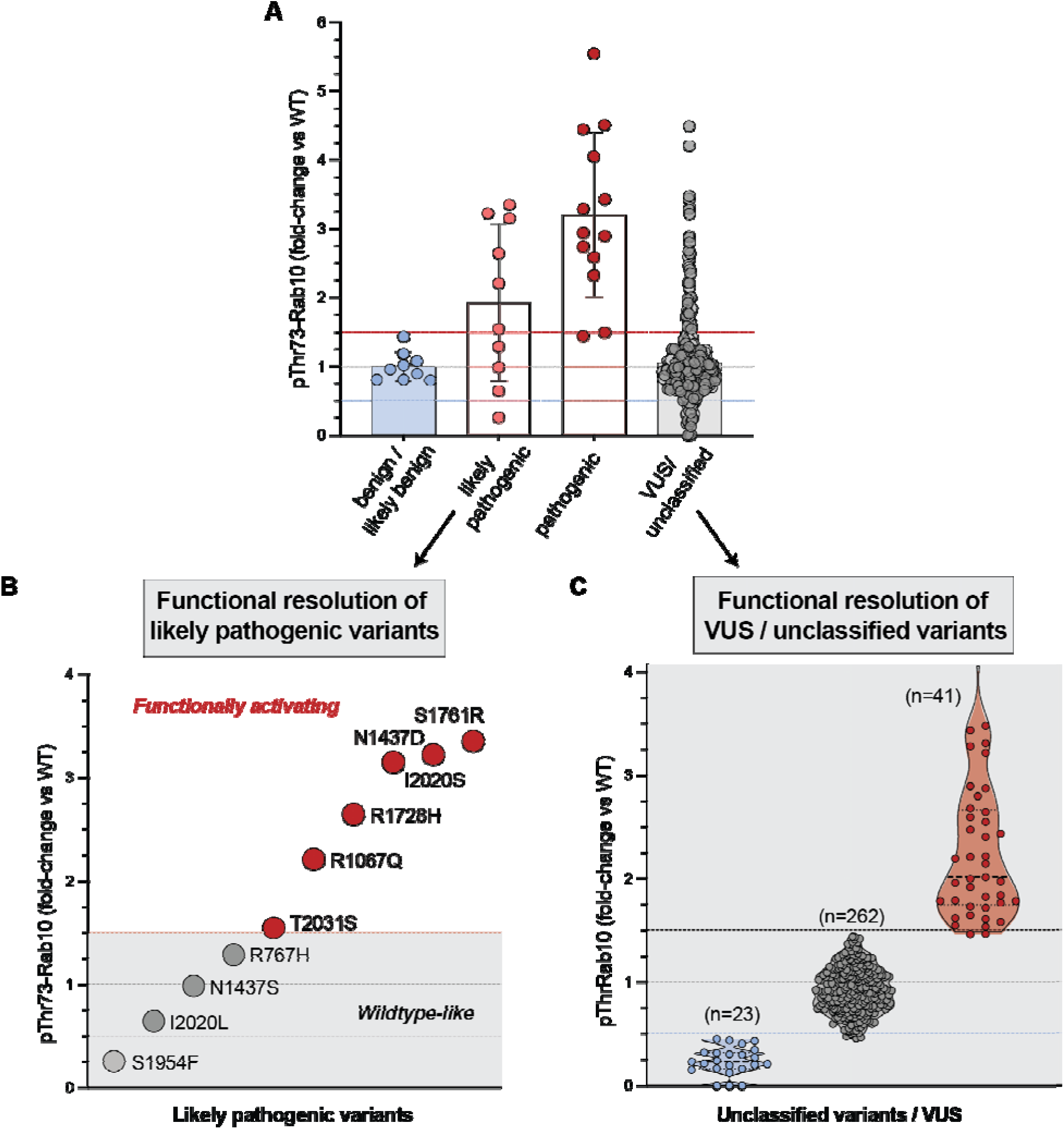
Functional effects of *LRRK2* variants in the context of genetic classification. **(A)** LRRK2 kinase activity, measured as LRRK2-dependent pThr73-Rab10 phosphorylation, stratified by MDSGene-curated ACMG classification ^11^. Additional unclassified variants (not curated by MDSGene) that were functionally evaluated in this study are included within the VUS category. Each point represents the mean fold-change relative to wild-type (WT) *LRRK2* from a standardized HEK293 overexpression assay, with bars indicating group means ± SD. Horizontal reference lines denote WT activity (1.0) and thresholds for activating (>1.5-fold) and reduced (<0.5-fold) kinase activity. Benign and likely benign variants display WT-like or reduced activity, whereas pathogenic variants uniformly show increased kinase activity. In contrast, likely pathogenic and VUS/unclassified variants exhibit heterogeneous functional effects, consistent with the dynamic nature of pathogenicity classification as additional functional, segregation, and population data emerge. **(B)** Variant-level functional resolution of likely pathogenic *LRRK2* variants. Individual variants are ordered by increasing kinase activity and segregate into functionally activating and WT-like/reduced groups, highlighting heterogeneity within this classification category. **(C)** Functional stratification of variants lacking definitive pathogenicity classification. VUS ^11^ and unclassified variants are grouped by functional outcome and shown as distributions of pThr73-Rab10 phosphorylation. Violin plots illustrate the density and spread of reduced, WT-like, and activating activities with individual data points overlaid. A subset of VUS and unclassified variants exhibits clear LRRK2 kinase activation, whereas most display WT-like or reduced activity.

Variants classified as likely pathogenic showed greater heterogeneity, with kinase activity values distributed both within the wild-type range (0.26-3.35-fold) and above the activation threshold (Supplementary Table 6). Similarly, variants categorized as VUS or unclassified exhibited a broad distribution of kinase activity, spanning reduced, wild-type–like, and activating properties (0-4.49-fold) (Fig 3A, Supplementary Table 6).

To enable detailed comparison at the variant level, MDSGene-curated pathogenicity classifications were integrated with available quantitative functional measurements and additional annotation parameters (Supplementary Table 6). For each variant, concordance between genetic classification and observed functional group is indicated. Notably, concordance was complete for pathogenic (except p.G2019S which was borderline) and benign / likely benign variants under the predefined criteria, whereas discordance was otherwise confined to variants within the likely pathogenic category (4 variants with activity <1.5-fold and 6 with increased activity ≥1.5).

To further assess resolution within individual categories, VUS were stratified by kinase activity (Fig 3B, C). Within this group, 41 variants exceeded the activation threshold, 262 displayed wild-type-like activity, and 23 demonstrated reduced kinase activity. Under current ACMG frameworks, robust functional evidence consistent with the established disease mechanism (PS3) provides support for pathogenicity classification. Accordingly, the 41 activating VUS represent candidates for reclassification as likely pathogenic, substantially increasing the number of variants with supportive functional evidence.

### *LRRK2* variant frequencies in PDGENEration and GP2

To place the functionally characterized *LRRK2* variants in a population context, we examined their occurrence in two large Parkinson’s disease sequencing resources, PDGENEration (n = 29,213 cases) ^5^ and GP2 (n = 13,063 cases and n = 9912 controls) ^21,24^ (Table S7). Among the amino acid substitutions included in the functional dataset, 134 were observed in PDGENEration and 100 in GP2 cases, with substantial overlap including 85 substitutions detected in both resources. Many variants were observed only rarely, including several of the 41 LRRK2 kinase-activating VUS. In some instances, additional genetic and clinical evidence was available. For example, one highly activating variant, LRRK2 p.F1436S (4.21-fold; SD 1.81) was identified in a multigenerational autosomal dominant PD pedigree comprising six affected individuals across three generations. Together, these data illustrate how population-based datasets, family studies, clinical observations and functional characterization provide complementary evidence for the interpretation of rare LRRK2 variants.

### Activating *LRRK2* variants overview

To enable direct comparison of activation magnitude across studies and domains, all *LRRK2* variants previously identified as kinase-activating were re-expressed and analysed in parallel under standardized experimental conditions in three independent biological replicates (Fig. 4A-B, Supplementary Fig 1, 5). This series comprised 25 variants previously reported as activating ^13–15,19^, together with additional activating variants identified in the present screen. Normalizing pThr73-Rab10 phosphorylation to total *LRRK2* protein levels enabled calibrated comparison of effect sizes while controlling for differences in expression levels and batch effects (Fig. 4B).

**Figure 4.**
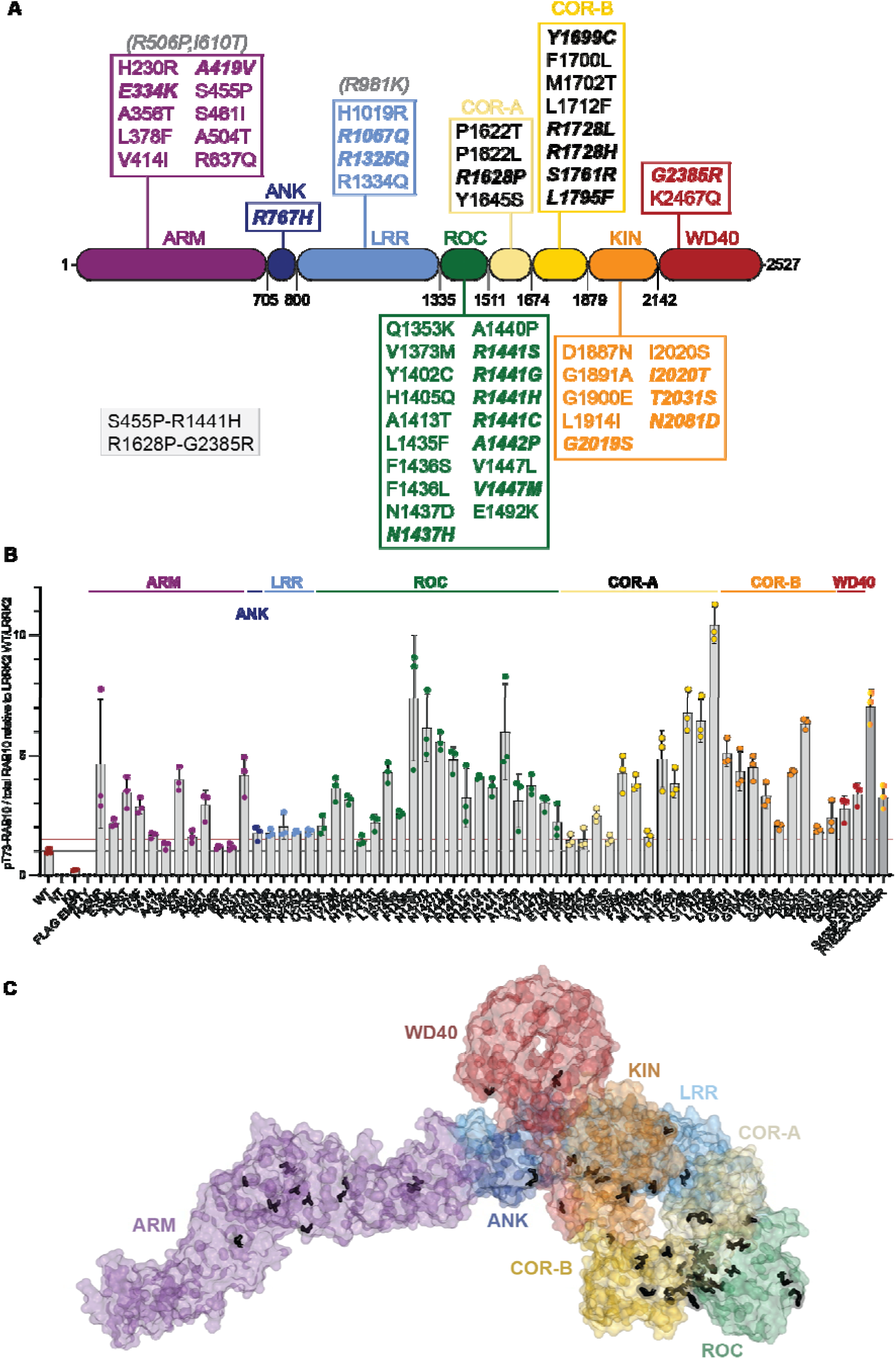
Overview of validated *LRRK2* kinase–activating variants. **(A)** Schematic representation of the LRRK2 protein domain architecture (ARM, ANK, LRR, ROC, COR-A, COR-B, kinase, and WD40) indicating the location of all confirmed *LRRK2* kinase–activating variants identified across independent cellular functional screens. Domain boundaries are shown for orientation. **(B)** Quantitative assessment of LRRK2 kinase activity for all variants previously reported or identified as kinase-activating. Activation was defined as a ≥1.5-fold increase in LRRK2-dependent pThr73-Rab10 phosphorylation relative to wild-type *LRRK2*. All candidate activating variants were re-expressed and analysed in parallel under standardized HEK293 overexpression conditions in three independent biological replicates, alongside wild-type *LRRK2*, kinase-dead *LRRK2*, and non-transfected controls. Bars represent the mean fold-change in pThr73-Rab10 phosphorylation relative to wild-type *LRRK2*, normalized to total LRRK2 protein levels; individual data points indicate biological replicates. Variants are arranged according to ascending amino acid position within LRRK2, with double variants displayed separately at the right end of the axis. Of the 25 previously reported activating variants, all but one exceeded the predefined activation threshold in this standardized validation experiment, demonstrating strong reproducibility of kinase activation across independent studies. **(C)** Structural representation of full-length *LRRK2* (PDB 8VH4) with activating variants shown as black sticks. Domains are coloured and labelled throughout the model. This panel provides an overview of the spatial distribution of activating variants across the full-length protein and highlights the prominent concentration of variants within the ROC-COR region relative to other parts of *LRRK2*.

Under these conditions, all but one (p.A419V) of the 25 previously reported activating variants exceeded the predefined activation threshold (Fig. 4B, Supplementary Fig 1, Supplementary Table 5). *LRRK2* p.A419V showed a modest but reproducible increase in Rab10 phosphorylation (1.2-fold; SD 0.16), placing it below the activation cutoff within the wildtype-like range. *LRRK2* p.R981K, previously suggested to be, did not reproduce increased kinase activity in validation experiments ^13^ or in our analysis and was therefore not included in this final screen (Fig. 4A). Similarly, *LRRK2* p.I610T and p.A506P, which had shown mild activation in earlier experiments, did not exceed the activation threshold when reassessed under standardised conditions (Fig. 4A, B, Supplementary Table 2,3).

Direct comparison revealed substantial differences in activation magnitudes. Variants within the enzymatic core - particularly the ROC, COR-B and kinase domains - generally exhibited larger effect sizes than variants in non-enzymatic regions (Fig. 4B, Supplementary Table 3). Mapping onto the cryo-EM structure showed that variants with the strongest activating effects cluster within the ROC–COR region, identifying this interface as a key regulatory hotspot linking GTPase and kinase activity.

All constructs produced detectable total *LRRK2* protein levels (Supplementary Fig. 1, 2A and Supplementary Table 5), and normalization to total protein confirmed that observed differences in Rab10 phosphorylation were not attributable to altered protein expression. Consistent with previous studies, several activating *LRRK2* variants were associated with reduced phosphorylation of *LRRK2* p.Ser935 ^1^ (Supplementary Fig. 1, 2B), a modification linked to a conformational shift toward the active kinase state and reduced 14-3-3 binding. ^25^ In line with this model, several ROC domain variants displayed reduced Ser935 phosphorylation (Supplementary Fig. 1, 2B), supporting a link between ROC–COR conformational destabilization and increased kinase activation.

Together, these data establish a unified, internally calibrated framework for quantifying *LRRK2* kinase activation and enable direct comparison of variant effect sizes across structural domains.

### ROC–COR interface as regulatory hub for LRRK2 activation

Mapping of activating variants onto the cryo-EM structure of full-length *LRRK2* revealed that substitutions with the largest activating effects localise predominantly to the ROC-Cor region (Fig. 5A), which forms the interface between the ROC GTPase and kinase domains and is therefore well positioned to regulate communication between these enzymatic centres.

**Figure 5.**
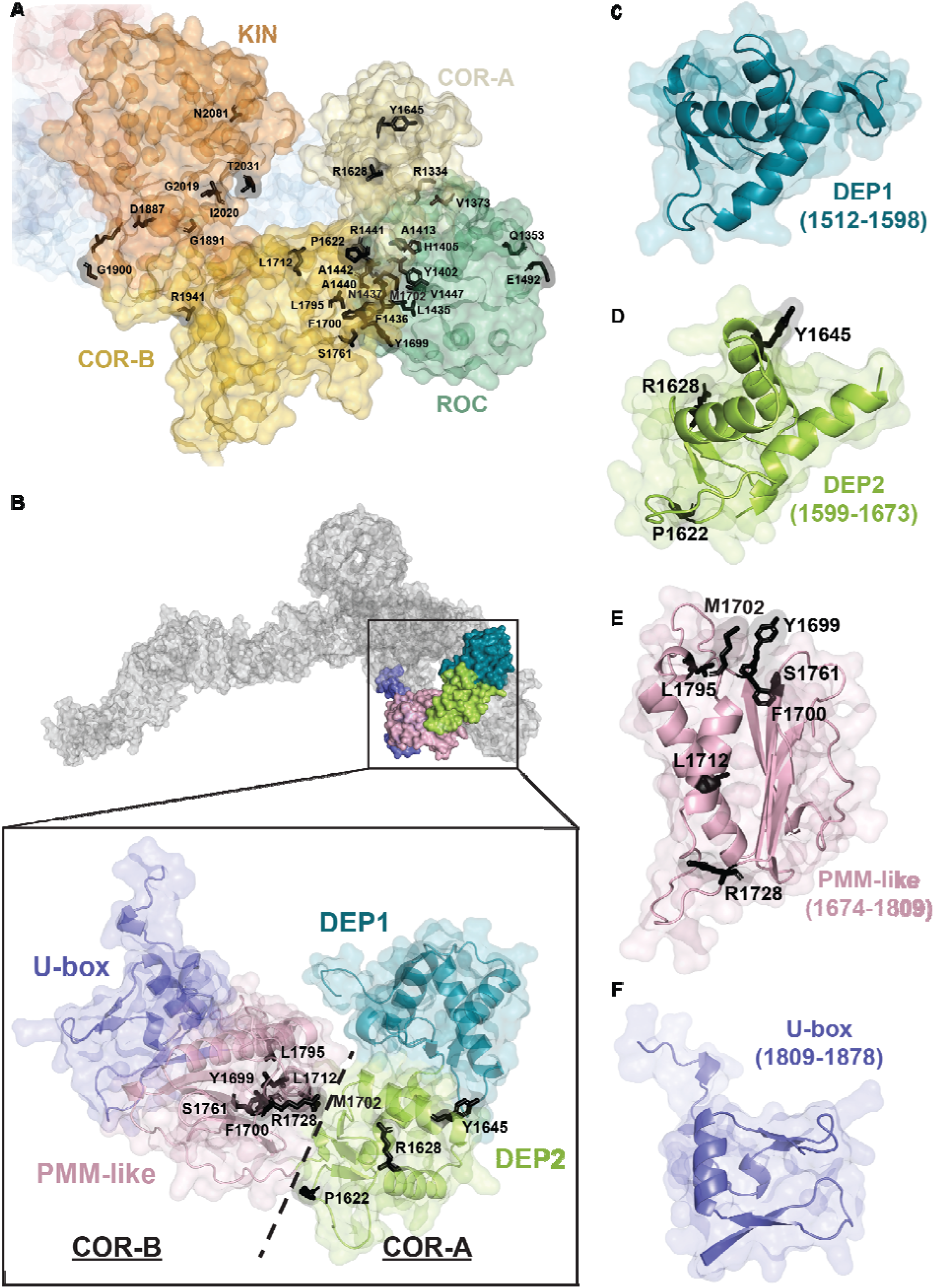
Structural analysis of activating variants in the ROC-COR region and redefinition of the COR scaffold. **(A)** Focused view of the ROC-COR-kinase region of LRRK2 (PDB 8VH4). Activating variants are shown as black sticks and labelled by residue number. This panel highlights the clustering of activating substitutions within the ROC/COR region and at interfaces linking ROC, COR and kinase. **(B)** Isolated view of the COR region in the conventional COR-A/COR-B framework, with activating variants shown as black sticks. This panel illustrates the distribution of activating variants within the traditionally defined COR architecture. **(C)** Isolated COR region subdivided into four proposed subdomains, termed DEP1, DEP2, PMM-like and U-box-like. This panel presents a more in depth structural framework proposed here for interpreting the COR region as a composite regulatory scaffold rather than a simple COR-A/COR-B unit (AlphaFold3 model was used to complement the available cryo-EM structure, which does not fully resolve all regions of the LRRK2 COR domain and therefore lack some local structural details). **(D)** Isolated view of the DEP1 subdomain from panel C. This panel highlights the position and structural boundaries of the first proposed COR subdomain within the revised four-part framework. **(E)** Isolated view of the DEP2 subdomain from panel C. This panel highlights the position and structural boundaries of the second proposed COR subdomain and provides the structural context for variants mapping to this region. **(F)** Isolated view of the PMM-like subdomain from panel C. This panel highlights the position and structural boundaries of the mutation-enriched PMM-like region, which contains the largest number of patient-derived activating variants among the four proposed COR subdomains. **(G)** Isolated view of the U-box-like subdomain from panel C. This panel highlights the position and structural boundaries of the fourth proposed COR subdomain within the revised COR architecture.

The COR scaffold is evolutionarily conserved across ROCO GTPase-containing proteins and is present throughout diverse taxa including animals, bacteria and plants. ^26^ Structural studies indicate that the COR-B domain contributes directly to *LRRK2* dimerization ^6^, while recent cryo-EM analysis of the *LRRK2*–14-3-3 complex demonstrates additional interaction surfaces spanning both COR-A and COR-B, distinct from the principal phospho-dependent docking sites. ^27^ Together, these observations support a model in which the ROC–COR region forms a central regulatory platform governing conformational transitions linked to kinase activation.

#### Structural Subdivision of COR scaffold reveals mutation-enriched modules

To define the structural basis of variant clustering within the COR region, we performed comparative structural analyses using DALI-based alignments together with AlphaFold models. These analyses support a revised view in which the COR region comprises four structurally distinct subdomains forming a composite regulatory scaffold (Fig. 5C) conserved across diverse taxa (Fig 5, figure supplement 1).

Within COR-A, two tandem globular modules display strong structural similarity to DEP (Dishevelled, Egl-10, Pleckstrin) domains, a structure commonly found in membrane-interacting signalling proteins. ^28^ Structural comparisons with signalling regulators, including RAPGEF3, PREX1, PREX2 and DEPTOR that contain such a module (Fig. 5, figure supplement 2A) supports designation of these regions as DEP1 and DEP2 (Fig. 5C–E). Activating variants are absent from DEP1 but cluster within DEP2, including at residues P1622/ R1628/ Y1645. Notably, the Y1645 surface overlaps with a secondary 14-3-3 interaction interface (Fig. 5, figure supplement 1B), suggesting that disruption of DEP2-mediated contacts may promote kinase activation.

In models of Rab29-mediated membrane recruitment ^29^, the DEP2 surface faces the membrane and contains clusters of positively charged residues consistent with potential phospholipid interaction ^30^, whereas DEP1 shows similarity to the Dictyostelium mitochondrial calcium transporter MCU ^31^ (Fig 5, figure supplement 2C-F) raising the possibility that membrane association or metal-dependent conformational changes influence COR-mediated coupling between ROC and kinase domains.

The COR-B region contains a conserved module with similarity to regulatory domains in prokaryotic phosphomannomutase ^32^, as well as human S-adenosylmethionine decarboxylase and Spermine synthase that we termed here PMM-like subdomain (Fig 5, figure supplement 3A). Structural similarity of the PMM-like subdomain to several metabolic enzyme folds raises the possibility that it may function as a small-molecule-responsive regulatory element capable of influencing local conformation. This region harbours the greatest density of activating substitutions (at residues Y1699, F1700, M1702, L1712, R1728, S1761 and L1795, Fig 5F), consistent with a major role in controlling the activation state of LRRK2. R1728 lies close to the dimer interface and near to the secondary 14-3-3 binding surfaces, suggesting that altered dimer stability or reduced 14-3-3 association may favour adoption of an active kinase conformation (Fig. 5, figure supplement 1B).

The PMM-like module lies adjacent to ROC switch-II, with activating residues Y1402 and H1405 located within the switch-II helix, and M1702 contacting this element, providing structural explanation by which local perturbations within the COR scaffold could influence ROC conformational dynamics and thereby modulate kinase output.

A fourth compact β-sheet–rich module at the distal end of COR-B exhibits structural similarity to U-box domains found in E3 ubiquitin ligases such as CHIP and TRIM2 (Fig. 5C, G, figure supplement 3B). Although no intrinsic ubiquitin ligase activity has been demonstrated for LRRK2, conservation of this scaffold across ROCO proteins suggests an evolutionarily retained regulatory interaction surface. Notably, activating variants were not observed within this region, further underscoring the selective enrichment of variants within specific COR subdomains.

### Clinical correlation of LRRK2 kinase activation

We previously demonstrated that LRRK2 kinase pathway activity can be assessed in peripheral blood immune cells, including neutrophils and monocytes, and validated this approach using selective LRRK2 inhibition to confirm pathway specificity. ^17,18,33^ Building on this framework, we assembled clinical biosamples from carriers of multiple rare *LRRK2* variants and analysed them under matched experimental conditions. The series included carriers of variants not previously examined for LRRK2 kinase pathway activity (p.R506P, p.R1325Q, p.I2020T, p.A1440P), together with previously reported carriers of rare variants (p.V1447L, p.F1700L, and p.Y1699C) ^15,19,34^ as well as a carrier of p.R1441C, collectively spanning 11 distinct variants (Fig. 6A–C, Supplementary Fig. 3, Supplementary Table 5). Integrating newly generated and published samples enabled direct comparison of multiple rare *LRRK2* variants side by side in patient-derived material. The variants spanned a broad range of effect sizes (up to 6-fold) in the standardized HEK293 assay (Supplementary Fig. 4). For each carrier, at least one matched control sample collected and processed at the same time. Clinical information for all participants is provided in Supplementary Table 5.

**Figure 6.**
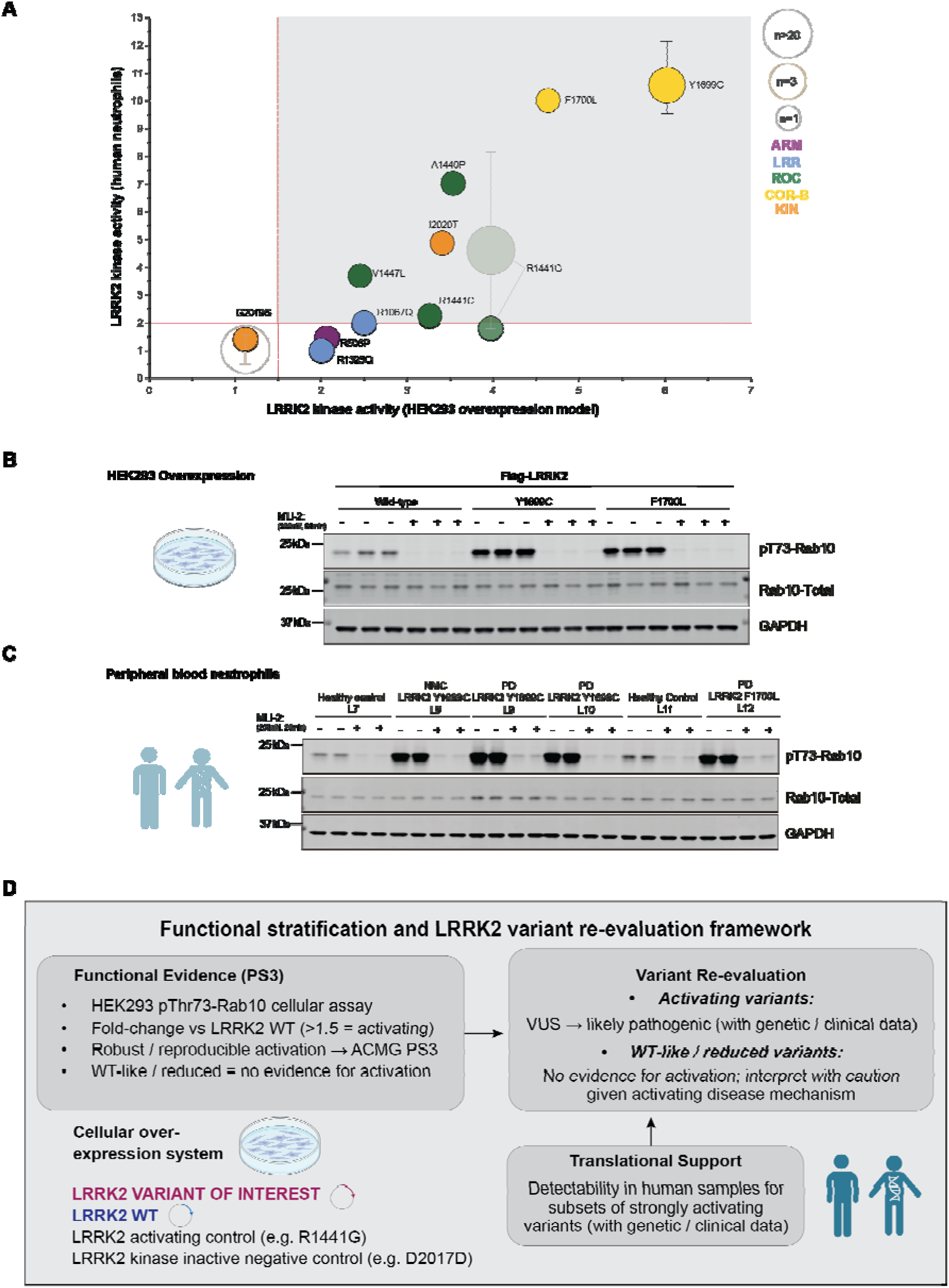
Clinical framework for LRRK2 variant re-evaluation. **(A)** Variant-level concordance between LRRK2 kinase activity measured in a standardized HEK293 overexpression assay (pThr73-Rab10 fold-change versus wild-type *LRRK2*) and LRRK2 pathway activation measured in patient-derived immune cells (pThr73-Rab10 fold-change versus controls) for selected *LRRK2* missense variants. Each symbol represents an individual variant (color-coded by protein domain; symbol size reflects the number of available individuals and/or biological replicates). Variants exhibiting stronger kinase activation in cellular assays are more likely to show detectable pathway activation in human biosamples. No statistical testing is shown due to limited sample sizes for rare variants. Previously published data for 21 G2019S and 21 R1441G carriers are included, with error bars indicating range ^18^. **(B)** Representative immunoblots from HEK293 cells transiently expressing wild-type *LRRK2* or selected strongly activating variants (Y1699C and F1700L), showing pThr73-Rab10 as a readout of kinase activity together with activating and kinase-inactive controls. **(C)** Representative immunoblots of pThr73-Rab10 in neutrophils derived from fresh peripheral blood from carriers of rare strongly activating *LRRK2* variants, with matched healthy controls. Total Rab10 and GAPDH are shown as loading controls. PD = variant carrier with manifesting Parkinson’s disease; NMC = non-manifesting carrier. **(D)** Functional stratification and framework for *LRRK2* variant re-evaluation. Robust and reproducible kinase activation in the HEK293 pThr73-Rab10 assay may contribute ACMG functional evidence category PS3 when interpreted in conjunction with independent genetic and clinical data. For variants of uncertain significance (VUS), such evidence may support re-evaluation toward likely pathogenic. Variants exhibiting wild-type–like or reduced kinase activity provide no evidence of kinase activation within this mechanistic context and should be interpreted cautiously. Detection of LRRK2 pathway activation in human biosamples provides supportive translational validation for subsets of strongly activating variants but is not required for functional classification.

Increased pThr73-Rab10 phosphorylation was detectable in peripheral blood immune cells from carriers of strongly activating variants identified in the cellular assay (Fig 6 A-C). Consistent with this, variants with wild-type–like activity in the HEK293 system did not show increased Rab10 phosphorylation in patient-derived samples (Fig 6A, supplemental data). At the individual variant level, greater kinase activation in the cellular assay was associated with greater detectability of pathway activation in peripheral blood immune cells (Fig 6A).

These findings demonstrate concordance between the magnitude of kinase activation observed in the standardized overexpression assay and detectability of LRRK2 pathway activation in clinically accessible immune cells. Variants with larger effect sizes were more likely to show measurable Rab10 phosphorylation ex vivo.

A schematic summarizing functional stratification and variant interpretation based on kinase activation status is presented in Fig 6 E.

### Functional - genetic integration for variant re-evaluation

To explore how calibrated functional data may inform variant interpretation, we examined selected variants currently classified as likely pathogenic or VUS in MDSGene and integrated quantitative kinase activity measurements with available genetic and clinical annotations (Supplementary Table 1).

Among likely pathogenic variants, a subset demonstrated robust kinase activation exceeding the predefined threshold and falling within the activation range observed for established pathogenic variants. When considered alongside available genetic evidence such as rarity in population databases, reported segregation, and phenotypic consistency, these variants exhibit convergent functional and genetic features compatible with potential re-evaluation toward pathogenic or likely pathogenic status within established classification frameworks.

In contrast, several variants currently categorized as likely pathogenic demonstrated wild-type-like kinase activity in the standardized assay. In the absence of functional evidence supporting kinase activation, the established disease mechanism in *LRRK2*-associated PD, such variants would require additional independent genetic data to justify retention of a likely pathogenic classification.

Within the VUS category, 41 variants exceeded the activation threshold, whereas the majority exhibited wild-type-like or reduced kinase activity. For activating VUS, integration of quantitative functional effect size with genetic evidence, including population frequency, segregation data, and clinical phenotype, may support reassessment in selected cases. However, functional evidence alone is generally insufficient for reclassification, and formal reassignment would require structured multidisciplinary review and comprehensive evidence integration beyond the scope of this study.

## Discussion

*LRRK2*-associated Parkinson’s disease represents a mechanistically defined gain-of-function disorder driven by increased kinase pathway activity. ^7–9^ Despite this biological clarity, interpretation of the rapidly expanding catalogue of *LRRK2* variants remains a major challenge, with the majority classified as VUS. ^11^

Here, we address this gap through large-scale functional annotation of *LRRK2* variants within a standardized experimental framework, enabling direct comparison of kinase activity across a broad spectrum of genetic variation. Our results define a continuous activation landscape and demonstrate that robust kinase activation aligns with established pathogenic variants, whereas discordance is largely confined to variants with limited genetic evidence.

Within a mechanistically defined gain-of-function disease ^7^, robust kinase activation provides functional evidence consistent with the established pathogenic model and may contribute meaningfully to variant interpretation when integrated with independent genetic and clinical data. ^14,15,19^ Conversely, absence of detectable activation in a Rab10-based assay does not exclude pathogenicity, as some disease-relevant effects may fall below assay sensitivity or operate through mechanisms not captured by this readout. ^35^ Functional evidence therefore operates asymmetrically in this context: clear activation is informative; whereas absence of activation warrants cautious interpretation.

Parallel re-expression of previously reported activating variants enabled direct comparison of activation magnitudes across structural domains within a unified framework. Activation strength was distributed along a continuum rather than as a binary state, with variants in the enzymatic core generally exhibiting larger effect sizes. The predefined ≥1.5-fold activation threshold applied here is operational and anchored in prior work; variants clustering near this boundary highlight the need for empirical recalibration. The graded activation spectrum raises the possibility that kinase effect size may influence penetrance, age at onset, or disease expressivity - hypotheses requiring systematic evaluation in larger well-phenotyped cohorts such as GP2 or PDGENEration. ^5,21,24^. The rarity of many variants highlights the challenges of variant interpretation based on genetic data alone and emphasizes the importance of complementary functional evidence. Large multi-ancestry resources such as GP2 will also be essential for interpreting variants within diverse ancestry-specific genetic backgrounds.

Positioned between the ROC GTPase and kinase catalytic modules, the COR scaffold is ideally placed to coordinate interdomain communication and to influence the conformational transitions that govern inactive–active state equilibrium. Our structural analyses indicate that the COR region is organised into four discrete subdomains that together form a composite allosteric scaffold controlling LRRK2 kinase activation. Structural elements within COR also participate in dimerization and in secondary interactions with the negative regulator 14-3-3, providing multiple routes by which local perturbations may alter kinase activation states.

Activating variants are not uniformly distributed across the COR region but cluster within specific structural elements, most prominently the DEP2 and PMM-like subdomains. This selective enrichment indicates that distinct structural elements within the COR scaffold contribute differentially to the regulation of kinase activity. Possible mechanisms include altered stability of LRRK2 dimerization, reduced association with 14-3-3 proteins, destabilisation of interdomain interactions that normally maintain the inactive conformation, changes in membrane engagement, metal-dependent conformational changes, binding of small-molecule-responsive regulatory elementor perturbation of ROC switch-region positioning. These effects are likely to shift the conformational landscape of LRRK2 towards more active states. Together, these findings provide a structural framework that begins to explain both the high frequency and mechanistic heterogeneity of activating variants within the ROC–COR region and identify this region as a central regulatory hub controlling LRRK2 signalling.

Validation in peripheral blood immune cells provides additional biological context. Variants demonstrating strong activation in the standardized assay are more likely to show detectable pathway activation in patient-derived neutrophils and monocytes, whereas variants displaying wild-type generally do not. Although detection in clinical biosamples is not required for functional interpretation, this correspondence reinforces the biological relevance of calibrated effect sizes measured in cellular systems and provides an important link between cellular functional assays and measurable pathway activation in human biosamples.

This framework should be viewed as foundational rather than definitive. Phosphorylation of pRab10 captures one dimension of LRRK2 signalling biology but does not encompass the full complexity of the pathway. Future refinement will require systematic evaluation of additional substrates, multiplexed phosphoproteomic analysis, and integration of lipid biomarkers such as bis(monoacylglycerol) phosphates, including intracellular species. ^35–37^ Mass spectrometry–based quantification may further improve sensitivity and dynamic range. Comprehensive deep mutational scanning of the 2,527–amino acid LRRK2 protein will enable data-driven mapping of activation landscapes and empirical recalibration of effect-size thresholds. Notably, ACMG PS3 criteria require well-established functional evidence rather than computational prediction ^16^. In this mechanistically defined context, experimentally measured kinase activation therefore provides critical evidence that cannot be substituted by in-silico scores.

From a translational perspective, establishing a calibrated activation reference scale for kinase activation has important implications for clinical development. As LRRK2-targeted therapies advance, mechanism-informed stratification strategies may become increasingly relevant. Quantitative assessment of pathway activation may help define biologically relevant subgroups both in monogenic LRRK2-associated PD and within subsets of idiopathic disease. In addition, it may serve as a pharmacodynamic readout to monitor target engagement in clinically accessible samples.

In summary, this study provides a quantitative framework for the functional interpretation of more than 350 *LRRK2* variants, grounded in a validated disease mechanism. By anchoring variant classification to calibrated kinase activation and integrating functional and structural, genetic and clinical data, this work bridges large-scale variant discovery with mechanism-driven precision medicine in PD. As such, this framework establishes a foundation for incorporating functional evidence into precision medicine strategies for PD.

## Supporting information

Supplementary material is available separately.

## Acknowledgements

We gratefully acknowledge all patients and volunteers for their participation in this study. We thank MRC Reagents and Services for outstanding technical support, and our collaborators and laboratory members for their valuable contributions and discussions. Figures were created using https://BioRender.com and further edited in Adobe Illustrator (2026). We particularly thank Dr Eugénie Mutez (Lille) for patient involvement and Dr Sara Gomes (Dundee) for helpful discussions.

This project was supported by the Global Parkinson’s Genetics Program (GP2; https://gp2.org). GP2 is funded by the Aligning Science Across Parkinson’s (ASAP) (https://ror.org/03zj4c476) initiative and implemented by The Michael J. Fox Foundation for Parkinson’s Research (MJFF) (https://ror.org/03arq3225). For a complete list of GP2 members, see https://doi.org/10.5281/zenodo.7904831.

## Funding

This work was supported by core funding from the MRC Core funding, a Carnegie Trust PhD studentship [PHD010656] (NP), and a Personal CSO Scottish Senior Clinical Academic Fellowship [SCAF/18/01] (ES).

This research was supported in part by the Intramural Research Program of the National Institutes of Health (NIH). The contributions of the NIH author(s) were made as part of their official duties as NIH federal employees, are in compliance with agency policy requirements, and are considered Works of the United States Government. However, the findings and conclusions presented in this paper are those of the author(s) and do not necessarily reflect the views of the NIH or the U.S. Department of Health and Human Services.

## Competing interests

The authors report no competing interests.

## Supplementary material

Supplementary material is available separately.

## Notes

### Competing Interest Statement

The authors have declared no competing interest.

### Author Declarations

The Ethics Committee of the Medical University of Vienna, Austria and the North East-Newcastle and North Tyneside 2 Research Ethics Committee, United Kingdom and the Ethics Committee of the University of Luebeck, Germany.

