## Supplementary material is available separately. for "Large-scale functional annotation establishes a reference framework for human *LRRK2* variants"

**Supplementary Table 1: Overview of LRRK2 variants reported in MDSGene [1] with available functional evidence from published studies [2, 3].**

| **#** | **Pos.** | **Variant** | **Domain** | **Variant type** | **MDS ACMG Classification** | **Functional data available** | **Functional outcome** |
| --- | --- | --- | --- | --- | --- | --- | --- |
| 1 | 10 | E10K | ARM | missense | VUS | [1] | Non-activating |
| 2 | 53 | K53R | ARM | missense | VUS | - | NA |
| 3 | 100 | M100T | ARM | missense | VUS | [1] | Non-activating |
| 4 | 115 | H115P | ARM | missense | Likely Benign | [1] | Non-activating |
| 5 | 116 | Q116R | ARM | missense | VUS | - | NA |
| 6 | 119 | L119P | ARM | missense | Likely Benign | [1] | Non-activating |
| 7 | 128 | S128G | ARM | missense | VUS | - | NA |
| 8 | 153 | L153W | ARM | missense | VUS | [1] | Non-activating |
| 9 | 173 | N173S | ARM | missense | VUS | - | NA |
| 10 | 192 | S192A | ARM | missense | VUS | - | NA |
| 11 | 193 | E193A | ARM | missense | VUS | - | NA |
| 12 | 193 | E193K | ARM | missense | VUS | - | NA |
| 13 | 211 | A211V | ARM | missense | VUS | [1] | Non-activating |
| 14 | 228 | C228S | ARM | missense | VUS | - | NA |
| 15 | 230 | H230R | ARM | missense | VUS | - | NA |
| 16 | 231 | S231P | ARM | missense | VUS | - | NA |
| 17 | 238 | N238I | ARM | missense | VUS | - | NA |
| 18 | 250 | Y250C | ARM | missense | VUS | - | NA |
| 19 | 255 | E255K | ARM | missense | VUS | - | NA |
| 20 | 258 | K258N | ARM | missense | VUS | - | NA |
| 21 | 259 | A259E | ARM | missense | VUS | - | NA |
| 22 | 262 | M262V | ARM | missense | Likely Benign | [1] | Non-activating |
| 23 | 286 | L286V | ARM | missense | VUS | - | NA |
| 24 | 291 | V291A | ARM | missense | VUS | - | NA |
| 25 | 306 | A306V | ARM | missense | VUS | - | NA |
| 26 | 308 | L308M | ARM | missense | VUS | - | NA |
| 27 | 312 | A312V | ARM | missense | VUS | - | NA |
| 28 | 320 | T320S | ARM | missense | VUS | - | NA |
| 29 | 335 | N335S | ARM | missense | VUS | - | NA |
| 30 | 370 | A370T | ARM | missense | VUS | - | NA |
| 31 | 378 | L378F | ARM | missense | VUS | - | NA |
| 32 | 379 | M379I | ARM | missense | VUS | - | NA |
| 33 | 388 | I388T | ARM | missense | VUS | [1] | Non-activating |
| 34 | 414 | V414I | ARM | missense | VUS | - | NA |
| 35 | 419 | A419V | ARM | missense | VUS | [1] | Activating |
| 36 | 425 | L425P | ARM | missense | VUS | - | NA |
| 37 | 437 | L437S | ARM | missense | VUS | - | NA |
| 38 | 438 | S438P | ARM | missense | VUS | - | NA |
| 39 | 459 | A459S | ARM | missense | VUS | [1] | Non-activating |
| 40 | 461 | S461I | ARM | missense | VUS | - | NA |
| 41 | 474 | N474S | ARM | missense | VUS | - | NA |
| 42 | 521 | R521G | ARM | missense | VUS | - | NA |
| 43 | 544 | K544E | ARM | missense | VUS | [1] | Non-activating |
| 44 | 579 | M579V | ARM | missense | VUS | - | NA |
| 45 | 610 | I610T | ARM | missense | VUS | - | NA |
| 46 | 616 | K616R | ARM | missense | VUS | [1] | Non-activating |
| 47 | 633 | S633F | ARM | missense | VUS | - | NA |
| 48 | 646 | T646A | ARM | missense | VUS | - | NA |
| 49 | 654 | A654V | ARM | missense | VUS | - | NA |
| 50 | 661 | S661F | ARM | missense | VUS | - | NA |
| 51 | 675 | I675M | ARM | missense | VUS | - | NA |
| 52 | 676 | F676S | ARM | missense | VUS | - | NA |
| 53 | 722 | S722N | ANK | missense | VUS | [1] | Non-activating |
| 54 | 724 | M724V | ANK | missense | VUS | - | NA |
| 55 | 734 | D734N | ANK | missense | VUS | - | NA |
| 56 | 739 | K739R | ANK | missense | VUS | - | NA |
| 57 | 755 | P755L | ANK | missense | Likely Benign | [1] | Non-activating |
| 58 | 764 | S764N | ANK | missense | VUS | - | NA |
| 59 | 767 | R767H | ANK | missense | Likely Pathogenic | [1] | Activating |
| 60 | 767 | R767C | ANK | missense | VUS | - | NA |
| 61 | 772 | R772* | ANK | nonsense | VUS | - | NA |
| 62 | 776 | T776M | ANK | missense | VUS | [1] | Non-activating |
| 63 | 777 | I777M | ANK | missense | VUS | - | NA |
| 64 | 784 | S784R | ANK | missense | VUS | - | NA |
| 65 | 786 | I786F | ANK | missense | VUS | - | NA |
| 66 | 792 | R792K | ANK | missense | VUS | [1] | Non-activating |
| 67 | 793 | R793M | ANK | missense | VUS | [1] | Non-activating |
| 68 | 827 | S827F | LRR | missense | VUS | - | NA |
| 69 | 844 | I844N | LRR | missense | VUS | - | NA |
| 70 | 848 | M848T | LRR | missense | VUS | - | NA |
| 71 | 858 | S858* | LRR | nonsense | VUS | - | NA |
| 72 | 865 | S865F | LRR | missense | Likely Benign | [1] | Non-activating |
| 73 | 885 | S885C | LRR | missense | VUS | - | NA |
| 74 | 899 | E899D | LRR | missense | VUS | - | NA |
| 75 | 922 | L922* | LRR | nonsense | VUS | - | NA |
| 76 | 923 | Q923H | LRR | missense | VUS | [1] | Non-activating |
| 77 | 925 | C925Y | LRR | missense | Likely Benign | [1] | Non-activating |
| 78 | 930 | Q930R | LRR | missense | VUS | [1] | Non-activating |
| 79 | 941 | D941Y | LRR | missense | VUS | - | NA |
| 80 | 944 | D944V | LRR | missense | VUS | [1] | Non-activating |
| 81 | 948 | R948Q | LRR | missense | VUS | - | NA |
| 82 | 958 | S958L | LRR | missense | VUS | - | NA |
| 83 | 972 | D972G | LRR | missense | VUS | - | NA |
| 84 | 973 | S973N | LRR | missense | VUS | [1] | Non-activating |
| 85 | 973 | S973G | LRR | missense | VUS | - | NA |
| 86 | 981 | R981K | LRR | missense | VUS | [1] | Activating |
| 87 | 1007 | S1007T | LRR | missense | VUS | [1] | Non-activating |
| 88 | 1019 | H1019R | LRR | missense | VUS | - | NA |
| 89 | 1029 | Q1029H | LRR | missense | VUS | - | NA |
| 90 | 1067 | R1067Q | LRR | missense | Likely Pathogenic | [1], [2] | Activating |
| 91 | 1089 | N1089S | LRR | missense | VUS | - | NA |
| 92 | 1096 | S1096C | LRR | missense | VUS | [1] | Non-activating |
| 93 | 1114 | L1114L | LRR | synonymous | VUS | - | NA |
| 94 | 1120 | S1120P | LRR | missense | VUS | - | NA |
| 95 | 1122 | I1122V | LRR | missense | VUS | [1] | Non-activating |
| 96 | 1128 | L1128M | LRR | missense | VUS | - | NA |
| 97 | 1151 | A1151T | LRR | missense | VUS | [1] | Non-activating |
| 98 | 1152 | C1152F | LRR | missense | VUS | - | NA |
| 99 | 1159 | S1159R | LRR | missense | VUS | - | NA |
| 100 | 1165 | L1165P | LRR | missense | VUS | - | NA |
| 101 | 1181 | S1181Y | LRR | missense | VUS | - | NA |
| 102 | 1192 | I1192V | LRR | missense | VUS | [1] | Non-activating |
| 103 | 1192 | I1192M | LRR | missense | VUS | - | NA |
| 104 | 1215 | A1215T | LRR | missense | VUS | - | NA |
| 105 | 1216 | H1216R | LRR | missense | VUS | - | NA |
| 106 | 1221 | N1221K | LRR | missense | Likely Pathogenic | - | NA |
| 107 | 1228 | S1228T | LRR | missense | VUS | [1] | Non-activating |
| 108 | 1271 | T1271I | LRR | missense | VUS | - | NA |
| 109 | 1281 | L1281L | LRR | synonymous | Likely Benign | - | NA |
| 110 | 1295 | W1295R | LRR | missense | VUS | - | NA |
| 111 | 1304 | L1304F | LRR | missense | VUS | - | NA |
| 112 | 1320 | R1320S | LRR | missense | VUS | [1] | Non-activating |
| 113 | 1325 | R1325Q | LRR | missense | Pathogenic | [1] | Activating |
| 114 | 1334 | R1334Q | LRR | missense | VUS | - | NA |
| 115 | 1339 | I1339M | LRR | missense | VUS | - | NA |
| 116 | 1340 | V1340M | LRR | missense | VUS | - | NA |
| 117 | 1353 | Q1353K | LRR | missense | VUS | - | NA |
| 118 | 1369 | V1369A | LRR | missense | VUS | - | NA |
| 119 | 1371 | I1371V | ROC | missense | Likely Benign | [1] | Non-activating |
| 120 | 1373 | V1373M | LRR | missense | VUS | - | NA |
| 121 | 1379 | Q1379R | LRR | missense | VUS | - | NA |
| 122 | 1388 | L1388I | LRR | missense | VUS | - | NA |
| 123 | 1389 | V1389I | LRR | missense | VUS | - | NA |
| 124 | 1398 | R1398C | LRR | missense | VUS | - | NA |
| 125 | 1398 | R1398R | LRR | synonymous | VUS | - | NA |
| 126 | 1402 | Y1402C | LRR | missense | VUS | - | NA |
| 127 | 1403 | S1403R | LRR | missense | Likely Pathogenic | - | NA |
| 128 | 1405 | H1405Q | LRR | missense | VUS | - | NA |
| 129 | 1413 | A1413T | LRR | missense | VUS | - | NA |
| 130 | 1427 | E1427G | LRR | missense | VUS | - | NA |
| 131 | 1436 | F1436L | ROC | missense | VUS | - | NA |
| 132 | 1437 | N1437H | ROC | missense | Pathogenic | [1] | Activating |
| 133 | 1437 | N1437D | ROC | missense | Likely Pathogenic | - | NA |
| 134 | 1437 | N1437S | ROC | missense | Likely Pathogenic | - | NA |
| 135 | 1440 | A1440P | ROC | missense | VUS | - | NA |
| 136 | 1441 | R1441C | ROC | missense | Pathogenic | [1] | Activating |
| 137 | 1441 | R1441G | ROC | missense | Pathogenic | [1] | Activating |
| 138 | 1441 | R1441H | ROC | missense | Pathogenic | [1] | Activating |
| 139 | 1441 | R1441S | ROC | missense | Pathogenic | [1] | Activating |
| 140 | 1441 | R1441R | ROC | synonymous | Likely Benign | - | NA |
| 141 | 1442 | A1442P | ROC | missense | Pathogenic | [1] | Activating |
| 142 | 1445 | S1445C | ROC | missense | VUS | - | NA |
| 143 | 1445 | S1445F | ROC | missense | VUS | - | NA |
| 144 | 1446 | P1446L | ROC | missense | VUS | - | NA |
| 145 | 1447 | V1447M | ROC | missense | Pathogenic | [1] | Activating |
| 146 | 1450 | V1450I | ROC | missense | VUS | - | NA |
| 147 | 1451 | G1451S | ROC | missense | VUS | - | NA |
| 148 | 1464 | A1464G | ROC | missense | VUS | [1] | Non-activating |
| 149 | 1468 | K1468E | ROC | missense | VUS | [1] | Non-activating |
| 150 | 1480 | P1480L | ROC | missense | VUS | - | NA |
| 151 | 1481 | A1481T | ROC | missense | VUS | - | NA |
| 152 | 1483 | R1483Q | ROC | missense | VUS | - | NA |
| 153 | 1490 | A1490V | ROC | missense | VUS | - | NA |
| 154 | 1492 | E1492K | ROC | missense | VUS | - | NA |
| 155 | 1501 | R1501W | ROC | missense | VUS | - | NA |
| 156 | 1508 | S1508G | ROC | missense | VUS | [1] | Non-activating |
| 157 | 1514 | R1514Q | COR-A | missense | Benign | - | NA |
| 158 | 1520 | G1520A | COR-A | missense | VUS | - | NA |
| 159 | 1527 | Y1527Y | COR-A | synonymous | Likely Benign | - | NA |
| 160 | 1541 | V1541M | COR-A | missense | VUS | - | NA |
| 161 | 1548 | I1548V | COR-A | missense | VUS | - | NA |
| 162 | 1552 | R1552* | COR-A | nonsense | VUS | - | NA |
| 163 | 1583 | L1583R | COR-A | missense | VUS | - | NA |
| 164 | 1589 | A1589S | COR-A | missense | VUS | [1] | Non-activating |
| 165 | 1613 | V1613A | COR-A | missense | VUS | [1] | Non-activating |
| 166 | 1615 | V1615V | COR-A | synonymous | VUS | - | NA |
| 167 | 1615 | V1615M | COR-A | missense | VUS | - | NA |
| 168 | 1616 | E1616K | COR-A | missense | VUS | - | NA |
| 169 | 1619 | P1619L | COR-A | missense | VUS | - | NA |
| 170 | 1620 | K1620R | COR-A | missense | VUS | - | NA |
| 171 | 1621 | H1621R | COR-A | missense | VUS | - | NA |
| 172 | 1621 | H1621Q | COR-A | missense | VUS | - | NA |
| 173 | 1622 | P1622L | COR-A | missense | VUS | - | NA |
| 174 | 1622 | P1622P | COR-A | synonymous | VUS | - | NA |
| 175 | 1622 | P1622T | COR-A | missense | VUS | - | NA |
| 176 | 1623 | K1623E | COR-A | missense | VUS | - | NA |
| 177 | 1626 | I1626V | COR-A | missense | VUS | - | NA |
| 178 | 1627 | S1627T | COR-A | missense | VUS | - | NA |
| 179 | 1627 | S1627L | COR-A | missense | VUS | - | NA |
| 180 | 1628 | R1628C | COR-A | missense | VUS | [1] | Non-activating |
| 181 | 1645 | Y1645S | COR-A | missense | VUS | - | NA |
| 182 | 1648 | Q1648R | COR-A | missense | VUS | - | NA |
| 183 | 1649 | Y1649S | COR-A | missense | VUS | - | NA |
| 184 | 1658 | I1658F | COR-A | missense | Likely Pathogenic | - | NA |
| 185 | 1661 | P1661T | COR-A | missense | VUS | - | NA |
| 186 | 1677 | R1677S | COR-B | missense | VUS | [1] | Non-activating |
| 187 | 1691 | I1691T | COR-B | missense | VUS | - | NA |
| 188 | 1693 | R1693Q | COR-B | missense | VUS | - | NA |
| 189 | 1699 | Y1699C | COR-B | missense | Pathogenic | [1] | Activating |
| 190 | 1700 | F1700L | COR-B | missense | Pathogenic | [3] | Activating |
| 191 | 1706 | S1706* | COR-B | nonsense | VUS | - | NA |
| 192 | 1707 | R1707K | COR-B | missense | VUS | - | NA |
| 193 | 1712 | L1712F | COR-B | missense | VUS | - | NA |
| 194 | 1725 | R1725Q | COR-B | missense | VUS | [1] | Non-activating |
| 195 | 1728 | R1728H | COR-B | missense | Likely Pathogenic | [1] | Activating |
| 196 | 1752 | S1752C | COR-B | missense | VUS | - | NA |
| 197 | 1756 | D1756G | COR-B | missense | VUS | - | NA |
| 198 | 1756 | D1756Y | COR-B | missense | VUS | - | NA |
| 199 | 1758 | H1758P | COR-B | missense | VUS | - | NA |
| 200 | 1761 | S1761R | COR-B | missense | Likely Pathogenic | [1] | Activating |
| 201 | 1771 | R1771T | COR-B | missense | VUS | - | NA |
| 202 | 1774 | C1774Y | COR-B | missense | VUS | - | NA |
| 203 | 1795 | L1795F | COR-B | missense | Pathogenic | [1] | Activating |
| 204 | 1806 | T1806I | COR-B | missense | VUS | - | NA |
| 205 | 1819 | G1819R | COR-B | missense | VUS | - | NA |
| 206 | 1823 | Q1823K | COR-B | missense | VUS | [1] | Non-activating |
| 207 | 1869 | M1869T | COR-B | missense | VUS | [1] | Non-activating |
| 208 | 1869 | M1869V | COR-B | missense | VUS | - | NA |
| 209 | 1873 | D1873H | COR-B | missense | VUS | - | NA |
| 210 | 1874 | E1874* | COR-B | nonsense | VUS | - | NA |
| 211 | 1887 | D1887N | KIN | missense | VUS | - | NA |
| 212 | 1891 | G1891A | KIN | missense | VUS | - | NA |
| 213 | 1914 | L1914I | KIN | missense | VUS | - | NA |
| 214 | 1941 | R1941H | KIN | missense | VUS | [1] | Non-activating |
| 215 | 1948 | E1948Q | KIN | missense | VUS | - | NA |
| 216 | 1954 | S1954F^1^ | KIN | missense | Likely Pathogenic | - | NA |
| 217 | 1961 | Q1961R | KIN | missense | VUS | - | NA |
| 218 | 1991 | I1991V | KIN | missense | VUS | [1] | Non-activating |
| 219 | 2006 | Y2006H | KIN | missense | VUS | [1] | Non-activating |
| 220 | 2010 | A2010T | KIN | missense | VUS | - | NA |
| 221 | 2012 | I2012T | KIN | missense | VUS | [1] | Non-activating |
| 222 | 2018 | Y2018Y | KIN | synonymous | Likely Benign | - | NA |
| 223 | 2019 | G2019S | KIN | missense | Pathogenic | [1] | Activating |
| 224 | 2020 | I2020T | KIN | missense | Pathogenic | [1] | Activating |
| 225 | 2020 | I2020L | KIN | missense | Likely Pathogenic | - | NA |
| 226 | 2020 | I2020S | KIN | missense | Likely Pathogenic | - | NA |
| 227 | 2031 | T2031S | KIN | missense | Likely Pathogenic | [1] | Activating |
| 228 | 2063 | L2063* | KIN | nonsense | VUS | - | NA |
| 229 | 2074 | V2074I | KIN | missense | VUS | - | NA |
| 230 | 2108 | E2108K | KIN | missense | VUS | - | NA |
| 231 | 2133 | N2133S | KIN | missense | VUS | - | NA |
| 232 | 2139 | C2139S | KIN | missense | VUS | - | NA |
| 233 | 2150 | V2150I | WD40 | missense | VUS | - | NA |
| 234 | 2168 | W2168L | WD40 | missense | VUS | - | NA |
| 235 | 2175 | D2175H | WD40 | missense | VUS | [1] | Non-activating |
| 236 | 2189 | Y2189C | WD40 | missense | VUS | [1] | Non-activating |
| 237 | 2230 | E2230K | WD40 | missense | VUS | - | NA |
| 238 | 2236 | H2236R | WD40 | missense | VUS | - | NA |
| 239 | 2251 | N2251T | WD40 | missense | VUS | - | NA |
| 240 | 2261 | N2261K | WD40 | missense | VUS | - | NA |
| 241 | 2294 | G2294R | WD40 | missense | VUS | - | NA |
| 242 | 2308 | N2308D | WD40 | missense | VUS | [1] | Non-activating |
| 243 | 2310 | T2310M | WD40 | missense | VUS | - | NA |
| 244 | 2313 | N2313S | WD40 | missense | VUS | [1] | Non-activating |
| 245 | 2323 | I2323F | WD40 | missense | VUS | - | NA |
| 246 | 2336 | I2336V | WD40 | missense | VUS | - | NA |
| 247 | 2350 | S2350I | WD40 | missense | VUS | [1] | Non-activating |
| 248 | 2356 | T2356I | WD40 | missense | VUS | [1] | Non-activating |
| 249 | 2390 | V2390M | WD40 | missense | VUS | [1] | Non-activating |
| 250 | 2390 | V2390A | WD40 | missense | VUS | - | NA |
| 251 | 2391 | H2391Q | WD40 | missense | Likely Benign | - | NA |
| 252 | 2395 | E2395K | WD40 | missense | VUS | - | NA |
| 253 | 2408 | M2408I | WD40 | missense | VUS | - | NA |
| 254 | 2423 | T2423S | WD40 | missense | VUS | - | NA |
| 255 | 2434 | I2434V | WD40 | missense | VUS | - | NA |
| 256 | 2439 | L2439I | WD40 | missense | VUS | [1] | Non-activating |
| 257 | 2461 | A2461V | WD40 | missense | VUS | - | NA |
| 258 | 2463 | L2463P | WD40 | missense | VUS | - | NA |
| 259 | 2490 | Q2490H | WD40 | missense | VUS | - | NA |
| 260 | 2494 | T2494I | WD40 | missense | VUS | [1] | Non-activating |
| 261 | 2495 | V2495I | WD40 | missense | VUS | - | NA |
| 262 | 2524 | T2524A | WD40 | missense | VUS | - | NA |
| 263 | 5000 | IVS5+33T>C | other | other | Benign | - | NA |
| 264 | 5000 | IVS19-10dupT | other | other | Benign | - | NA |
| 265 | 5000 | IVS4-68T>C | other | other | VUS | - | NA |
| 266 | 5000 | IVS8+1G>A | other | other | VUS | - | NA |
| 267 | 5000 | IVS19+5delGTAA | other | other | VUS | - | NA |
| 268 | 5000 | IVS31+3A>G | other | other | VUS | - | NA |
| 269 | 5000 | IVS33+78T>C | other | other | VUS | - | NA |
| 270 | 5000 | IVS33+79T>C | other | other | VUS | - | NA |
| 271 | 5000 | IVS36-29G>T | other | other | VUS | - | NA |
| 272 | 5000 | IVS4-38A>G | other | other | Likely Benign | - | NA |
| 273 | 5000 | IVS30+1G>C | other | other | VUS | - | NA |
| 274 | 5000 | IVS33+6T>A | other | other | VUS | - | NA |
| 275 | 5000 | IVS33+70T>C | other | other | VUS | - | NA |
| 276 | 5000 | IVS40-28A>T | other | other | VUS | - | NA |
| 277 | 5000 | IVS49+178A>G | other | other | VUS | - | NA |
| 278 | 5000 | Exon 49 deletion | other | other | VUS | - | NA |
| 279 | 5000 | K535Nfs*13 | other | other | VUS | - | NA |
| 280 | 5000 | R1639Kfs*13 | other | other | VUS | - | NA |
| 281 | 5000 | I1505Rfs*16 | other | other | VUS | - | NA |
| 282 | 5000 | K1772del | other | other | VUS | - | NA |
| 283 | 5000 | T1491del | other | other | VUS | - | NA |

Residue position, domain, variant type, MDSGene ACMG pathogenicity classification, availability of prior functional data, functional outcome (activating or non-activating) [1-3], and presence in the MDSGene browser. NA, not applicable; Refs., references; ACMG, American College of Medical Genetics and Genomics; ARM, armadillo; ANK, ankyrin; LRR, leucine-rich repeat; ROC, Ras of complex proteins; CORA/B, C-terminal of ROC subdomains A/B; KIN, kinase; WD40, WD40 repeat.

**Supplementary Table 2: All LRRK2 variants that had previously been functionally evaluated under standard cellular overexpression assay conditions [****1**-4**]**

| **Pos.** | **Variant** | **Domain** | **REVEL** | **Consurf-Score** | **Functional outcome** | **Refs.** | **Variant in MDSGene** | **MDS ACMG Classification** |
| --- | --- | --- | --- | --- | --- | --- | --- | --- |
| 10 | E10K | ARM | 0.207 | 1 |  | [1] | ✓ | VUS |
| 100 | M100T | ARM | 0.209 | 2 |  | [1] | ✓ | VUS |
| 115 | H115P | ARM | 0.566 | 9 |  | [1] | ✓ | likely benign |
| 119 | L119P | ARM | 0.464 | 8 |  | [1] | ✓ | VUS |
| 153 | L153W | ARM | 0.277 | 8 |  | [1] | ✓ | VUS |
| 211 | A211V | ARM | 0.206 | 6 |  | [1] | ✓ | VUS |
| 262 | M262V | ARM | 0.013 | 1 |  | [1] | ✓ | VUS |
| 334 | E334K | ARM | 0.194 | 6 | Activating | [1] | - | - |
| 363 | N363S | ARM | 0.039 | 5 |  | [1] | - | - |
| 388 | I388T | ARM | 0.161 | 7 |  | [1] | ✓ | VUS |
| 393 | G393V | ARM | 0.124 | 1 |  | [1] | - | - |
| 419 | A419V | ARM | 0.175 | 7 | Activating | [1] | ✓ | VUS |
| 459 | A459S | ARM | 0.089 | 6 |  | [1] | ✓ | VUS |
| 478 | D478Y | ARM | 0.369 | 9 |  | [1] | - | - |
| 479 | I479V | ARM | 0.026 | 1 |  | [1] | - | - |
| 544 | K544E | ARM | 0.288 | 2 |  | [1] | ✓ | VUS |
| 551 | N551K | ARM | 0.248 | 9 |  | [1] | - | - |
| 616 | K616R | ARM | 0.264 | 6 |  | [1] | ✓ | VUS |
| 712 | M712V | ANK | 0.228 | 6 |  | [1] | - | - |
| 722 | S722N | ANK | 0.098 | 5 |  | [1] | ✓ | VUS |
| 723 | I723V | ANK | 0.045 | 1 |  | [1] | - | - |
| 755 | P755L | ANK | 0.173 | 5 |  | [1] | ✓ | VUS |
| 767 | R767H | ANK | 0.157 | 7 | Activating | [1] | ✓ | likely pathogenic |
| 776 | T776M | ANK | 0.046 | 1 |  | [1] | ✓ | VUS |
| 792 | R792K | ANK | 0.023 | 1 |  | [1] | ✓ | VUS |
| 793 | R793M | ANK | 0.305 | 8 |  | [1] | ✓ | VUS |
| 810 | I810V | LRR | 0.016 | 2 |  | [1] | ✓ | VUS |
| 865 | S865F | LRR | 0.149 | 1 |  | [1] | ✓ | VUS |
| 885 | S885N | LRR | 0.025 | 2 |  | [1] | - | - |
| 923 | Q923H | LRR | 0.272 | 1 |  | [1] | ✓ | VUS |
| 925 | C925Y | LRR | 0.039 | 1 |  | [1] | ✓ | VUS |
| 930 | Q930R | LRR | 0.286 | 1 |  | [1] | ✓ | VUS |
| 944 | D944V | LRR | 0.140 | 1 |  | [1] | ✓ | VUS |
| 973 | S973N | LRR | 0.032 | 6 |  | [1] | ✓ | VUS |
| 981 | R981K | LRR | 0.062 | 3 | Activating | [1] | ✓ | VUS |
| 1007 | S1007T | LRR | 0.054 | 2 |  | [1] | ✓ | VUS |
| 1067 | R1067Q | LRR | 0.282 | 8 | Activating | [1], [2] | ✓ | likely pathogenic |
| 1096 | S1096C | LRR | 0.204 | 2 |  | [1] | ✓ | VUS |
| 1111 | Q1111H | LRR | 0.219 | 8 |  | [1] | - | - |
| 1122 | I1122V | LRR | 0.244 | 3 |  | [1] | ✓ | VUS |
| 1138 | K1138E | LRR | 0.354 | 7 |  | [1] | - | - |
| 1151 | A1151T | LRR | 0.029 | 1 |  | [1] | ✓ | VUS |
| 1192 | I1192V | LRR | 0.215 | 5 |  | [1] | ✓ | VUS |
| 1228 | S1228T | LRR | 0.307 | 7 |  | [1] | ✓ | VUS |
| 1320 | R1320S | LRR | 0.319 | 9 |  | [1] | ✓ | VUS |
| 1325 | R1325Q | LRR | 0.553 | 9 | Activating | [1] | ✓ | Pathogenic |
| 1371 | I1371V | ROC | 0.453 | 8 |  | [1] | ✓ | VUS |
| 1398 | R1398H | ROC | 0.369 | 7 |  | [1] | - | - |
| 1415 | Y1415E | ROC | - | 6 | Non-activating |  | - | - |
| 1417 | A1417V | ROC | 0.199 | 8 | Activating |  | - | - |
| 1417 | A1417E | ROC | - | 8 | Non-activating |  | - | - |
| 1435 | L1435F | ROC | 0.746 | 9 | Activating |  | - | - |
| 1435 | L1435E | ROC | - | 9 | Non-activating |  | - | - |
| 1437 | N1437H | ROC | 0.603 | 9 | Activating | [1] | ✓ | Pathogenic |
| 1438 | I1438E | ROC | - | 9 | Activating |  | - | - |
| 1438 | I1438T | ROC | 0.948 | 9 | Activating |  | - | - |
| 1438 | I1438V | ROC | 0.582 | 9 | Non-activating |  | - | - |
| 1441 | R1441S | ROC | 0.660 | 7 | Activating | [1] | ✓ | Pathogenic |
| 1441 | R1441H | ROC | 0.635 | 7 | Activating | [1] | ✓ | Pathogenic |
| 1441 | R1441C | ROC | 0.727 | 7 | Activating | [1] | ✓ | Pathogenic |
| 1441 | R1441G | ROC | 0.705 | 7 | Activating | [1] | ✓ | Pathogenic |
| 1442 | A1442P | ROC | 0.783 | 9 | Activating | [1] | ✓ | Pathogenic |
| 1447 | V1447M | ROC | 0.749 | 9 | Activating | [1] | ✓ | Pathogenic |
| 1447 | V1447L | ROC | - | 9 | Activating | [4] | - | - |
| 1447 | V1447G | ROC | 0.941 | 9 | Activating |  | - | - |
| 1447 | V1447E | ROC | 0.948 | 9 | Non-activating |  | - | - |
| 1464 | A1464G | ROC | 0.209 | 3 |  | [1] | ✓ | VUS |
| 1468 | K1468E | ROC | 0.585 | 9 |  | [1] | ✓ | VUS |
| 1508 | S1508R | ROC | 0.522 | 6 |  | [1] | - | - |
| 1508 | S1508G | ROC | 0.390 | 6 |  | [1] | ✓ | VUS |
| 1514 | R1514G | CORA | 0.354 | 9 |  | [1] | - | - |
| 1514 | R1514Q | CORA | 0.100 | 9 |  | [1] | - | - |
| 1542 | P1542S | CORA | 0.228 | 4 |  | [1] | - | - |
| 1589 | A1589S | CORA | 0.253 | 8 |  | [1] | ✓ | VUS |
| 1613 | V1613A | CORA | 0.585 | 5 |  | [1] | ✓ | VUS |
| 1628 | R1628C | CORA | 0.451 | 8 |  | [1] | ✓ | VUS |
| 1628 | R1628P | CORA | 0.546 | 8 | Activating | [1] | - | - |
| 1646 | M1646T | CORA | 0.184 | 3 |  | [1] | - | - |
| 1647 | S1647T | CORA | 0.086 | 2 |  | [1] | - | - |
| 1677 | R1677S | CORB | 0.386 | 7 |  | [1] | ✓ | VUS |
| 1699 | Y1699C | CORB | 0.870 | 9 | Activating | [1] | ✓ | Pathogenic |
| 1700 | F1700L | COR-B | - | 9 | Activating | [3] | ✓ | Pathogenic |
| 1725 | R1725Q | CORB | 0.095 | 6 |  | [1] | ✓ | VUS |
| 1728 | R1728L | CORB | 0.763 | 8 | Activating | [1] | - | - |
| 1728 | R1728H | CORB | 0.721 | 8 | Activating | [1] | ✓ | likely pathogenic |
| 1761 | S1761R | CORB | 0.521 | 8 | Activating | [1] | ✓ | likely pathogenic |
| 1795 | L1795F | CORB | 0.638 | 9 | Activating | [1] | ✓ | Pathogenic |
| 1823 | Q1823K | CORB | 0.188 | 2 |  | [1] | - | - |
| 1869 | M1869T | CORB | 0.514 | 6 |  | [1] | - | - |
| 1941 | R1941H | KIN | 0.240 | 4 |  | [1] | - | - |
| 1991 | I1991V | KIN | 0.448 | 8 |  | [1] | - | - |
| 2006 | Y2006H | KIN | 0.301 | 8 |  | [1] | - | - |
| 2012 | I2012T | KIN | 0.664 | 8 |  | [1] | - | - |
| 2019 | G2019S | KIN | 0.970 | 9 | Activating | [1] | yes | Pathogenic |
| 2020 | I2020T | KIN | 0.957 | 9 | Activating | [1] | yes | Pathogenic |
| 2031 | T2031S | KIN | 0.280 | 9 | Activating | [1] | ✓ | likely pathogenic |
| 2081 | N2081D | KIN | 0.138 | 9 | Activating | [1] | - | - |
| 2141 | T2141M | KIN | 0.342 | 5 |  | [1] | - | - |
| 2143 | R2143H | WD40 | 0.277 | 1 |  | [1] | - | - |
| 2143 | R2143M | WD40 | N/A | 1 |  | [1] | - | - |
| 2175 | D2175H | WD40 | 0.307 | 1 |  | [1] | ✓ | VUS |
| 2189 | Y2189C | WD40 | 0.394 | 1 |  | [1] | ✓ | VUS |
| 2308 | N2308D | WD40 | 0.025 | 1 |  | [1] | ✓ | VUS |
| 2313 | N2313S | WD40 | 0.043 | 3 |  | [1] | ✓ | VUS |
| 2350 | S2350I | WD40 | 0.121 | 5 |  | [1] | ✓ | VUS |
| 2356 | T2356I | WD40 | 0.154 | 6 |  | [1] | ✓ | VUS |
| 2385 | G2385R | WD40 | 0.044 | 5 | Activating | [1] | - | - |
| 2390 | V2390M | WD40 | 0.205 | 4 |  | [1] | ✓ | VUS |
| 2397 | M2397T | WD40 | 0.098 | 1 |  | [1] | - | - |
| 2439 | L2439I | WD40 | 0.135 | 7 |  | [1] | ✓ | VUS |
| 2466 | L2466H | WD40 | 0.311 | 2 |  | [1] | - | - |
| 2494 | T2494I | WD40 | 0.065 | 5 |  | [1] | ✓ | VUS |

Variants previously evaluated under standard cellular overexpression assay conditions [2,3]. Pos., position; Refs., references; ACMG, American College of Medical Genetics and Genomics; VUS, variant of uncertain significance.

**Supplementary Table 3. Summary of functional data for all LRRK2 variants that have functionally evaluated under standard conditions.**

| **#** | **Pos.** | **Variant** | **Domain** | **Variant type** | **Consurf score** | **MDS**  **ACMG** | **Published functional outcome** | **Current study outcome** | **Mean**  **pRAB10/**  **RAB10**  **(all studies)** | **SD** | **Repl.**  **(n)** |
| --- | --- | --- | --- | --- | --- | --- | --- | --- | --- | --- | --- |
| 1 | 10 | E10K | ARM | missense | 4 | VUS | Non-activating | Non-activating | 0.99 | 0.34 | 7 |
| 2 | 53 | K53R | ARM | missense | 4 | VUS |  | Non-activating | 1.30 | 0.34 | 3 |
| 3 | 80 | V80M | ARM | missense | 5 | N/A |  | Non-activating | 0.81 | 0.15 | 3 |
| 4 | 96 | M96T | ARM | missense | 5 | N/A |  | Non-activating | 0.89 | 0.36 | 3 |
| 5 | 100 | M100T | ARM | missense | 5 | VUS | Non-activating | Non-activating | 1.44 | 0.38 | 5 |
| 6 | 104 | D104Y | ARM | missense | 5 | N/A |  | Non-activating | 0.90 | 0.04 | 2 |
| 7 | 115 | H115P | ARM | missense | 8 | Likely benign | Non-activating | Non-activating | 1.44 | 0.38 | 5 |
| 8 | 116 | Q116R | ARM | missense | 5 | VUS |  | Non-activating | 0.89 | 0.01 | 3 |
| 9 | 119 | L119P | ARM | missense | 6 | Likely benign | Non-activating | Non-activating | 0.80 | 0.27 | 8 |
| 10 | 128 | S128G | ARM | missense | 5 | VUS |  | Non-activating | 1.03 | 0.07 | 3 |
| 11 | 149 | T149S | ARM | missense | 7 | N/A |  | Non-activating | 0.91 | 0.04 | 3 |
| 12 | 153 | L153W | ARM | missense | 9 | VUS | Non-activating | Non-activating | 1.41 | 0.30 | 5 |
| 13 | 160 | F160L | ARM | missense | 9 | N/A |  | Non-activating | 1.23 | 0.31 | 3 |
| 14 | 173 | N173S | ARM | missense | 5 | VUS |  | Non-activating | 0.98 | 0.10 | 3 |
| 15 | 176 | V176L | ARM | missense | 5 | N/A |  | Non-activating | 0.69 | 0.37 | 3 |
| 16 | 192 | S192A | ARM | missense | 8 | VUS |  | Non-activating | 1.46 | 0.62 | 2 |
| 17 | 193 | E193A | ARM | missense | 9 | VUS |  | Non-activating | 0.75 | 0.19 | 3 |
| 18 | 193 | E193K | ARM | missense | 9 | VUS |  | Non-activating | 1.41 | 0.35 | 11 |
| 19 | 211 | A211V | ARM | missense | 5 | VUS | Non-activating | Non-activating | 1.22 | 0.36 | 6 |
| 20 | 228 | C228S | ARM | missense | 8 | VUS |  | Non-activating | 1.08 | 0.11 | 3 |
| 21 | 230 | H230R | ARM | missense | 5 | VUS |  | Activating | 3.43 | 1.20 | 16 |
| 22 | 231 | S231P | ARM | missense | 8 | VUS |  | Non-activating | 0.85 | 0.05 | 2 |
| 23 | 238 | N238I | ARM | missense | 8 | VUS |  | Non-activating | 1.14 | 0.71 | 3 |
| 24 | 250 | Y250C | ARM | missense | 9 | VUS |  | Non-activating | 0.70 | 0.05 | 5 |
| 25 | 255 | E255K | ARM | missense | 5 | VUS |  | Non-activating | 1.07 | 0.15 | 4 |
| 26 | 258 | K258N | ARM | missense | 4 | VUS |  | Non-activating | 0.83 | 0.08 | 3 |
| 27 | 259 | A259E | ARM | missense | 3 | VUS |  | Non-activating | 1.14 | 0.26 | 3 |
| 28 | 262 | M262V | ARM | missense | 1 | Likely benign | Non-activating | Non-activating | 0.89 | 0.47 | 5 |
| 29 | 286 | L286V | ARM | missense | 9 | VUS |  | Non-activating | 1.11 | 0.65 | 3 |
| 30 | 291 | V291A | ARM | missense | 9 | VUS |  | Non-activating | 0.77 | 0.05 | 3 |
| 31 | 292 | H292Q | ARM | missense | 8 | N/A |  | Non-activating | 0.74 | 0.19 | 3 |
| 32 | 306 | A306V | ARM | missense | 6 | VUS |  | Non-activating | 0.61 | 0.49 | 4 |
| 33 | 308 | L308M | ARM | missense | 8 | VUS |  | Non-activating | 1.00 | 0.10 | 4 |
| 34 | 312 | A312V | ARM | missense | 6 | VUS |  | Activating | 1.46 | 0.62 | 6 |
| 35 | 313 | L313F | ARM | missense | 9 | N/A |  | Non-activating | 0.75 | 0.32 | 3 |
| 36 | 320 | T320S | ARM | missense | 9 | VUS |  | Non-activating | 1.22 | 0.48 | 6 |
| 37 | 334 | E334K | ARM | missense | 5 | N/A |  | Activating | 1.73 | 0.76 | 22 |
| 38 | 335 | N335S | ARM | missense | 3 | VUS |  | Non-activating | 1.05 | 0.10 | 3 |
| 39 | 353 | C353Y | ARM | missense | 8 | N/A |  | Non-activating | 0.79 | 0.22 | 3 |
| 40 | 356 | A356T | ARM | missense | 9 | N/A |  | Activating | 2.64 | 0.75 | 9 |
| 41 | 363 | N363S | ARM | missense | 6 | N/A |  | Non-activating | 1.03 | 0.60 | 5 |
| 42 | 365 | H365N | ARM | missense | 4 | N/A |  | Non-activating | 0.89 | 0.15 | 5 |
| 43 | 370 | A370T | ARM | missense | 9 | VUS |  | Non-activating | 1.07 | 0.59 | 3 |
| 44 | 378 | L378F | ARM | missense | 8 | VUS |  | Activating | 1.82 | 0.33 | 9 |
| 45 | 379 | M379I | ARM | missense | 7 | VUS |  | Non-activating | 1.23 | 0.10 | 3 |
| 46 | 385 | H385R | ARM | missense | 8 | N/A |  | Non-activating | 0.65 | 0.09 | 5 |
| 47 | 388 | I388T | ARM | missense | 8 | VUS | Non-activating | Non-activating | 0.72 | 0.30 | 5 |
| 48 | 397 | A397T | ARM | missense | 5 | N/A | Non-activating | Non-activating | 0.93 | 0.31 | 2 |
| 49 | 414 | V414I | ARM | missense | 6 | VUS |  | Activating | 2.55 | 1.12 | 17 |
| 50 | 419 | A419V | ARM | missense | 8 | VUS | Activating | Activating | 1.79 | 0.93 | 22 |
| 51 | 425 | L425P | ARM | missense | 9 | VUS |  | Non-activating | 0.97 | 0.21 | 3 |
| 52 | 437 | L437S | ARM | missense | 7 | VUS |  | Non-activating | 1.16 | 0.50 | 9 |
| 53 | 438 | S438P | ARM | missense | 7 | VUS |  | Non-activating | 0.79 | 0.03 | 3 |
| 54 | 442 | H442Y | ARM | missense | 5 | N/A |  | Non-activating | 1.13 | 0.10 | 3 |
| 55 | 455 | S455P | ARM | missense | 3 | N/A |  | Activating | 2.87 | 0.95 | 13 |
| 56 | 459 | A459S | ARM | missense | 9 | VUS | Non-activating | Non-activating | 1.17 | 0.54 | 5 |
| 57 | 461 | S461I | ARM | missense | 9 | VUS |  | Activating | 1.61 | 0.50 | 12 |
| 58 | 474 | N474S | ARM | missense | 3 | VUS |  | Non-activating | 0.93 | 0.30 | 3 |
| 59 | 478 | D478Y | ARM | missense | 9 | N/A |  | Non-activating | 1.39 | 0.45 | 5 |
| 60 | 479 | I479V | ARM | missense | 4 | N/A |  | Non-activating | 1.15 | 0.34 | 5 |
| 61 | 488 | L488P |  | missense |  | N/A |  | Activating | 2.00 | 0.00 |  |
| 62 | 504 | A504T | ARM | missense | 8 | N/A |  | Activating | 2.68 | 1.71 | 13 |
| 63 | 506 | R506P | ARM | missense | 6 | N/A |  | Activating | 1.78 | 0.68 | 17 |
| 64 | 506 | R506Q | ARM | missense | 6 | N/A |  | Non-activating | 1.12 | 0.11 | 3 |
| 65 | 518 | E518K | ARM | missense | 5 | N/A |  | Non-activating | 1.21 | 0.36 | 6 |
| 66 | 521 | R521G | ARM | missense | 1 | VUS |  | Non-activating | 1.14 | 0.14 | 3 |
| 67 | 544 | K544E | ARM | missense | 5 | VUS | Non-activating | Non-activating | 0.82 | 0.54 | 5 |
| 68 | 551 | N551K | ARM | missense | 5 | N/A |  | Non-activating | 0.62 | 0.42 | 5 |
| 69 | 581 | S581T | ARM | missense | 5 | N/A |  | Non-activating | 0.99 | 0.22 | 5 |
| 70 | 610 | I610T | ARM | missense | 4 | VUS |  | Non-activating | 1.30 | 0.55 | 9 |
| 71 | 616 | K616R | ARM | missense | 5 | VUS | Non-activating | Non-activating | 1.03 | 0.67 | 6 |
| 72 | 633 | S633F | ARM | missense | 8 | VUS |  | Non-activating | 1.04 | 0.52 | 6 |
| 73 | 637 | R637Q | ARM | missense | 9 | N/A |  | Activating | 2.80 | 1.29 | 17 |
| 74 | 654 | A654V | ARM | missense | 3 | VUS |  | Non-activating | 0.88 | 0.06 | 3 |
| 75 | 661 | S661F | ARM | missense | 4 | VUS |  | Non-activating | 0.70 | 0.13 | 3 |
| 76 | 675 | I675M | ARM | missense | 5 | VUS |  | Non-activating | 0.78 | 0.03 | 3 |
| 77 | 691 | F691S | ARM | missense | 4 | N/A |  | Non-activating | 0.63 | 0.21 | 2 |
| 78 | 712 | M712V | ANK | missense | 4 | N/A |  | Non-activating | 0.94 | 0.61 | 7 |
| 79 | 722 | S722N | ANK | missense | 7 | VUS | Non-activating | Non-activating | 0.74 | 0.33 | 5 |
| 80 | 723 | I723V | ANK | missense | 3 | N/A |  | Non-activating | 0.82 | 0.49 | 5 |
| 81 | 724 | M724V | ANK | missense | 8 | VUS |  | Non-activating | 1.14 | 0.12 | 3 |
| 82 | 734 | D734N | ANK | missense | 9 | VUS |  | Non-activating | 0.80 | 0.15 | 5 |
| 83 | 739 | K739R | ANK | missense | 4 | VUS |  | Non-activating | 0.74 | 0.13 | 3 |
| 84 | 755 | P755L | ANK | missense | 7 | Likely benign | Non-activating | Non-activating | 0.80 | 0.34 | 6 |
| 85 | 757 | L757V | ANK | missense | 8 | N/A |  | Non-activating | 0.74 | 0.16 | 4 |
| 86 | 764 | S764N | ANK | missense | 3 | VUS |  | Non-activating | 1.15 | 0.39 | 3 |
| 87 | 767 | R767H | ANK | missense | 4 | Likely pathogenic | Activating | Non-activating | 1.29 | 0.67 | 18 |
| 88 | 767 | R767C | ANK | missense | 4 | VUS |  | Non-activating | 0.95 | 0.61 | 2 |
| 89 | 776 | T776M | ANK | missense | 3 | VUS | Non-activating | Non-activating | 1.03 | 0.70 | 8 |
| 90 | 777 | I777M | ANK | missense | 9 | VUS |  | Non-activating | 0.99 | 0.14 | 3 |
| 91 | 784 | S784R | ANK | missense | 7 | VUS |  | Non-activating | 1.17 | 0.03 | 3 |
| 92 | 786 | I786F | ANK | missense | 3 | VUS |  | Non-activating | 1.04 | 0.10 | 3 |
| 93 | 792 | R792K | ANK | missense | 4 | VUS | Non-activating | Non-activating | 1.05 | 0.13 | 4 |
| 94 | 793 | R793M | ANK | missense | 8 | VUS | Non-activating | Non-activating | 0.81 | 0.20 | 5 |
| 95 | 810 | I810V | LRR | missense | 3 | N/A |  | Non-activating | 0.83 | 0.57 | 5 |
| 96 | 812 | K812N | LRR | missense | 6 | N/A |  | Non-activating | 1.25 | 0.06 | 2 |
| 97 | 817 | W817C | LRR | missense | 8 | N/A |  | Non-activating | 1.33 | 0.36 | 5 |
| 98 | 827 | S827F | LRR | missense | 4 | VUS |  | Non-activating | 1.05 | 0.16 | 3 |
| 99 | 844 | I844N | LRR | missense | 5 | VUS |  | Non-activating | 1.24 | 0.37 | 3 |
| 100 | 848 | M848T | LRR | missense | 3 | VUS |  | Non-activating | 1.22 | 0.07 | 3 |
| 101 | 860 | S860R | LRR | missense | 5 | N/A |  | Non-activating | 0.95 | 0.21 | 5 |
| 102 | 865 | S865F | LRR | missense | 3 | Likely benign | Non-activating | Non-activating | 1.08 | 0.75 | 5 |
| 103 | 867 | D867N | LRR | missense | 4 | N/A |  | Non-activating | 0.92 | 0.18 | 5 |
| 104 | 885 | S885N | LRR | missense | 4 | N/A |  | Non-activating | 1.23 | 0.90 | 8 |
| 105 | 885 | S885C | LRR | missense | 4 | VUS |  | Non-activating | 0.85 | 0.16 | 3 |
| 106 | 899 | E899D | LRR | missense | 7 | VUS |  | Non-activating | 0.92 | 0.18 | 5 |
| 107 | 918 | R918Q | LRR | missense | 2 | N/A |  | Non-activating | 0.92 | 0.18 | 5 |
| 108 | 923 | Q923H | LRR | missense | 7 | VUS | Non-activating | Non-activating | 1.37 | 0.62 | 6 |
| 109 | 925 | C925Y | LRR | missense | 8 | Likely benign | Non-activating | Non-activating | 1.01 | 0.26 | 5 |
| 110 | 930 | Q930R | LRR | missense | 4 | VUS | Non-activating | Non-activating | 1.08 | 0.45 | 5 |
| 111 | 931 | R931K | LRR | missense | 5 | N/A |  | Non-activating | 0.86 | 0.02 | 3 |
| 112 | 941 | D941Y | LRR | missense | 6 | VUS |  | Non-activating | 0.90 | 0.07 | 3 |
| 113 | 944 | D944V | LRR | missense | 5 | VUS | Non-activating | Non-activating | 0.83 | 0.19 | 5 |
| 114 | 948 | R948Q | LRR | missense | 3 | VUS |  | Non-activating | 0.68 | 0.07 | 3 |
| 115 | 954 | S954T | LRR | missense | 5 | N/A |  | Non-activating | 1.23 | 0.36 | 2 |
| 116 | 958 | S958L | LRR | missense | 6 | VUS |  | Non-activating | 0.83 | 0.16 | 3 |
| 117 | 969 | R969K | LRR | missense | 5 | N/A |  | Non-activating | 0.76 | 0.28 | 5 |
| 118 | 972 | D972G | LRR | missense | 3 | VUS |  | Non-activating | 0.93 | 0.18 | 3 |
| 119 | 973 | S973G | LRR | missense | 9 | VUS |  | Non-activating | 1.28 | 0.29 | 3 |
| 120 | 973 | S973N | LRR | missense | 9 | VUS | Non-activating | Non-activating | 1.18 | 0.32 | 7 |
| 121 | 981 | R981K | LRR | missense | 7 | VUS | Activating | Non-activating | 1.20 | 0.53 | 11 |
| 122 | 984 | I984V | LRR | missense | 9 | N/A |  | Non-activating | 0.64 | 0.08 | 3 |
| 123 | 1007 | S1007T | LRR | missense | 4 | VUS | Non-activating | Non-activating | 0.84 | 0.44 | 5 |
| 124 | 1015 | K1015R | LRR | missense | 5 | N/A | Non-activating | Non-activating | 0.65 | 0.07 | 3 |
| 125 | 1019 | H1019R | LRR | missense | 5 | VUS |  | Activating | 1.54 | 0.39 | 12 |
| 126 | 1029 | Q1029H | LRR | missense | 4 | VUS |  | Non-activating | 0.93 | 0.14 | 3 |
| 127 | 1067 | R1067Q | LRR | missense | 9 | VUS | Activating | Activating | 2.21 | 0.85 | 24 |
| 128 | 1089 | N1089S | LRR | missense | 8 | VUS |  | Non-activating | 0.60 | 0.08 | 3 |
| 129 | 1096 | S1096C | LRR | missense | 5 | VUS | Non-activating | Non-activating | 0.93 | 0.33 | 5 |
| 130 | 1099 | P1099S | LRR | missense | 9 | N/A |  | Non-activating | 0.92 | 0.06 | 2 |
| 131 | 1111 | Q1111H | LRR | missense | 6 | N/A |  | Non-activating | 0.78 | 0.28 | 5 |
| 132 | 1122 | I1122V | LRR | missense | 4 | VUS | Non-activating | Non-activating | 0.60 | 0.28 | 5 |
| 133 | 1128 | L1128M | LRR | missense | 7 | VUS |  | Non-activating | 0.61 | 0.12 | 3 |
| 134 | 1132 | K1132E | LRR | missense | 8 | N/A |  | Non-activating | 0.94 | 0.38 | 7 |
| 135 | 1138 | K1138E | LRR | missense | 9 | N/A |  | Non-activating | 1.10 | 0.63 | 5 |
| 136 | 1141 | I1141V | LRR | missense | 9 | N/A |  | Non-activating | 1.19 | 0.44 | 2 |
| 137 | 1151 | A1151T | LRR | missense | 4 | VUS | Non-activating | Non-activating | 0.60 | 0.35 | 5 |
| 138 | 1152 | C1152F | LRR | missense | 5 | VUS |  | Non-activating | 0.67 | 0.20 | 3 |
| 139 | 1159 | S1159R | LRR | missense | 5 | VUS |  | Non-activating | 1.07 | 0.14 | 3 |
| 140 | 1160 | A1160D | LRR | missense | 9 | N/A |  | Non-activating | 0.84 | 0.01 | 2 |
| 141 | 1165 | L1165P | LRR | missense | 7 | VUS |  | Non-activating | 0.85 | 0.23 | 4 |
| 142 | 1181 | S1181Y | LRR | missense | 8 | VUS |  | Non-activating | 0.74 | 0.19 | 3 |
| 143 | 1192 | I1192V | LRR | missense | 7 | VUS | Non-activating | Non-activating | 0.82 | 0.30 | 5 |
| 144 | 1192 | I1192M | LRR | missense | 7 | VUS |  | Non-activating | 0.48 | 0.11 | 5 |
| 145 | 1215 | A1215T | LRR | missense | 4 | VUS |  | Non-activating | 0.89 | 0.13 | 2 |
| 146 | 1216 | H1216R | LRR | missense | 5 | VUS |  | Non-activating | 0.99 | 0.09 | 3 |
| 147 | 1228 | S1228T | LRR | missense | 5 | VUS | Non-activating | Non-activating | 1.05 | 0.35 | 5 |
| 148 | 1262 | P1262A | LRR | missense | 6 | N/A |  | Non-activating | 0.85 | 0.26 | 5 |
| 149 | 1271 | T1271I | LRR | missense | 8 | VUS |  | Non-activating | 0.51 | 0.17 | 7 |
| 150 | 1295 | W1295R | LRR | missense | 8 | VUS |  | Non-activating | 0.30 | 0.05 | 4 |
| 151 | 1304 | L1304F | LRR | missense | 9 | VUS |  | Non-activating | 0.63 | 0.21 | 3 |
| 152 | 1313 | C1313* | LRR | nonsense | 4 | N/A |  | Non-activating | 0.00 | 0.00 | 3 |
| 153 | 1320 | R1320S | LRR | missense | 9 | VUS | Non-activating | Non-activating | 0.77 | 0.40 | 5 |
| 154 | 1325 | R1325Q | LRR | missense | 9 | Pathogenic | Activating | Activating | 1.49 | 0.77 | 19 |
| 155 | 1330 | V1330M | LRR | missense | 9 | N/A |  | Non-activating | 1.24 | 0.40 | 5 |
| 156 | 1334 | R1334Q | LRR | missense | 9 | VUS |  | Activating | 2.59 | 1.33 | 17 |
| 157 | 1339 | I1339M | ROC | missense | 5 | VUS |  | Non-activating | 1.10 | 0.22 | 4 |
| 158 | 1340 | V1340M | ROC | missense | 9 | VUS |  | Non-activating | 0.66 | 0.13 | 3 |
| 159 | 1347 | K1347R | ROC | missense | 9 | N/A |  | Non-activating | 0.82 | 0.21 | 3 |
| 160 | 1348 | T1348P | ROC | missense | 9 | N/A |  | Non-activating | 0.24 | 0.15 | 3 |
| 161 | 1353 | Q1353K | ROC | missense | 9 | VUS |  | Activating | 1.62 | 0.38 | 12 |
| 162 | 1355 | M1355K | ROC | missense | 7 | N/A |  | Non-activating | 0.95 | 0.23 | 5 |
| 163 | 1366 | S1366N | ROC | missense | 3 | N/A |  | Non-activating | 0.78 | 0.11 | 4 |
| 164 | 1369 | V1369A | ROC | missense | 8 | VUS |  | Non-activating | 1.36 | 0.16 | 4 |
| 165 | 1371 | I1371V | ROC | missense | 9 | Likely benign | Non-activating | Non-activating | 0.79 | 0.41 | 9 |
| 166 | 1373 | V1373M | ROC | missense | 9 | VUS |  | Activating | 2.20 | 0.56 | 17 |
| 167 | 1379 | Q1379R | ROC | missense | 4 | VUS |  | Non-activating | 1.05 | 0.12 | 3 |
| 168 | 1388 | L1388I | ROC | missense | 4 | VUS |  | Activating | 1.65 | 0.99 | 4 |
| 169 | 1389 | V1389I | ROC | missense | 3 | VUS |  | Non-activating | 0.69 | 0.07 | 3 |
| 170 | 1396 | A1396S | ROC | missense | 8 | N/A |  | Non-activating | 1.12 | 0.54 | 2 |
| 171 | 1398 | R1398H | ROC | missense | 4 | N/A |  | Non-activating | 1.17 | 0.32 | 5 |
| 172 | 1402 | Y1402C | ROC | missense | 8 | VUS |  | Activating | 1.78 | 0.40 | 12 |
| 173 | 1405 | H1405Q | ROC | missense | 9 | VUS |  | Non-activating | 1.42 | 0.35 | 11 |
| 174 | 1410 | T1410M | ROC | missense | 8 | N/A |  | Non-activating | 0.93 | 0.15 | 3 |
| 175 | 1413 | A1413T | ROC | missense | 8 | VUS |  | Activating | 2.01 | 0.93 | 17 |
| 176 | 1415 | Y1415E | ROC | missense | 6 | N/A | Non-activating | Non-activating | 0.15 | 0.03 | 3 |
| 177 | 1417 | A1417V | ROC | missense | 8 | N/A | Activating | Activating | 1.58 | 0.34 | 3 |
| 178 | 1417 | A1417E | ROC | missense | 8 | N/A | Non-activating | Non-activating | 0.46 | 0.05 | 3 |
| 179 | 1419 | Y1419C | ROC | missense | 6 | N/A |  | Non-activating | 1.25 | 0.18 | 3 |
| 180 | 1427 | E1427G | ROC | missense | 9 | VUS |  | Non-activating | 1.20 | 0.25 | 6 |
| 181 | 1435 | L1435F | ROC | missense | 9 | N/A | Activating | Activating | 3.48 | 0.44 | 7 |
| 182 | 1435 | L1435E | ROC | missense | 9 | N/A | Non-activating | Non-activating | 0.55 | 0.00 | 3 |
| 183 | 1436 | F1436S | ROC | missense | 9 | N/A |  | Activating | 4.21 | 1.81 | 13 |
| 184 | 1436 | F1436L | ROC | missense | 9 | VUS |  | Activating | 2.47 | 0.80 | 15 |
| 185 | 1437 | N1437H | ROC | missense | 9 | Pathogenic | Activating | Activating | 4.05 | 1.89 | 21 |
| 186 | 1437 | N1437D | ROC | missense | 9 | Likely pathogenic |  | Activating | 3.22 | 1.47 | 10 |
| 187 | 1437 | N1437S | ROC | missense | 9 | Likely pathogenic |  | Non-activating | 0.98 | 0.15 | 5 |
| 188 | 1438 | I1438E | ROC | missense | 8 | N/A | Activating | Activating | 3.78 | 1.10 | 3 |
| 189 | 1438 | I1438T | ROC | missense | 8 | N/A | Activating | Activating | 2.40 | 0.33 | 3 |
| 190 | 1438 | I1438V | ROC | missense | 8 | N/A | Non-activating | Non-activating | 1.04 | 0.13 | 3 |
| 191 | 1439 | K1439R | ROC | missense | 8 | N/A |  | Non-activating | 0.74 | 0.31 | 2 |
| 192 | 1440 | A1440P | ROC | missense | 9 | VUS |  | Activating | 2.89 | 0.98 | 16 |
| 193 | 1441 | R1441G | ROC | missense | 9 | Pathogenic | Activating | Activating | 3.29 | 1.07 | 31 |
| 194 | 1441 | R1441C | ROC | missense | 9 | Pathogenic | Activating | Activating | 2.94 | 1.11 | 23 |
| 195 | 1441 | R1441S | ROC | missense | 9 | Pathogenic | Activating | Activating | 2.89 | 0.91 | 20 |
| 196 | 1441 | R1441H | ROC | missense | 9 | Pathogenic | Activating | Activating | 2.32 | 1.62 | 19 |
| 197 | 1442 | A1442P | ROC | missense | 9 | Pathogenic | Activating | Activating | 2.58 | 1.04 | 19 |
| 198 | 1442 | A1442S | ROC | missense | 9 | N/A |  | Non-activating | 0.66 | #DIV/0! | 1 |
| 199 | 1442 | A1442T | ROC | missense | 9 | N/A |  | Non-activating | 0.67 | 0.69 | 2 |
| 200 | 1443 | S1443A | ROC | missense | 8 | N/A |  | Non-activating | 0.65 | 0.33 | 2 |
| 201 | 1443 | S1443F | ROC | missense | 8 | N/A |  | Non-activating | 0.21 | #DIV/0! | 1 |
| 202 | 1445 | S1445F | ROC | missense | 9 | VUS |  | Non-activating | 0.53 | 0.14 | 3 |
| 203 | 1445 | S1445C | ROC | missense | 9 | VUS |  | Non-activating | 0.53 | 0.04 | 3 |
| 204 | 1446 | P1446L | ROC | missense | 9 | VUS |  | Non-activating | 0.27 | 0.07 | 5 |
| 205 | 1447 | V1447L | ROC | missense | 9 | N/A | Activating | Activating | 3.28 | 0.79 | 16 |
| 206 | 1447 | V1447M | ROC | missense | 9 | Pathogenic | Activating | Activating | 2.74 | 0.83 | 22 |
| 207 | 1447 | V1447G | ROC | missense | 9 | N/A | Activating | Activating | 1.54 | 1.26 | 6 |
| 208 | 1447 | V1447E | ROC | missense | 9 | N/A | Non-activating | Non-activating | 0.20 | 0.07 | 3 |
| 209 | 1450 | V1450I | ROC | missense | 8 | VUS |  | Non-activating | 0.72 | 0.54 | 3 |
| 210 | 1451 | G1451S | ROC | missense | 9 | VUS |  | Non-activating | 0.31 | 0.07 | 6 |
| 211 | 1462 | R1462H | ROC | missense | 8 | N/A |  | Non-activating | 0.82 | 0.24 | 5 |
| 212 | 1464 | A1464G | ROC | missense | 7 | VUS | Non-activating | Non-activating | 1.06 | 0.30 | 9 |
| 213 | 1468 | K1468E | ROC | missense | 8 | VUS | Non-activating | Non-activating | 1.27 | 0.46 | 5 |
| 214 | 1480 | P1480L | ROC | missense | 9 | VUS | Non-activating | Non-activating | 1.04 | 0.38 | 5 |
| 215 | 1481 | A1481T | ROC | missense | 6 | VUS |  | Non-activating | 1.03 | 0.14 | 3 |
| 216 | 1483 | R1483Q | ROC | missense | 9 | VUS |  | Non-activating | 1.12 | 0.17 | 4 |
| 217 | 1490 | A1490V | ROC | missense | 9 | VUS |  | Non-activating | 1.07 | 0.34 | 3 |
| 218 | 1492 | E1492K | ROC | missense | 8 | VUS |  | Activating | 2.14 | 0.88 | 12 |
| 219 | 1501 | R1501W | ROC | missense | 9 | VUS |  | Non-activating | 1.25 | 0.16 | 3 |
| 220 | 1508 | S1508G | ROC | missense | 9 | VUS | Non-activating | Non-activating | 0.85 | 0.23 | 5 |
| 221 | 1508 | S1508R | ROC | missense | 9 | N/A |  | Non-activating | 0.23 | 0.13 | 8 |
| 222 | 1514 | R1514Q | COR-A | missense | 9 | N/A |  | Non-activating | 0.95 | 0.54 | 5 |
| 223 | 1514 | R1514G | COR-A | missense | 9 | N/A |  | Non-activating | 0.88 | #DIV/0! | 1 |
| 224 | 1520 | G1520A | COR-A | missense | 9 | VUS |  | Non-activating | 0.43 | 0.10 | 4 |
| 225 | 1538 | R1538H | COR-A | missense | 9 | N/A |  | Non-activating | 0.91 | 0.29 | 6 |
| 226 | 1538 | R1538C | COR-A | missense | 9 | N/A |  | Non-activating | 0.79 | 0.28 | 3 |
| 227 | 1541 | V1541M | COR-A | missense | 7 | VUS |  | Non-activating | 0.77 | 0.20 | 3 |
| 228 | 1542 | P1542S | COR-A | missense | 9 | N/A |  | Non-activating | 0.93 | 0.30 | 5 |
| 229 | 1548 | I1548V | COR-A | missense | 8 | VUS |  | Non-activating | 1.02 | 0.22 | 4 |
| 230 | 1550 | R1550Q | COR-A | missense | 3 | N/A |  | Non-activating | 0.56 | 0.47 | 2 |
| 231 | 1569 | L1569I | COR-A | missense | 9 | N/A |  | Non-activating | 0.91 | 0.13 | 3 |
| 232 | 1573 | V1573A | COR-A | missense | 5 | N/A |  | Non-activating | 0.74 | 0.16 | 4 |
| 233 | 1589 | A1589S | COR-A | missense | 9 | VUS | Non-activating | Non-activating | 0.16 | 0.11 | 7 |
| 234 | 1613 | V1613A | COR-A | missense | 8 | VUS | Non-activating | Non-activating | 0.66 | 0.09 | 5 |
| 235 | 1615 | V1615M | COR-A | missense | 8 | VUS |  | Non-activating | 1.00 | 0.17 | 3 |
| 236 | 1616 | E1616K | COR-A | missense | 8 | VUS |  | Non-activating | 0.78 | 0.05 | 3 |
| 237 | 1619 | P1619L | COR-A | missense | 3 | VUS |  | Non-activating | 0.91 | 0.12 | 3 |
| 238 | 1620 | K1620R | COR-A | missense | 8 | VUS |  | Non-activating | 1.04 | 0.37 | 5 |
| 239 | 1621 | H1621R | COR-A | missense | 4 | VUS |  | Non-activating | 1.19 | 0.36 | 5 |
| 240 | 1621 | H1621Q | COR-A | missense | 4 | VUS |  | Non-activating | 1.14 | 0.22 | 5 |
| 241 | 1621 | H1621L | COR-A | missense | 4 | N/A |  | Non-activating | 0.77 | 0.29 | 3 |
| 242 | 1622 | P1622L | COR-A | missense | 9 | VUS |  | Activating | 1.72 | 0.54 | 17 |
| 243 | 1622 | P1622T | COR-A | missense | 9 | VUS |  | Non-activating | 1.44 | 0.46 | 18 |
| 244 | 1623 | K1623E | COR-A | missense | 6 | VUS |  | Non-activating | 1.10 | 0.34 | 5 |
| 245 | 1626 | I1626V | COR-A | missense | 9 | VUS |  | Non-activating | 1.07 | 0.37 | 3 |
| 246 | 1627 | S1627T | COR-A | missense | 5 | VUS |  | Non-activating | 1.13 | 0.20 | 3 |
| 247 | 1628 | R1628P | COR-A | missense | 9 | N/A |  | Non-activating | 1.28 | 0.79 | 18 |
| 248 | 1628 | R1628C | COR-A | missense | 9 | VUS | Non-activating | Non-activating | 0.87 | 0.29 | 5 |
| 249 | 1628 | R1628H | COR-A | missense | 9 | N/A |  | Non-activating | 0.71 | 0.30 | 6 |
| 250 | 1645 | Y1645S | COR-A | missense | 8 | VUS |  | Non-activating | 1.38 | 0.64 | 12 |
| 251 | 1646 | M1646T | COR-A | missense | 3 | N/A |  | Non-activating | 1.00 | 0.58 | 5 |
| 252 | 1648 | Q1648R | COR-A | missense | 9 | VUS |  | Non-activating | 0.87 | 0.28 | 8 |
| 253 | 1649 | Y1649S | COR-A | missense | 9 | VUS |  | Non-activating | 0.73 | 0.33 | 3 |
| 254 | 1654 | E1654C | COR-A | missense | 9 | N/A |  | Non-activating | 0.96 | 0.44 | 3 |
| 255 | 1657 | Q1657* | COR-A | nonsense | 9 | N/A |  | Non-activating | 0.00 | 0.00 | 3 |
| 256 | 1661 | P1661T | COR-A | missense | 9 | VUS |  | Non-activating | 0.73 | 0.34 | 3 |
| 257 | 1677 | R1677S | COR-B | missense | 8 | VUS | Non-activating | Non-activating | 0.91 | 0.28 | 5 |
| 258 | 1691 | I1691T | COR-B | missense | 9 | VUS |  | Non-activating | 1.23 | 0.23 | 6 |
| 259 | 1693 | R1693Q | COR-B | missense | 9 | VUS |  | Non-activating | 0.16 | 0.15 | 6 |
| 260 | 1698 | P1698L | COR-B | missense | 9 | N/A |  | Non-activating | 0.50 | 0.56 | 2 |
| 261 | 1699 | Y1699C | COR-B | missense | 7 | Pathogenic | Activating | Activating | 5.55 | 1.92 | 23 |
| 262 | 1700 | F1700L | COR-B | missense | 9 | Pathogenic | Activating | Activating | 4.45 | 1.61 | 17 |
| 263 | 1702 | M1702T | COR-B | missense | 7 | N/A |  | Activating | 1.98 | 1.41 | 17 |
| 264 | 1702 | M1702V | COR-B | missense | 7 | N/A |  | Non-activating | 0.70 | 0.25 | 2 |
| 265 | 1707 | R1707K | COR-B | missense | 9 | VUS |  | Non-activating | 0.46 | 0.22 | 5 |
| 266 | 1712 | L1712F | COR-B | missense | 7 | VUS |  | Activating | 3.31 | 0.89 | 13 |
| 267 | 1725 | R1725Q | COR-B | missense | 5 | VUS | Non-activating | Non-activating | 0.83 | 0.38 | 6 |
| 268 | 1728 | R1728L | COR-B | missense | 9 | N/A |  | Activating | 3.22 | 1.46 | 19 |
| 269 | 1728 | R1728H | COR-B | missense | 9 | Likely pathogenic | Activating | Activating | 2.65 | 0.98 | 19 |
| 270 | 1752 | S1752C | COR-B | missense | 8 | VUS |  | Non-activating | 0.74 | 0.23 | 6 |
| 271 | 1756 | D1756G | COR-B | missense | 6 | VUS |  | Non-activating | 1.07 | 0.10 | 3 |
| 272 | 1756 | D1756Y | COR-B | missense | 6 | VUS |  | Non-activating | 0.79 | 0.11 | 3 |
| 273 | 1758 | H1758P | COR-B | missense | 5 | VUS |  | Non-activating | 0.66 | 0.04 | 3 |
| 274 | 1759 | P1759L | COR-B | missense | 2 | N/A |  | Non-activating | 0.59 | 0.32 | 2 |
| 275 | 1761 | S1761R | COR-B | missense | 9 | Likely pathogenic | Activating | Activating | 3.35 | 1.37 | 19 |
| 276 | 1761 | S1761N | COR-B | missense | 9 | N/A |  | Non-activating | 0.52 | #DIV/0! | 1 |
| 277 | 1762 | F1762S | COR-B | missense | 5 | N/A |  | Non-activating | 1.06 | 0.84 | 2 |
| 278 | 1771 | R1771T | COR-B | missense | 9 | VUS |  | Non-activating | 0.85 | 0.23 | 3 |
| 279 | 1774 | C1774Y | COR-B | missense | 7 | VUS |  | Non-activating | 0.98 | 0.20 | 3 |
| 280 | 1795 | L1795F | COR-B | missense | 9 | Pathogenic | Activating | Activating | 4.51 | 1.50 | 19 |
| 281 | 1797 | E1797D | COR-B | missense | 8 | N/A |  | Non-activating | 0.58 | #DIV/0! | 1 |
| 282 | 1806 | T1806I | COR-B | missense | 9 | VUS |  | Non-activating | 0.50 | 0.40 | 2 |
| 283 | 1819 | G1819R | COR-B | missense | 7 | VUS |  | Non-activating | 0.90 | 0.20 | 3 |
| 284 | 1823 | Q1823H | COR-B | missense | 4 | N/A |  | Non-activating | 1.16 | 0.56 | 5 |
| 285 | 1823 | Q1823K | COR-B | missense | 4 | VUS | Non-activating | Non-activating | 0.81 | 0.16 | 5 |
| 286 | 1827 | L1827F | COR-B | missense | 8 | N/A |  | Non-activating | 0.32 | 0.15 | 3 |
| 287 | 1852 | I1852V | COR-B | missense | 9 | N/A |  | Non-activating | 0.63 | 0.26 | 3 |
| 288 | 1865 | P1865R | COR-B | missense | 9 | N/A |  | Non-activating | 1.08 | 0.52 | 5 |
| 289 | 1869 | M1869V | COR-B | missense | 6 | VUS |  | Non-activating | 1.33 | 0.56 | 5 |
| 290 | 1869 | M1869T | COR-B | missense | 6 | VUS | Non-activating | Non-activating | 0.45 | 0.16 | 6 |
| 291 | 1887 | D1887N | KIN | missense | 7 | VUS |  | Activating | 1.84 | 0.99 | 15 |
| 292 | 1891 | G1891A | KIN | missense | 8 | VUS |  | Activating | 2.43 | 0.74 | 15 |
| 293 | 1900 | G1900E | KIN | missense | 9 | N/A |  | Activating | 2.22 | 1.07 | 16 |
| 294 | 1914 | L1914I | KIN | missense | 5 | VUS |  | Activating | 1.96 | 0.84 | 17 |
| 295 | 1941 | R1941H | KIN | missense | 4 | VUS | Non-activating | Non-activating | 0.21 | 0.24 | 5 |
| 296 | 1954 | S1954F | KIN | missense | 9 | Likely pathogenic |  | Non-activating | 0.26 | 0.14 | 3 |
| 297 | 1957 | R1957G | KIN | missense | 7 | N/A |  | Non-activating | 0.82 | 0.31 | 3 |
| 298 | 1961 | Q1961R | KIN | missense | 5 | VUS |  | Non-activating | 0.72 | 0.28 | 6 |
| 299 | 1991 | I1991V | KIN | missense | 8 | VUS | Non-activating | Non-activating | 0.53 | 0.21 | 6 |
| 300 | 2006 | Y2006H | KIN | missense | 6 | VUS | Non-activating | Non-activating | 1.13 | 0.31 | 5 |
| 301 | 2010 | A2010T | KIN | missense | 7 | VUS | Non-activating | Non-activating | 0.70 | 0.30 | 5 |
| 302 | 2012 | I2012T | KIN | missense | 8 | VUS | Non-activating | Non-activating | 0.77 | 0.36 | 6 |
| 303 | 2019 | G2019S | KIN | missense | 8 | Pathogenic | Activating | Activating | 1.44 | 0.45 | 25 |
| 304 | 2020 | I2020T | KIN | missense | 9 | Pathogenic | Activating | Activating | 3.43 | 0.86 | 20 |
| 305 | 2020 | I2020S | KIN | missense | 9 | Likely pathogenic |  | Activating | 3.15 | 1.50 | 17 |
| 306 | 2020 | I2020L | KIN | missense | 9 | Likely pathogenic |  | Non-activating | 0.64 | 0.33 | 3 |
| 307 | 2031 | T2031S | KIN | missense | 9 | Likely pathogenic | Activating | Activating | 1.54 | 0.63 | 19 |
| 308 | 2063 | L2063* | KIN | nonsense | 9 | VUS |  | Non-activating | 0.00 | 0.00 | 3 |
| 309 | 2071 | G2071D | KIN | missense | 1 | N/A |  | Non-activating | 1.02 | 0.14 | 3 |
| 310 | 2074 | V2074I | KIN | missense | 3 | VUS |  | Non-activating | 1.26 | 0.17 | 3 |
| 311 | 2081 | N2081D | KIN | missense | 8 | N/A |  | Activating | 1.77 | 0.53 | 19 |
| 312 | 2084 | D2084N | KIN | missense | 9 | N/A |  | Non-activating | 0.92 | 0.34 | 5 |
| 313 | 2108 | E2108K | KIN | missense | 8 | VUS |  | Non-activating | 0.30 | 0.14 | 6 |
| 314 | 2133 | N2133S | KIN | missense | 9 | VUS |  | Non-activating | 0.88 | 0.32 | 3 |
| 315 | 2139 | C2139S | KIN | missense | 9 | VUS |  | Non-activating | 0.71 | 0.15 | 4 |
| 316 | 2141 | T2141M | KIN | missense | 6 | N/A |  | Non-activating | 0.53 | 0.45 | 5 |
| 317 | 2143 | R2143H | WD40 | missense | 2 | N/A |  | Non-activating | 0.95 | 0.57 | 6 |
| 318 | 2143 | R2143M | WD40 | missense | 2 | N/A |  | Non-activating | 0.65 | 0.25 | 5 |
| 319 | 2150 | V2150I | WD40 | missense | 1 | VUS |  | Non-activating | 1.10 | 0.14 | 3 |
| 320 | 2168 | W2168L | WD40 | missense | 4 | VUS |  | Non-activating | 0.76 | 0.44 | 3 |
| 321 | 2175 | D2175H | WD40 | missense | 4 | VUS | Non-activating | Non-activating | 0.86 | 0.22 | 6 |
| 322 | 2189 | Y2189C | WD40 | missense | 2 | VUS | Non-activating | Non-activating | 0.81 | 0.29 | 5 |
| 323 | 2213 | S2213C | WD40 | missense | 5 | N/A |  | Non-activating | 1.02 | 0.24 | 3 |
| 324 | 2236 | H2236R | WD40 | missense | 9 | VUS |  | Non-activating | 0.20 | 0.05 | 6 |
| 325 | 2251 | N2251T | WD40 | missense | 3 | VUS |  | Non-activating | 1.31 | 0.35 | 3 |
| 326 | 2255 | K2255M | WD40 | missense | 6 | N/A |  | Non-activating | 1.16 | 0.47 | 2 |
| 327 | 2261 | N2261K | WD40 | missense | 3 | VUS |  | Non-activating | 0.97 | 0.33 | 3 |
| 328 | 2278 | K2278E | WD40 | missense | 4 | N/A |  | Non-activating | 0.98 | 0.23 | 4 |
| 329 | 2294 | G2294R | WD40 | missense | 9 | VUS |  | Non-activating | 0.40 | 0.10 | 6 |
| 330 | 2296 | V2296F | WD40 | missense | 3 | N/A |  | Non-activating | 0.98 | 0.30 | 2 |
| 331 | 2308 | N2308D | WD40 | missense | 4 | VUS | Non-activating | Non-activating | 1.09 | 0.45 | 5 |
| 332 | 2310 | T2310A | WD40 | missense | 1 | N/A |  | Non-activating | 1.06 | 0.31 | 5 |
| 333 | 2310 | T2310M | WD40 | missense | 1 | VUS |  | Non-activating | 0.94 | 0.14 | 5 |
| 334 | 2313 | N2313S | WD40 | missense | 4 | VUS | Non-activating | Non-activating | 0.85 | 0.44 | 5 |
| 335 | 2323 | I2323F | WD40 | missense | 3 | VUS |  | Non-activating | 0.74 | 0.16 | 3 |
| 336 | 2336 | I2336V | WD40 | missense | 9 | VUS |  | Non-activating | 0.91 | 0.26 | 3 |
| 337 | 2350 | S2350I | WD40 | missense | 5 | VUS | Non-activating | Non-activating | 0.98 | 0.33 | 5 |
| 338 | 2356 | T2356I | WD40 | missense | 6 | VUS | Non-activating | Non-activating | 1.09 | 0.48 | 6 |
| 339 | 2385 | G2385R | WD40 | missense | 5 | N/A |  | Activating | 1.92 | 0.57 | 25 |
| 340 | 2390 | V2390A | WD40 | missense | 5 | VUS |  | Non-activating | 1.20 | 0.50 | 3 |
| 341 | 2390 | V2390M | WD40 | missense | 5 | VUS | Non-activating | Non-activating | 0.35 | 0.12 | 5 |
| 342 | 2391 | H2391Q | WD40 | missense | 5 | Likely benign |  | Non-activating | 1.18 | 0.31 | 5 |
| 343 | 2395 | E2395K | WD40 | missense | 6 | N/A |  | Non-activating | 0.87 | 0.15 | 3 |
| 344 | 2397 | M2397T | WD40 | missense | 2 | N/A |  | Non-activating | 1.05 | 0.29 | 10 |
| 345 | 2423 | T2423S | WD40 | missense | 7 | VUS |  | Non-activating | 0.57 | 0.05 | 3 |
| 346 | 2434 | I2434V | WD40 | missense | 4 | VUS |  | Non-activating | 0.81 | 0.18 | 3 |
| 347 | 2436 | L2436P | WD40 | missense | 6 | N/A |  | Non-activating | 0.12 | 0.04 | 4 |
| 348 | 2439 | L2439I | WD40 | missense | 6 | VUS | Non-activating | Non-activating | 0.70 | 0.16 | 5 |
| 349 | 2461 | A2461V | WD40 | missense | 5 | VUS |  | Non-activating | 0.92 | 0.19 | 3 |
| 350 | 2463 | L2463P | WD40 | missense | 5 | VUS |  | Non-activating | 1.00 | 0.16 | 3 |
| 351 | 2466 | L2466H | WD40 | missense | 6 | N/A |  | Non-activating | 0.72 | 0.24 | 5 |
| 352 | 2467 | K2467Q | WD40 | missense | 5 | N/A |  | Activating | 2.21 | 0.84 | 16 |
| 353 | 2484 | Q2484K | WD40 | missense | 3 | N/A |  | Non-activating | 1.03 | 0.14 | 4 |
| 354 | 2490 | Q2490H | WD40 | missense | 5 | VUS |  | Non-activating | 0.80 | 0.23 | 3 |
| 355 | 2494 | T2494I | WD40 | missense | 7 | VUS | Non-activating | Non-activating | 0.44 | 0.21 | 5 |
| 356 | 2495 | V2495I | WD40 | missense | 3 | VUS |  | Non-activating | 0.95 | 0.39 | 4 |
| 357 | 2524 | T2524A | WD40 | missense | 2 | VUS |  | Non-activating | 1.11 | 0.16 | 3 |
| 358 | 1628 +2385 | R1628P-G2385R |  | double mutation |  | N/A |  | Activating | 2.35 | 0.48 | 10 |
| 359 | 445+1441 | S455P-R1441H |  | double mutation |  | N/A |  | Activating | 4.49 | 0.96 | 8 |

Pos., residue position; MDS ACMG, MDSGene ACMG classification; SD, standard deviation; Repl. (n), number of independent measurements. Functional outcomes were classified as activating or non-activating. Mean pRAB10/RAB10 values were calculated across all available studies performed under comparable experimental conditions; for variants assessed in a single study, the reported value corresponds to that study. Abbreviations: ACMG, American College of Medical Genetics and Genomics; VUS, variant of uncertain significance.

**Supplementary Table 4: List of plasmids**

| **DU Number** | **Construct** | **Plasmid** |
| --- | --- | --- |
| DU41799 | FLAG-empty | pCMV5D |
| DU6841 | FLAG LRRK2 wildtype | pCMV5 |
| DU10128 | FLAG LRRK2 D2017A (KD) | pCMV5 |
| DU13826 | FLAG LRRK2 E10K | pCMV5 |
| DU80689 | FLAG LRRK2 K53R | pCMV5 |
| DU77979 | FLAG LRRK2 V80M | pCMV5 |
| DU77980 | FLAG LRRK2 M96T | pCMV5 |
| DU62019 | FLAG LRRK2 M100T | pCMV5 |
| DU72415 | FLAG LRRK2 D104Y | pCMV5 |
| DU68340 | FLAG LRRK2 H115P | pCMV5 |
| DU77849 | FLAG LRRK2 Q116R | pCMV5 |
| DU68326 | FLAG LRRK2 L119P | pCMV5 |
| DU80443 | FLAG LRRK2 S128G | pCMV5 |
| DU77870 | FLAG LRRK2 T149S | pCMV5 |
| DU62020 | FLAG LRRK2 L153W | pCMV5 |
| DU77782 | FLAG LRRK2 F160L | pCMV5 |
| DU80444 | FLAG LRRK2 N173S | pCMV5 |
| DU80738 | FLAG LRRK2 V176L | pCMV5 |
| DU72416 | FLAG LRRK2 S192A | pCMV5 |
| DU62183 | FLAG LRRK2 E193K | pCMV5 |
| DU80188 | FLAG LRRK2 E193A | pCMV5 |
| DU26913 | FLAG LRRK2 A211V | pCMV5 |
| DU80435 | FLAG LRRK2 C228S | pCMV5 |
| DU61842 | FLAG LRRK2 H230R | pCMV5 |
| DU72417 | FLAG LRRK2 S231P | pCMV5 |
| DU72361 | FLAG LRRK2 Y250C | pCMV5 |
| DU72077 | FLAG LRRK2 E255K | pCMV5 |
| DU80436 | FLAG LRRK2 K258N | pCMV5 |
| DU80690 | FLAG LRRK2 A259E | pCMV5 |
| DU68327 | FLAG LRRK2 M262V | pCMV5 |
| DU80165 | FLAG LRRK2 L286V | pCMV5 |
| DU80166 | FLAG LRRK2 V291A | pCMV5 |
| DU77830 | FLAG LRRK2 H292Q | pCMV5 |
| DU77449 | FLAG LRRK2 A306V | pCMV5 |
| DU72418 | FLAG LRRK2 L308M | pCMV5 |
| DU80176 | FLAG LRRK2 A312V | pCMV5 |
| DU77866 | FLAG LRRK2 L313F | pCMV5 |
| DU80175 | FLAG LRRK2 T320S | pCMV5 |
| DU26914 | FLAG LRRK2 E334K | pCMV5 |
| DU80437 | FLAG LRRK2 N335S | pCMV5 |
| DU77719 | FLAG LRRK2 C353Y | pCMV5 |
| DU77873 | FLAG LRRK2 A356T | pCMV5 |
| DU26911 | FLAG LRRK2 N363S | pCMV5 |
| DU68969 | FLAG LRRK2 H365N | pCMV5 |
| DU62006 | FLAG LRRK2 A370T | pCMV5 |
| DU80438 | FLAG LRRK2 L378F | pCMV5 |
| DU80439 | FLAG LRRK2 M379I | pCMV5 |
| DU72045 | FLAG LRRK2 H385R | pCMV5 |
| DU68328 | FLAG LRRK2 I388T | pCMV5 |
| DU72361 | FLAG LRRK2 Y250C | pCMV5 |
| DU72077 | FLAG LRRK2 E255K | pCMV5 |
| DU80436 | FLAG LRRK2 K258N | pCMV5 |
| DU80690 | FLAG LRRK2 A259E | pCMV5 |
| DU68327 | FLAG LRRK2 M262V | pCMV5 |
| DU80165 | FLAG LRRK2 L286V | pCMV5 |
| DU80166 | FLAG LRRK2 V291A | pCMV5 |
| DU77830 | FLAG LRRK2 H292Q | pCMV5 |
| DU77449 | FLAG LRRK2 A306V | pCMV5 |
| DU72418 | FLAG LRRK2 L308M | pCMV5 |
| DU80176 | FLAG LRRK2 A312V | pCMV5 |
| DU77866 | FLAG LRRK2 L313F | pCMV5 |
| DU80175 | FLAG LRRK2 T320S | pCMV5 |
| DU26914 | FLAG LRRK2 E334K | pCMV5 |
| DU80437 | FLAG LRRK2 N335S | pCMV5 |
| DU77719 | FLAG LRRK2 C353Y | pCMV5 |
| DU77873 | FLAG LRRK2 A356T | pCMV5 |
| DU26911 | FLAG LRRK2 N363S | pCMV5 |
| DU68969 | FLAG LRRK2 H365N | pCMV5 |
| DU62006 | FLAG LRRK2 A370T | pCMV5 |
| DU80438 | FLAG LRRK2 L378F | pCMV5 |
| DU80439 | FLAG LRRK2 M379I | pCMV5 |
| DU72045 | FLAG LRRK2 H385R | pCMV5 |
| DU68328 | FLAG LRRK2 I388T | pCMV5 |
| DU27513 | FLAG LRRK2 A397T | pCMV5 |
| DU68566 | FLAG LRRK2 V414I | pCMV5 |
| DU26842 | FLAG LRRK2 A419V | pCMV5 |
| DU80196 | FLAG LRRK2 L425P | pCMV5 |
| DU80189 | FLAG LRRK2 L437S | pCMV5 |
| DU80466 | FLAG LRRK2 S438P | pCMV5 |
| DU77781 | FLAG LRRK2 H442Y | pCMV5 |
| DU77437 | FLAG LRRK2 S455P | pCMV5 |
| DU62596 | FLAG LRRK2 A459S | pCMV5 |
| DU80163 | FLAG LRRK2 S461I | pCMV5 |
| DU80755 | FLAG LRRK2 N474S | pCMV5 |
| DU68330 | FLAG LRRK2 D478Y | pCMV5 |
| DU68331 | FLAG LRRK2 I479V | pCMV5 |
| DU80990 | FLAG LRRK2 L488P | pCMV5 |
| DU77438 | FLAG LRRK2 A504T | pCMV5 |
| DU68845 | FLAG LRRK2 R506P | pCMV5 |
| DU77874 | FLAG LRRK2 R506Q | pCMV5 |
| DU77875 | FLAG LRRK2 E518K | pCMV5 |
| DU80721 | FLAG LRRK2 R521G | pCMV5 |
| DU13049 | FLAG LRRK2 K544E | pCMV5 |
| DU26736 | FLAG LRRK2 N551K | pCMV5 |
| DU68565 | FLAG LRRK2 S581T | pCMV5 |
| DU77727 | FLAG LRRK2 I610T | pCMV5 |
| DU62834 | FLAG LRRK2 K616R | pCMV5 |
| DU80440 | FLAG LRRK2 S633F | pCMV5 |
| DU72355 | FLAG LRRK2 R637Q | pCMV5 |
| DU80445 | FLAG LRRK2 A654V | pCMV5 |
| DU80446 | FLAG LRRK2 S661F | pCMV5 |
| DU80447 | FLAG LRRK2 I675M | pCMV5 |
| DU72419 | FLAG LRRK2 F691S | pCMV5 |
| DU26915 | FLAG LRRK2 M712V | pCMV5 |
| DU62002 | FLAG LRRK2 S722N | pCMV5 |
| DU68332 | FLAG LRRK2 I723V | pCMV5 |
| DU80573 | FLAG LRRK2 M724V | pCMV5 |
| DU68709 | FLAG LRRK2 D734N | pCMV5 |
| DU80448 | FLAG LRRK2 K739R | pCMV5 |
| DU26926 | FLAG LRRK2 P755L | pCMV5 |
| DU72384 | FLAG LRRK2 L757V | pCMV5 |
| DU80441 | FLAG LRRK2 S764N | pCMV5 |
| DU26708 | FLAG LRRK2 R767H | pCMV5 |
| DU72420 | FLAG LRRK2 R767C | pCMV5 |
| DU68333 | FLAG LRRK2 T776M | pCMV5 |
| DU80450 | FLAG LRRK2 I777M | pCMV5 |
| DU77717 | FLAG LRRK2 S784R | pCMV5 |
| DU80451 | FLAG LRRK2 I786F | pCMV5 |
| DU62016 | FLAG LRRK2 R792K | pCMV5 |
| DU26912 | FLAG LRRK2 R793M | pCMV5 |
| DU26907 | FLAG LRRK2 I810V | pCMV5 |
| DU72431 | FLAG LRRK2 K812N | pCMV5 |
| DU72188 | FLAG LRRK2 W817C | pCMV5 |
| DU80691 | FLAG LRRK2 S827F | pCMV5 |
| DU80472 | FLAG LRRK2 I844N | pCMV5 |
| DU80692 | FLAG LRRK2 M848T | pCMV5 |
| DU72354 | FLAG LRRK2 S860R | pCMV5 |
| DU26722 | FLAG LRRK2 S865F | pCMV5 |
| DU72358 | FLAG LRRK2 D867N | pCMV5 |
| DU26709 | FLAG LRRK2 S885N | pCMV5 |
| DU80452 | FLAG LRRK2 S885C | pCMV5 |
| DU72209 | FLAG LRRK2 E899D | pCMV5 |
| DU72460 | FLAG LRRK2 R918Q | pCMV5 |
| DU62855 | FLAG LRRK2 Q923H | pCMV5 |
| DU62012 | FLAG LRRK2 C925Y | pCMV5 |
| DU13164 | FLAG LRRK2 Q930R | pCMV5 |
| DU80291 | FLAG LRRK2 R931K | pCMV5 |
| DU80453 | FLAG LRRK2 D941Y | pCMV5 |
| DU68334 | FLAG LRRK2 D944V | pCMV5 |
| DU80693 | FLAG LRRK2 R948Q | pCMV5 |
| DU72438 | FLAG LRRK2 S954T | pCMV5 |
| DU80722 | FLAG LRRK2 S958L | pCMV5 |
| DU68567 | FLAG LRRK2 R969K | pCMV5 |
| DU80565 | FLAG LRRK2 D972G | pCMV5 |
| DU80454 | FLAG LRRK2 S973G | pCMV5 |
| DU13082 | FLAG LRRK2 S973N | pCMV5 |
| DU62038 | FLAG LRRK2 R981K | pCMV5 |
| DU80290 | FLAG LRRK2 I984V | pCMV5 |
| DU62003 | FLAG LRRK2 S1007T | pCMV5 |
| DU80289 | FLAG LRRK2 K1015R | pCMV5 |
| DU80118 | FLAG LRRK2 H1019R | pCMV5 |
| DU80694 | FLAG LRRK2 Q1029H | pCMV5 |
| DU13043 | FLAG LRRK2 R1067Q | pCMV5 |
| DU80353 | FLAG LRRK2 N1089S | pCMV5 |
| DU13044 | FLAG LRRK2 S1096C | pCMV5 |
| DU72432 | FLAG LRRK2 P1099S | pCMV5 |
| DU13988 | FLAG LRRK2 Q1111H | pCMV5 |
| DU13286 | FLAG LRRK2 I1122V | pCMV5 |
| DU80363 | FLAG LRRK2 L1128M | pCMV5 |
| DU72387 | FLAG LRRK2 K1132E | pCMV5 |
| DU26724 | FLAG LRRK2 K1138E | pCMV5 |
| DU72439 | FLAG LRRK2 I1141V | pCMV5 |
| DU19010 | FLAG LRRK2 A1151T | pCMV5 |
| DU80119 | FLAG LRRK2 C1152F | pCMV5 |
| DU80475 | FLAG LRRK2 S1159R | pCMV5 |
| DUA1160D | FLAG LRRK2 A1160D | pCMV5 |
| DUL1165P | FLAG LRRK2 L1165P | pCMV5 |
| DU80120 | FLAG LRRK2 S1181Y | pCMV5 |
| DU17133 | FLAG LRRK2 I1192V | pCMV5 |
| DU72365 | FLAG LRRK2 I1192M | pCMV5 |
| DU13045 | FLAG LRRK2 A1215T | pCMV5 |
| DU80695 | FLAG LRRK2 H1216R | pCMV5 |
| DU13045 | FLAG LRRK2 S1228T | pCMV5 |
| DU72359 | FLAG LRRK2 P1262A | pCMV5 |
| DU68913 | FLAG LRRK2 T1271I | pCMV5 |
| DU72435 | FLAG LRRK2 W1295R | pCMV5 |
| DU80121 | FLAG LRRK2 L1304F | pCMV5 |
| DU80360 | FLAG LRRK2 C1313* | pCMV5 |
| DU62001 | FLAG LRRK2 R1320S | pCMV5 |
| DU62011 | FLAG LRRK2 R1325Q | pCMV5 |
| DU72190 | FLAG LRRK2 V1330M | pCMV5 |
| DU68926 | FLAG LRRK2 R1334Q | pCMV5 |
| DU72434 | FLAG LRRK2 I1339M | pCMV5 |
| DU80122 | FLAG LRRK2 V1340M | pCMV5 |
| DU80292 | FLAG LRRK2 K1347R | pCMV5 |
| DU72581 | FLAG LRRK2 T1348P | pCMV5 |
| DU80123 | FLAG LRRK2 Q1353K | pCMV5 |
| DU68561 | FLAG LRRK2 M1355K | pCMV5 |
| DU72465 | FLAG LRRK2 S1366N | pCMV5 |
| DU72450 | FLAG LRRK2 V1369A | pCMV5 |
| DU13046 | FLAG LRRK2 I1371V | pCMV5 |
| DU72367 | FLAG LRRK2 V1373M | pCMV5 |
| DU80457 | FLAG LRRK2 Q1379R | pCMV5 |
| DU77850 | FLAG LRRK2 L1388I | pCMV5 |
| DU80471 | FLAG LRRK2 V1389I | pCMV5 |
| DU72974 | FLAG LRRK2 A1396S | pCMV5 |
| DU26565 | FLAG LRRK2 R1398H | pCMV5 |
| DU80132 | FLAG LRRK2 Y1402C | pCMV5 |
| DU80133 | FLAG LRRK2 H1405Q | pCMV5 |
| DU77773 | FLAG LRRK2 T1410M | pCMV5 |
| DU68711 | FLAG LRRK2 A1413T | pCMV5 |
| DU80014 | FLAG LRRK2 Y1415E | pCMV5 |
| DU77993 | FLAG LRRK2 A1417V | pCMV5 |
| DU77994 | FLAG LRRK2 A1417E | pCMV5 |
| DU80729 | FLAG LRRK2 Y1419C | pCMV5 |
| DU80476 | FLAG LRRK2 E1427G | pCMV5 |
| DU77991 | FLAG LRRK2 L1435F | pCMV5 |
| DU77992 | FLAG LRRK2 L1435E | pCMV5 |
| DU77435 | FLAG LRRK2 F1436S | pCMV5 |
| DU80134 | FLAG LRRK2 F1436L | pCMV5 |
| DU26643 | FLAG LRRK2 N1437H | pCMV5 |
| DU77202 | FLAG LRRK2 N1437D | pCMV5 |
| DU61441 | FLAG LRRK2 N1437S | pCMV5 |
| DU77995 | FLAG LRRK2 I1438E | pCMV5 |
| DU77990 | FLAG LRRK2 I1438T | pCMV5 |
| DU80013 | FLAG LRRK2 I1438V | pCMV5 |
| DU77170 | FLAG LRRK2 K1439R | pCMV5 |
| DU61855 | FLAG LRRK2 A1440P | pCMV5 |
| DU26477 | FLAG LRRK2 R1441G | pCMV5 |
| DU13078 | FLAG LRRK2 R1441C | pCMV5 |
| DU62906 | FLAG LRRK2 R1441S | pCMV5 |
| DU13287 | FLAG LRRK2 R1441H | pCMV5 |
| DU13170 | FLAG LRRK2 A1442P | pCMV5 |
| DU77168 | FLAG LRRK2 A1442T | pCMV5 |
| DU77169 | FLAG LRRK2 A1442S | pCMV5 |
| DU48031 | FLAG LRRK2 S1443A | pCMV5 |
| DU77201 | FLAG LRRK2 S1443F | pCMV5 |
| DU80135 | FLAG LRRK2 S1445F | pCMV5 |
| DU80136 | FLAG LRRK2 S1445C | pCMV5 |
| DU72046 | FLAG LRRK2 P1446L | pCMV5 |
| DU77444 | FLAG LRRK2 V1447L | pCMV5 |
| DU62501 | FLAG LRRK2 V1447M | pCMV5 |
| DU77977 | FLAG LRRK2 V1447G | pCMV5 |
| DU77977 | FLAG LRRK2 V1447E | pCMV5 |
| DU72100 | FLAG LRRK2 V1450I | pCMV5 |
| DU80137 | FLAG LRRK2 G1451S | pCMV5 |
| DU68568 | FLAG LRRK2 R1462H | pCMV5 |
| DU62849 | FLAG LRRK2 A1464G | pCMV5 |
| DU68324 | FLAG LRRK2 K1468E | pCMV5 |
| DU68710 | FLAG LRRK2 P1480L | pCMV5 |
| DU80534 | FLAG LRRK2 A1481T | pCMV5 |
| DU72423 | FLAG LRRK2 R1483Q | pCMV5 |
| DU80458 | FLAG LRRK2 A1490V | pCMV5 |
| DU80128 | FLAG LRRK2 E1492K | pCMV5 |
| DU72597 | FLAG LRRK2 R1501W | pCMV5 |
| DU62836 | FLAG LRRK2 S1508G | pCMV5 |
| DU62848 | FLAG LRRK2 S1508R | pCMV5 |
| DU13047 | FLAG LRRK2 R1514Q | pCMV5 |
| DU67599 | FLAG LRRK2 R1514G | pCMV5 |
| DU72424 | FLAG LRRK2 G1520A | pCMV5 |
| DU80330 | FLAG LRRK2 R1538H | pCMV5 |
| DU80301 | FLAG LRRK2 R1538C | pCMV5 |
| DU80140 | FLAG LRRK2 V1541M | pCMV5 |
| DU68335 | FLAG LRRK2 P1542S | pCMV5 |
| DU72425 | FLAG LRRK2 I1548V | pCMV5 |
| DU26730 | FLAG LRRK2 R1550Q | pCMV5 |
| DU72426 | FLAG LRRK2 L1569I | pCMV5 |
| DU72427 | FLAG LRRK2 V1573A | pCMV5 |
| DU68336 | FLAG LRRK2 A1589S | pCMV5 |
| DU19019 | FLAG LRRK2 V1613A | pCMV5 |
| DU80497 | FLAG LRRK2 V1615M | pCMV5 |
| DU80459 | FLAG LRRK2 E1616K | pCMV5 |
| DU80696 | FLAG LRRK2 P1619L | pCMV5 |
| DU68636 | FLAG LRRK2 K1620R | pCMV5 |
| DU68629 | FLAG LRRK2 H1621R | pCMV5 |
| DU68637 | FLAG LRRK2 H1621Q | pCMV5 |
| DU77776 | FLAG LRRK2 H1621L | pCMV5 |
| DU68639 | FLAG LRRK2 P1622L | pCMV5 |
| DU68638 | FLAG LRRK2 P1622T | pCMV5 |
| DU68642 | FLAG LRRK2 K1623E | pCMV5 |
| DU80469 | FLAG LRRK2 I1626V | pCMV5 |
| DU80460 | FLAG LRRK2 S1627T | pCMV5 |
| DU19007 | FLAG LRRK2 R1628P | pCMV5 |
| DU68325 | FLAG LRRK2 R1628C | pCMV5 |
| DU72440 | FLAG LRRK2 R1628H | pCMV5 |
| DU80126 | FLAG LRRK2 Y1645S | pCMV5 |
| DU26840 | FLAG LRRK2 M1646T | pCMV5 |
| DU72360 | FLAG LRRK2 Q1648R | pCMV5 |
| DU80141 | FLAG LRRK2 Y1649S | pCMV5 |
| DU77851 | FLAG LRRK2 E1654C | pCMV5 |
| DU76258 | FLAG LRRK2 Q1657* | pCMV5 |
| DU80142 | FLAG LRRK2 P1661T | pCMV5 |
| DU62517 | FLAG LRRK2 R1677S | pCMV5 |
| DU80124 | FLAG LRRK2 I1691T | pCMV5 |
| DU80143 | FLAG LRRK2 R1693Q | pCMV5 |
| DU77162 | FLAG LRRK2 P1698L | pCMV5 |
| DU26486 | FLAG LRRK2 Y1699C | pCMV5 |
| DU72356 | FLAG LRRK2 F1700L | pCMV5 |
| DU61454 | FLAG LRRK2 M1702T | pCMV5 |
| DU77163 | FLAG LRRK2 M1702V | pCMV5 |
| DU68719 | FLAG LRRK2 R1707K | pCMV5 |
| DU77436 | FLAG LRRK2 L1712F | pCMV5 |
| DU62840 | FLAG LRRK2 R1725Q | pCMV5 |
| DU17139 | FLAG LRRK2 R1728L | pCMV5 |
| DU17138 | FLAG LRRK2 R1728H | pCMV5 |
| DU80413 | FLAG LRRK2 S1752C | pCMV5 |
| DU80467 | FLAG LRRK2 D1756G | pCMV5 |
| DU80156 | FLAG LRRK2 D1756Y | pCMV5 |
| DU80455 | FLAG LRRK2 H1758P | pCMV5 |
| DU77164 | FLAG LRRK2 P1759L | pCMV5 |
| DU62839 | FLAG LRRK2 S1761R | pCMV5 |
| DU77165 | FLAG LRRK2 S1761N | pCMV5 |
| DU77166 | FLAG LRRK2 F1762S | pCMV5 |
| DU80456 | FLAG LRRK2 R1771T | pCMV5 |
| DU80468 | FLAG LRRK2 C1774Y | pCMV5 |
| DU17134 | FLAG LRRK2 L1795F | pCMV5 |
| DU77167 | FLAG LRRK2 E1797D | pCMV5 |
| DU72428 | FLAG LRRK2 T1806I | pCMV5 |
| DU80127 | FLAG LRRK2 G1819R | pCMV5 |
| DU72364 | FLAG LRRK2 Q1823H | pCMV5 |
| DU62838 | FLAG LRRK2 Q1823K | pCMV5 |
| DU80304 | FLAG LRRK2 L1827F | pCMV5 |
| DU77911 | FLAG LRRK2 I1852V | pCMV5 |
| DU72189 | FLAG LRRK2 P1865R | pCMV5 |
| DU72363 | FLAG LRRK2 M1869V | pCMV5 |
| DU13169 | FLAG LRRK2 M1869T | pCMV5 |
| DU80144 | FLAG LRRK2 D1887N | pCMV5 |
| DU80145 | FLAG LRRK2 G1891A | pCMV5 |
| DU72187 | FLAG LRRK2 G1900E | pCMV5 |
| DU72366 | FLAG LRRK2 L1914I | pCMV5 |
| DU13079 | FLAG LRRK2 R1941H | pCMV5 |
| DU77852 | FLAG LRRK2 S1954F | pCMV5 |
| DU77725 | FLAG LRRK2 R1957G | pCMV5 |
| DU80463 | FLAG LRRK2 Q1961R | pCMV5 |
| DU62837 | FLAG LRRK2 I1991V | pCMV5 |
| DU13880 | FLAG LRRK2 Y2006H | pCMV5 |
| DU72368 | FLAG LRRK2 A2010T | pCMV5 |
| DU13080 | FLAG LRRK2 I2012T | pCMV5 |
| DU10129 | FLAG LRRK2 G2019S | pCMV5 |
| DU13081 | FLAG LRRK2 I2020T | pCMV5 |
| DU72362 | FLAG LRRK2 I2020S | pCMV5 |
| DU44952 | FLAG LRRK2 I2020L | pCMV5 |
| DU17135 | FLAG LRRK2 T2031S | pCMV5 |
| DU80365 | FLAG LRRK2 L2063* | pCMV5 |
| DU77780 | FLAG LRRK2 G2071D | pCMV5 |
| DU80464 | FLAG LRRK2 V2074I | pCMV5 |
| DU26721 | FLAG LRRK2 N2081D | pCMV5 |
| DU72353 | FLAG LRRK2 D2084N | pCMV5 |
| DU80151 | FLAG LRRK2 E2108K | pCMV5 |
| DU80152 | FLAG LRRK2 N2133S | pCMV5 |
| DU72414 | FLAG LRRK2 C2139S | pCMV5 |
| DU17140 | FLAG LRRK2 T2141M | pCMV5 |
| DU72441 | FLAG LRRK2 R2143H | pCMV5 |
| DU17141 | FLAG LRRK2 R2143M | pCMV5 |
| DU80697 | FLAG LRRK2 V2150I | pCMV5 |
| DU80153 | FLAG LRRK2 W2168L | pCMV5 |
| DU62374 | FLAG LRRK2 D2175H | pCMV5 |
| DU62391 | FLAG LRRK2 Y2189C | pCMV5 |
| DU77726 | FLAG LRRK2 S2213C | pCMV5 |
| DU80154 | FLAG LRRK2 H2236R | pCMV5 |
| DU80473 | FLAG LRRK2 N2251T | pCMV5 |
| DU72451 | FLAG LRRK2 K2255M | pCMV5 |
| DU80489 | FLAG LRRK2 N2261K | pCMV5 |
| DU72386 | FLAG LRRK2 K2278E | pCMV5 |
| DU80158 | FLAG LRRK2 G2294R | pCMV5 |
| DU72453 | FLAG LRRK2 V2296F | pCMV5 |
| DU62004 | FLAG LRRK2 N2308D | pCMV5 |
| DU61451 | FLAG LRRK2 T2310A | pCMV5 |
| DU72357 | FLAG LRRK2 T2310M | pCMV5 |
| DU62005 | FLAG LRRK2 N2313S | pCMV5 |
| DU80465 | FLAG LRRK2 I2323F | pCMV5 |
| DU80155 | FLAG LRRK2 I2336V | pCMV5 |
| DU62502 | FLAG LRRK2 S2350I | pCMV5 |
| DU62375 | FLAG LRRK2 T2356I | pCMV5 |
| DU27381 | FLAG LRRK2 G2385R | pCMV5 |
| DU80488 | FLAG LRRK2 V2390A | pCMV5 |
| DU62376 | FLAG LRRK2 V2390M | pCMV5 |
| DU72045 | FLAG LRRK2 H2391Q | pCMV5 |
| DU80698 | FLAG LRRK2 E2395K | pCMV5 |
| DU26735 | FLAG LRRK2 M2397T | pCMV5 |
| DU80367 | FLAG LRRK2 T2423S | pCMV5 |
| DU80496 | FLAG LRRK2 I2434V | pCMV5 |
| DU72390 | FLAG LRRK2 L2436P | pCMV5 |
| DU62393 | FLAG LRRK2 L2439I | pCMV5 |
| DU80461 | FLAG LRRK2 A2461V | pCMV5 |
| DU80470 | FLAG LRRK2 L2463P | pCMV5 |
| DU17142 | FLAG LRRK2 L2466H | pCMV5 |
| DU61468 | FLAG LRRK2 K2467Q | pCMV5 |
| DU72385 | FLAG LRRK2 Q2484K | pCMV5 |
| DU80462 | FLAG LRRK2 Q2490H | pCMV5 |
| DU30901 | FLAG LRRK2 T2494I | pCMV5 |
| DU72429 | FLAG LRRK2 V2495I | pCMV5 |
| DU77911 | FLAG LRRK2 I1852V | pCMV5 |
| DU72189 | FLAG LRRK2 P1865R | pCMV5 |
| DU72363 | FLAG LRRK2 M1869V | pCMV5 |
| DU13169 | FLAG LRRK2 M1869T | pCMV5 |
| DU80144 | FLAG LRRK2 D1887N | pCMV5 |
| DU80145 | FLAG LRRK2 G1891A | pCMV5 |
| DU72187 | FLAG LRRK2 G1900E | pCMV5 |
| DU72366 | FLAG LRRK2 L1914I | pCMV5 |
| DU13079 | FLAG LRRK2 R1941H | pCMV5 |
| DU77852 | FLAG LRRK2 S1954F | pCMV5 |
| DU77725 | FLAG LRRK2 R1957G | pCMV5 |
| DU80463 | FLAG LRRK2 Q1961R | pCMV5 |
| DU62837 | FLAG LRRK2 I1991V | pCMV5 |
| DU13880 | FLAG LRRK2 Y2006H | pCMV5 |
| DU72368 | FLAG LRRK2 A2010T | pCMV5 |
| DU13080 | FLAG LRRK2 I2012T | pCMV5 |
| DU10129 | FLAG LRRK2 G2019S | pCMV5 |
| DU13081 | FLAG LRRK2 I2020T | pCMV5 |
| DU72362 | FLAG LRRK2 I2020S | pCMV5 |
| DU44952 | FLAG LRRK2 I2020L | pCMV5 |
| DU17135 | FLAG LRRK2 T2031S | pCMV5 |
| DU80365 | FLAG LRRK2 L2063* | pCMV5 |
| DU77780 | FLAG LRRK2 G2071D | pCMV5 |
| DU80464 | FLAG LRRK2 V2074I | pCMV5 |
| DU26721 | FLAG LRRK2 N2081D | pCMV5 |
| DU72353 | FLAG LRRK2 D2084N | pCMV5 |
| DU80151 | FLAG LRRK2 E2108K | pCMV5 |
| DU80152 | FLAG LRRK2 N2133S | pCMV5 |
| DU72414 | FLAG LRRK2 C2139S | pCMV5 |
| DU17140 | FLAG LRRK2 T2141M | pCMV5 |
| DU72441 | FLAG LRRK2 R2143H | pCMV5 |
| DU17141 | FLAG LRRK2 R2143M | pCMV5 |
| DU80697 | FLAG LRRK2 V2150I | pCMV5 |
| DU80153 | FLAG LRRK2 W2168L | pCMV5 |
| DU62374 | FLAG LRRK2 D2175H | pCMV5 |
| DU62391 | FLAG LRRK2 Y2189C | pCMV5 |
| DU77726 | FLAG LRRK2 S2213C | pCMV5 |
| DU80154 | FLAG LRRK2 H2236R | pCMV5 |
| DU80473 | FLAG LRRK2 N2251T | pCMV5 |
| DU72451 | FLAG LRRK2 K2255M | pCMV5 |
| DU80489 | FLAG LRRK2 N2261K | pCMV5 |
| DU72386 | FLAG LRRK2 K2278E | pCMV5 |
| DU80158 | FLAG LRRK2 G2294R | pCMV5 |
| DU72453 | FLAG LRRK2 V2296F | pCMV5 |
| DU62004 | FLAG LRRK2 N2308D | pCMV5 |
| DU61451 | FLAG LRRK2 T2310A | pCMV5 |
| DU72357 | FLAG LRRK2 T2310M | pCMV5 |
| DU62005 | FLAG LRRK2 N2313S | pCMV5 |
| DU80465 | FLAG LRRK2 I2323F | pCMV5 |
| DU80155 | FLAG LRRK2 I2336V | pCMV5 |
| DU62502 | FLAG LRRK2 S2350I | pCMV5 |
| DU62375 | FLAG LRRK2 T2356I | pCMV5 |
| DU27381 | FLAG LRRK2 G2385R | pCMV5 |
| DU80488 | FLAG LRRK2 V2390A | pCMV5 |
| DU62376 | FLAG LRRK2 V2390M | pCMV5 |
| DU72045 | FLAG LRRK2 H2391Q | pCMV5 |
| DU80698 | FLAG LRRK2 E2395K | pCMV5 |
| DU26735 | FLAG LRRK2 M2397T | pCMV5 |
| DU80367 | FLAG LRRK2 T2423S | pCMV5 |
| DU80496 | FLAG LRRK2 I2434V | pCMV5 |
| DU72390 | FLAG LRRK2 L2436P | pCMV5 |
| DU62393 | FLAG LRRK2 L2439I | pCMV5 |
| DU80461 | FLAG LRRK2 A2461V | pCMV5 |
| DU80470 | FLAG LRRK2 L2463P | pCMV5 |
| DU17142 | FLAG LRRK2 L2466H | pCMV5 |
| DU61468 | FLAG LRRK2 K2467Q | pCMV5 |
| DU72385 | FLAG LRRK2 Q2484K | pCMV5 |
| DU80462 | FLAG LRRK2 Q2490H | pCMV5 |
| DU30901 | FLAG LRRK2 T2494I | pCMV5 |
| DU72429 | FLAG LRRK2 V2495I | pCMV5 |
| DU80711 | FLAG LRRK2 T2524A | pCMV5 |
| DU68766 | FLAG LRRK2 R1628P-G2385R | pCMV5 |
| DU77660 | FLAG LRRK2 S455P-R1441H | pCMV5 |

All plasmids used in this study are available from MRC PPU Reagents and Services (https://mrcppureagents.dundee.ac.uk).

**Supplementary Table 5: Demographic characteristics of LRRK2 variant carriers.**

| Sample ID | LRRK2 Variant | PD Status | Age Range | AAO | Family History (Yes/No) |
| --- | --- | --- | --- | --- | --- |
| L2 | *R506P* | PD | N/A | N/A | N/A |
| L4 | *A1440P* | PD | 60-69 | 65-69 | Yes |
| L6 | *R1441C* | PD | 40-49 | 25-29 | N/A |
| L8 | *Y1699C* | NMC | 30-29 | - | Yes |
| L9 | *Y1699C* | PD | 70-79 | 55-59 | Yes |
| L10 | *Y1699C* | PD | 50-59 | 50-54 | Yes |
| L12 | *F1700L* | PD | 70-79 | 65-79 | Yes |
| L14 | *G2019S* | PD | 50-59 | 45-49 | Yes |
| L16 | *R1325Q* | PD | 50-59 | 50-54 | No |
| L18 | *R1441G* | PD | 70-79 | 65-69 | N/A |
| L20 | *V1447L* | PD | 50-59 | 40-44 | No |
| L22 | *G2019S* | PD | N/A | N/A | N/A |
| L23 | *I2020T* | PD | N/A | N/A | N/A |

Summary of identified LRRK2 variants and PD status in patients recruited across Europe and the United States. Age at assessment reported in 10-year age ranges, age at onset (AAO) reported in 5-year age ranges, and family history are shown where available. N/A = not available. Some participants have been reported before. [4-6]

**Supplementary Table 6. MDSGene pathogenicity classification integrated with LRRK2 kinase activity**

| **MDS - ACMG** | **Variant** | **CADD score** | **Domain** | **Con-Surf** | **GnomAD allele counts** | **mean pRab10 / total Rab10** | **SD** | **Repl.**  **(n)** | **Functional Group** | **Con-cordance** |
| --- | --- | --- | --- | --- | --- | --- | --- | --- | --- | --- |
| **Benign** | R1514Q | 21.4 | COR-A | 9 | 11182 | 0.95 | 0.54 | 5 | WT-like | yes |
|  | H115P | 25.7 | ARM | 8 | 242 | 1.44 | 0.38 | 5 | borderline | yes |
| **Likely benign** | L119P | 26.5 | ARM | 6 | 2922 | 0.80 | 0.27 | 8 | WT-like | yes |
|  | M262V | 0.004 | ARM | 1 | 64 | 0.89 | 0.47 | 5 | WT-like | yes |
|  | P755L | 21.0 | ANK | 7 | 395 | 0.80 | 0.34 | 6 | WT-like | yes |
|  | S865F | 21.2 | LRR | 3 | 290 | 1.08 | 0.75 | 5 | WT-like | yes |
|  | C925Y | 1.534 | LRR | 8 | 35 | 1.01 | 0.26 | 5 | WT-like | yes |
|  | I1371V | 19.30 | ROC | 9 | 1304 | 0.79 | 0.41 | 9 | Reduced | yes |
|  | H2391Q | 7.442 | WD40 | 5 | 39 | 1.18 | 0.31 | 5 | WT-like | yes |
|  | R767H | 17.88 | ANK | 4 | 44 | 1.29 | 0.67 | 18 | WT-like | no |
|  | R1067Q | 31 | LRR | 9 | 30 | 2.21 | 0.85 | 24 | Activating | yes |
| **Likely pathogenic** | N1437D | 25.6 | ROC | 9 | Not present | 3.22 | 1.47 | 10 | Activating | yes |
|  | N1437S | 24.0 | ROC | 9 | N/A | 0.98 | 0.15 | 5 | WT-like | no |
|  | R1728H | 29.1 | COR-B | 9 | 13 | 2.65 | 0.98 | 19 | Activating | yes |
|  | S1761R | 25.2 | COR-B | 9 | Not present | 3.35 | 1.37 | 19 | Activating | yes |
|  | I2020L | 23.3 | KIN | 9 | 1 | 0.64 | 0.33 | 3 | WT-like | no |
|  | I2020S | 26.4 | KIN | 9 | Not present | 3.15 | 1.50 | 17 | Activating | yes |
|  | T2031S | 18.74 | KIN | 9 | 1 | 1.54 | 0.63 | 19 | Activating | yes |
|  | S1954F | 29.2 | KIN | 9 | Not present | 0.26 | 0.14 | 3 | Reduced | yes |
|  | R1325Q | 31 | LRR | 9 | 591 | 1.49 | 0.77 | 19 | Activating | yes |
|  | N1437H | 25.3 | ROC | 9 | Not present | 4.05 | 1.89 | 21 | Activating | yes |
| **Pathogenic** | R1441C | 23.5 | ROC | 9 | 39 | 2.94 | 1.11 | 23 | Activating | yes |
|  | R1441G | 21.6 | ROC | 9 | 5 | 3.29 | 1.07 | 31 | Activating | yes |
|  | R1441H | 22.8 | ROC | 9 | 11 | 2.32 | 1.62 | 19 | Activating | yes |
|  | R1441S | 21.3 | ROC | 9 | Not present | 2.89 | 0.91 | 20 | Activating | yes |
|  | A1442P | 26.0 | ROC | 9 | Not present | 2.58 | 1.04 | 19 | Activating | yes |
|  | V1447M | 24.9 | ROC | 9 | Not present | 2.74 | 0.83 | 22 | Activating | yes |
|  | Y1699C | 26.8 | COR-B | 7 | Not present | 5.55 | 1.92 | 23 | Activating | yes |
|  | F1700L | 25.7 | COR-B | 9 | Not present | 4.45 | 1.61 | 17 | Activating | yes |
|  | L1795F | 17.38 | COR-B | 9 | 2 | 4.51 | 1.50 | 19 | Activating | yes |
|  | G2019S | 31 | KIN | 8 | 664 | 1.44 | 0.45 | 25 | Activating | yes |
|  | I2020T | 25.4 | KIN | 9 | Not present | 3.43 | 0.86 | 20 | Activating | yes |

Variants are grouped according to MDSGene ACMG pathogenicity classification and annotated with kinase activity measured by pRab10/total Rab10 phosphorylation in a standardized cellular overexpression assay. Functional groups were defined as activating, WT-like, or reduced activity. Mean pRab10/total Rab10 values are shown with standard deviation (SD) and number of independent biological replicates (Repl., n). Additional annotations include CADD score, ConSurf conservation score, and gnomAD allele counts[7]. Concordance indicates agreement between MDSGene classification and observed functional effect.

**Supplementary Table 7. Population frequencies of functionally characterized LRRK2 variants in the GP2[****8****] and PD GENEration[****9****] cohorts.**

| **Variant** | **Domain** | **MDS**  **ACMG** | **Functional Activation** | **Chr12 position position (hg38)** | **GP2 WGS AC**  **PD** | **GP2 WGS AN PD** | **GP2 WGS AF PD** | **GP2 WGS AC HC** | **GP2 WGS AN HC** | **GP2 WGS AF HC** | **GP2 CES AC PD** | **GP2 CES AN PD** | **GP2 CES AF PD** | **Chr12 position (hg19)** | **PDGENEration AC** | **PDGENEration AF** | **PDGENEration AC in AJ** | **PDGENEration AF in AJ** |
| --- | --- | --- | --- | --- | --- | --- | --- | --- | --- | --- | --- | --- | --- | --- | --- | --- | --- | --- |
| E10K | ARM | VUS | No | NA | NA | NA | NA | NA | NA | NA | NA | NA | NA | NA | 0 | 0 | 0 | 0 |
| K53R | ARM | VUS | No | NA | NA | NA | NA | NA | NA | NA | NA | NA | NA | NA | 0 | 0 | 0 | 0 |
| V80M | ARM | N/A | No | NA | NA | NA | NA | NA | NA | NA | NA | NA | NA | 40626076:G:A | 3 | 5.13E-05 | 0 | 0 |
| M96T | ARM | N/A | No | 40232323:T:C | 1 | 16500 | 6.06E-05 | 0 | 15116 | 0 |  |  |  | NA | 0 | 0 | 0 | 0 |
| M100T | ARM | VUS | No | NA | NA | NA | NA | NA | NA | NA | NA | NA | NA | NA | 0 | 0 | 0 | 0 |
| D104Y | ARM | N/A | No | NA | NA | NA | NA | NA | NA | NA | NA | NA | NA | NA | 0 | 0 | 0 | 0 |
| H115P | ARM | LB | No | 40232380:A:C | 0 | 16502 | 0 | 1 | 15118 | 6.61E-05 | 3 | 15720 | 1.91E-04 | 40626182:A:C | 13 | 2.23E-04 | 3 | 5.22E-04 |
| Q116R | ARM | VUS | No | NA | NA | NA | NA | NA | NA | NA | NA | NA | NA | 40626185:A:G | 6 | 1.03E-04 | 0 | 0 |
| L119P | ARM | LB | No | 40235634:T:C | 41 | 16500 | 2.48E-03 | 37 | 15116 | 2.45E-03 | 44 | 15664 | 2.81E-03 | 40629436:T:C | 131 | 2.24E-03 | 3 | 5.22E-04 |
| S128G | ARM | VUS | No | NA | NA | NA | NA | NA | NA | NA | NA | NA | NA | NA | 0 | 0 | 0 | 0 |
| T149S | ARM | N/A | No | 40237978:C:G | 0 | 16492 | 0 | 1 | 15112 | 6.62E-05 |  |  |  | 40631780:C:G | 1 | 1.71E-05 | 0 | 0 |
| L153W | ARM | VUS | No | NA | NA | NA | NA | NA | NA | NA | NA | NA | NA | NA | 0 | 0 | 0 | 0 |
| F160L | ARM | N/A | No | NA | NA | NA | NA | NA | NA | NA | NA | NA | NA | NA | 0 | 0 | 0 | 0 |
| N173S | ARM | VUS | No | 40238050:A:G | 0 | 16496 | 0 | 1 | 15116 | 6.62E-05 |  |  |  | NA | 0 | 0 | 0 | 0 |
| V176L | ARM | N/A | No | 40238058:G:C | 0 | 16494 | 0 | 1 | 15116 | 6.62E-05 |  |  |  | NA | 0 | 0 | 0 | 0 |
| S192A | ARM | VUS | No | NA | NA | NA | NA | NA | NA | NA | NA | NA | NA | NA | 0 | 0 | 0 | 0 |
| E193A | ARM | VUS | No | NA | NA | NA | NA | NA | NA | NA | NA | NA | NA | NA | 0 | 0 | 0 | 0 |
| E193K | ARM | VUS | No | NA | NA | NA | NA | NA | NA | NA | NA | NA | NA | NA | 0 | 0 | 0 | 0 |
| A211V | ARM | VUS | No | 40240543:C:T | 0 | 16486 | 0 | 1 | 15114 | 6.62E-05 |  |  |  | 40634345:C:T | 8 | 1.37E-04 | 2 | 3.48E-04 |
| C228S | ARM | VUS | No | 40240594:G:C | 5 | 16494 | 3.03E-04 | 1 | 15118 | 6.61E-05 | 2 | 15720 | 1.27E-04 | 40634396:G:C | 9 | 1.54E-04 | 1 | 1.74E-04 |
| H230R | ARM | VUS | Yes | NA | NA | NA | NA | NA | NA | NA | NA | NA | NA | NA | 0 | 0 | 0 | 0 |
| S231P | ARM | VUS | No | only S231F | NA | NA | NA | NA | NA | NA | NA | NA | NA | 40634404:T:C | 1 | 1.71E-05 | 0 | 0 |
| N238I | ARM | VUS | No | NA | NA | NA | NA | NA | NA | NA | NA | NA | NA | 40637358:A:T | 3 | 5.13E-05 | 0 | 0 |
| Y250C | ARM | VUS | No | NA | NA | NA | NA | NA | NA | NA | NA | NA | NA | NA | 0 | 0 | 0 | 0 |
| E255K | ARM | VUS | No | NA | NA | NA | NA | NA | NA | NA | NA | NA | NA | NA | 0 | 0 | 0 | 0 |
| K258N | ARM | VUS | No | NA | NA | NA | NA | NA | NA | NA | NA | NA | NA | NA | 0 | 0 | 0 | 0 |
| A259E | ARM | VUS | No | only A259S |  |  |  |  |  |  |  |  |  | 40637421:C:A | 1 | 1.71E-05 | 0 | 0 |
| M262V | ARM | LB | No | 40243627:A:G | 1 | 16482 | 6.07E-05 | 0 | 15108 | 0 |  |  |  | 40637429:A:G | 3 | 5.13E-05 | 0 | 0 |
| L286V | ARM | VUS | No | 40249843:C:G | 8 | 16488 | 4.85E-04 | 2 | 15112 | 1.32E-04 | 4 | 15720 | 2.54E-04 | 40643645:C:G | 14 | 2.40E-04 | 1 | 1.74E-04 |
| V291A | ARM | VUS | No | NA | NA | NA | NA | NA | NA | NA | NA | NA | NA | NA | 0 | 0 | 0 | 0 |
| H292Q | ARM | N/A | No | 40249863:T:G | 1 | 16494 | 6.06E-05 | 0 | 15108 | 0 |  |  |  | NA | 0 | 0 | 0 | 0 |
| A306V | ARM | VUS | No | NA | NA | NA | NA | NA | NA | NA | NA | NA | NA | NA | 0 | 0 | 0 | 0 |
| L308M | ARM | VUS | No | NA | NA | NA | NA | NA | NA | NA | NA | NA | NA | NA | 0 | 0 | 0 | 0 |
| A312V | ARM | VUS | Yes | NA | NA | NA | NA | NA | NA | NA | NA | NA | NA | 40643724:C:T | 3 | 5.13E-05 | 0 | 0 |
| L313F | ARM | N/A | No | NA | NA | NA | NA | NA | NA | NA | NA | NA | NA | NA | 0 | 0 | 0 | 0 |
| T320S | ARM | VUS | No | NA | NA | NA | NA | NA | NA | NA | NA | NA | NA | NA | 0 | 0 | 0 | 0 |
| E334K | ARM | N/A | Yes | 40251273:G:A | 15 | 16482 | 9.10E-04 | 14 | 15110 | 9.27E-04 | 16 | 15718 | 1.02E-03 | 40645075:G:A | 60 | 1.03E-03 | 3 | 5.22E-04 |
| N335S | ARM | VUS | No | NA | NA | NA | NA | NA | NA | NA | NA | NA | NA | 40645079:A:G | 1 | 1.71E-05 | 0 | 0 |
| C353Y | ARM | N/A | No | NA | NA | NA | NA | NA | NA | NA | NA | NA | NA | NA | 0 | 0 | 0 | 0 |
| A356T | ARM | N/A | Yes | NA | NA | NA | NA | NA | NA | NA | NA | NA | NA | 40645141:G:A | 1 | 1.71E-05 | 0 | 0 |
| N363S | ARM | N/A | No | NA | NA | NA | NA | NA | NA | NA | NA | NA | NA | NA | 0 | 0 | 0 | 0 |
| H365N | ARM | N/A | No | Only H365Q | NA | NA | NA | NA | NA | NA | NA | NA | NA | NA | 0 | 0 | 0 | 0 |
| A370T | ARM | VUS | No | NA | NA | NA | NA | NA | NA | NA | NA | NA | NA | NA | 0 | 0 | 0 | 0 |
| L378F | ARM | VUS | Yes | 40251495:C:T | 0 | 16492 | 0 | 1 | 15114 | 6.62E-05 |  |  |  | NA | 0 | 0 | 0 | 0 |
| M379I | ARM | VUS | No | NA | NA | NA | NA | NA | NA | NA | NA | NA | NA | NA | 0 | 0 | 0 | 0 |
| H385R | ARM | N/A | No | NA | NA | NA | NA | NA | NA | NA | NA | NA | NA | NA | 0 | 0 | 0 | 0 |
| I388T | ARM | VUS | No | 40251526:T:C | 1 | 16496 | 6.06E-05 | 1 | 15118 | 6.61E-05 |  |  |  | 40645328:T:C | 2 | 3.42E-05 | 0 | 0 |
| A397T | ARM | N/A | No | NA | NA | NA | NA | NA | NA | NA | NA | NA | NA | NA | 0 | 0 | 0 | 0 |
| V414I | ARM | VUS | Yes | 40252968:G:A | 1 | 16498 | 6.06E-05 | 0 | 15114 | 0 | 2 | 15720 | 1.27E-04 | 40646770:G:A | 3 | 5.13E-05 | 0 | 0 |
| A419V | ARM | VUS | Yes | 40252984:C:T | 2 | 16496 | 1.21E-04 | 3 | 15112 | 1.99E-04 | 2 | 15720 | 1.27E-04 | 40646786:C:T | 22 | 3.77E-04 | 0 | 0 |
| L425P | ARM | VUS | No | NA | NA | NA | NA | NA | NA | NA | NA | NA | NA | NA | 0 | 0 | 0 | 0 |
| L437S | ARM | VUS | No | NA | NA | NA | NA | NA | NA | NA | NA | NA | NA | NA | 0 | 0 | 0 | 0 |
| S438P | ARM | VUS | No | NA | NA | NA | NA | NA | NA | NA | NA | NA | NA | NA | 0 | 0 | 0 | 0 |
| H442Y | ARM | N/A | No | NA | NA | NA | NA | NA | NA | NA | NA | NA | NA | 40651085:C:T | 1 | 1.71E-05 | 0 | 0 |
| S455P | ARM | N/A | Yes | NA | NA | NA | NA | NA | NA | NA | NA | NA | NA | NA | 0 | 0 | 0 | 0 |
| A459S | ARM | VUS | No | NA | NA | NA | NA | NA | NA | NA | NA | NA | NA | NA | 0 | 0 | 0 | 0 |
| S461I | ARM | VUS | Yes | 40257341:G:T | 1 | 16476 | 6.07E-05 | 0 | 15116 | 0 | 1 | 15720 | 6.36E-05 | 40651143:G:T | 5 | 8.56E-05 | 0 | 0 |
| N474S | ARM | VUS | No | NA | NA | NA | NA | NA | NA | NA | NA | NA | NA | NA | 0 | 0 | 0 | 0 |
| D478Y | ARM | N/A | No | NA | NA | NA | NA | NA | NA | NA | NA | NA | NA | NA | 0 | 0 | 0 | 0 |
| I479V | ARM | N/A | No | 40259496:A:G | 1 | 16504 | 6.06E-05 | 0 | 15118 | 0 |  |  |  | NA | 0 | 0 | 0 | 0 |
| L488P | ARM | N/A | Yes | NA | NA | NA | NA | NA | NA | NA | NA | NA | NA | NA | 0 | 0 | 0 | 0 |
| A504T | ARM | N/A | Yes | only A504V |  |  |  |  |  |  |  |  |  | NA | 0 | 0 | 0 | 0 |
| R506P | ARM | N/A | Yes | 40259578:G:C |  |  |  |  |  |  | 1 | 15720 | 6.36E-05 | 40653380:G:C | 1 | 1.71E-05 | 0 | 0 |
| R506Q | ARM | N/A | No | 40259578:G:A | 4 | 16504 | 2.42E-04 | 0 | 15118 | 0 | 3 | 15720 | 1.91E-04 | 40653380:G:A | 9 | 1.54E-04 | 1 | 1.74E-04 |
| E518K | ARM | N/A | No | 40263797:G:A |  |  |  |  |  |  | 1 | 15720 | 6.36E-05 | 40657599:G:A | 1 | 1.71E-05 | 0 | 0 |
| R521G | ARM | VUS | No | 40263806:A:G |  |  |  |  |  |  | 13 | 15720 | 8.27E-04 | 40657608:A:G | 30 | 5.13E-04 | 21 | 3.65E-03 |
| K544E | ARM | VUS | No | NA | NA | NA | NA | NA | NA | NA | NA | NA | NA | 40657677:A:G | 4 | 6.85E-05 | 0 | 0 |
| N551K | ARM | N/A | No | 40263898:C:G | 1069 | 16480 | 0.0649 | 1122 | 15110 | 0.0743 | 1122 | 15716 | 0.0714 | 40657700:C:G | 4720 | 8.08E-02 | 479 | 8.33E-02 |
| S581T | ARM | N/A | No | 40274667:T:A | 0 | 16498 | 0 | 1 | 15118 | 6.61E-05 |  |  |  | NA | 0 | 0 | 0 | 0 |
| I610T | ARM | VUS | No | NA | NA | NA | NA | NA | NA | NA | NA | NA | NA | NA | 0 | 0 | 0 | 0 |
| K616R | ARM | VUS | No | NA | NA | NA | NA | NA | NA | NA | NA | NA | NA | NA | 0 | 0 | 0 | 0 |
| S633F | ARM | VUS | No | NA | NA | NA | NA | NA | NA | NA | NA | NA | NA | NA | 0 | 0 | 0 | 0 |
| R637Q | ARM | N/A | Yes | NA | NA | NA | NA | NA | NA | NA | NA | NA | NA | 40668764:G:A | 3 | 5.13E-05 | 0 | 0 |
| A654V | ARM | VUS | No | NA | NA | NA | NA | NA | NA | NA | NA | NA | NA | NA | 0 | 0 | 0 | 0 |
| S661F | ARM | VUS | No | NA | NA | NA | NA | NA | NA | NA | NA | NA | NA | NA | 0 | 0 | 0 | 0 |
| I675M | ARM | VUS | No | 40277971:A:G | 2 | 16498 | 1.21E-04 | 2 | 15116 | 1.32E-04 |  |  |  | 40671773:A:G | 3 | 5.13E-05 | 0 | 0 |
| F691S | ARM | N/A | No | NA | NA | NA | NA | NA | NA | NA | NA | NA | NA | 40671894:T:C | 1 | 1.71E-05 | 1 | 1.74E-04 |
| M712V | ANK | N/A | No | 40278154:A:G |  |  |  |  |  |  | 1 | 15720 | 6.36E-05 | 40671956:A:G | 1 | 1.71E-05 | 0 | 0 |
| S722N | ANK | VUS | No | NA | NA | NA | NA | NA | NA | NA | NA | NA | NA | NA | 0 | 0 | 0 | 0 |
| I723V | ANK | N/A | No | 40278187:A:G | 1161 | 16500 | 0.0704 | 1024 | 15116 | 0.0677 | 1309 | 15718 | 0.0833 | 40671989:A:G | 4209 | 7.20E-02 | 653 | 1.14E-01 |
| M724V | ANK | VUS | No | NA | NA | NA | NA | NA | NA | NA | NA | NA | NA | 40671992:A:G | 1 | 1.71E-05 | 0 | 0 |
| D734N | ANK | VUS | No | NA | NA | NA | NA | NA | NA | NA | NA | NA | NA | 40672022:G:A | 2 | 3.42E-05 | 1 | 1.74E-04 |
| K739R | ANK | VUS | No | NA | NA | NA | NA | NA | NA | NA | NA | NA | NA | NA | 0 | 0 | 0 | 0 |
| P755L | ANK | LB | No | NA | NA | NA | NA | NA | NA | NA | NA | NA | NA | 40677699:C:T | 13 | 2.23E-04 | 0 | 0 |
| L757V | ANK | N/A | No | NA | NA | NA | NA | NA | NA | NA | NA | NA | NA | NA | 0 | 0 | 0 | 0 |
| S764N | ANK | VUS | No | NA | NA | NA | NA | NA | NA | NA | NA | NA | NA | 40677726:G:A | 1 | 1.71E-05 | 0 | 0 |
| R767H | ANK | LP | Yes | only R767C | NA | NA | NA | NA | NA | NA | NA | NA | NA | 40677735:G:A | 1 | 1.71E-05 | 0 | 0 |
| R767C | ANK | VUS | No | 40283932:C:T | 3 | 16502 | 1.82E-04 | 2 | 15118 | 1.32E-04 | 2 | 15720 | 1.27E-04 | 40677734:C:T | 5 | 8.56E-05 | 0 | 0 |
| T776M | ANK | VUS | No | 40283960:C:T | 1 | 16498 | 6.06E-05 | 1 | 15118 | 6.61E-05 |  |  |  | NA | 0 | 0 | 0 | 0 |
| I777M | ANK | VUS | No | NA | NA | NA | NA | NA | NA | NA | NA | NA | NA | NA | 0 | 0 | 0 | 0 |
| S784R | ANK | VUS | No | 40283985:C:A | 0 | 16502 | 0 | 1 | 15118 | 6.61E-05 |  |  |  | 40677787:C:A | 1 | 1.71E-05 | 0 | 0 |
| I786F | ANK | VUS | No | NA | NA | NA | NA | NA | NA | NA | NA | NA | NA | NA | 0 | 0 | 0 | 0 |
| R792K | ANK | VUS | No | NA | NA | NA | NA | NA | NA | NA | NA | NA | NA | NA | 0 | 0 | 0 | 0 |
| R793M | ANK | VUS | No | 40284011:G:T | 7 | 16492 | 4.24E-04 | 8 | 15118 | 5.29E-04 | 7 | 15720 | 4.45E-04 | 40677813:G:T | 27 | 4.62E-04 | 0 | 0 |
| I810V | LRR | N/A | No | NA | NA | NA | NA | NA | NA | NA | NA | NA | NA | NA | 0 | 0 | 0 | 0 |
| K812N | LRR | N/A | No | NA | NA | NA | NA | NA | NA | NA | NA | NA | NA | NA | 0 | 0 | 0 | 0 |
| W817C | LRR | N/A | No | NA | NA | NA | NA | NA | NA | NA | NA | NA | NA | NA | 0 | 0 | 0 | 0 |
| S827F | LRR | VUS | No | NA | NA | NA | NA | NA | NA | NA | NA | NA | NA | 40677915:C:T | 1 | 1.71E-05 | 0 | 0 |
| I844N | LRR | VUS | No | NA | NA | NA | NA | NA | NA | NA | NA | NA | NA | NA | 0 | 0 | 0 | 0 |
| M848T | LRR | VUS | No | NA | NA | NA | NA | NA | NA | NA | NA | NA | NA | NA | 0 | 0 | 0 | 0 |
| S860R | LRR | N/A | No | NA | NA | NA | NA | NA | NA | NA | NA | NA | NA | NA | 0 | 0 | 0 | 0 |
| S865F | LRR | LB | No | 40287444:C:T | 2 | 16496 | 1.21E-04 | 2 | 15116 | 1.32E-04 | 9 | 15720 | 5.73E-04 | 40681246:C:T | 36 | 6.16E-04 | 26 | 4.52E-03 |
| D867N | LRR | N/A | No | NA | NA | NA | NA | NA | NA | NA | NA | NA | NA | NA | 0 | 0 | 0 | 0 |
| S885N | LRR | N/A | No | NA | NA | NA | NA | NA | NA | NA | NA | NA | NA | NA | 0 | 0 | 0 | 0 |
| S885C | LRR | VUS | No | NA | NA | NA | NA | NA | NA | NA | NA | NA | NA | NA | 0 | 0 | 0 | 0 |
| E899D | LRR | VUS | No | 40293552:A:C | 1 | 16492 | 6.06E-05 | 0 | 15118 | 0 |  |  |  | 40687354:A:C | 4 | 6.85E-05 | 2 | 3.48E-04 |
| R918Q | LRR | N/A | No | NA | NA | NA | NA | NA | NA | NA | NA | NA | NA | NA | 0 | 0 | 0 | 0 |
| Q923H | LRR | VUS | No | 40293624:G:C | 6 | 16496 | 3.64E-04 | 8 | 15110 | 5.29E-04 | 2 | 15720 | 1.27E-04 | 40687426:G:C | 13 | 2.23E-04 | 2 | 3.48E-04 |
| C925Y | LRR | LB | No | NA | NA | NA | NA | NA | NA | NA | NA | NA | NA | 40687431:G:A | 3 | 5.13E-05 | 0 | 0 |
| Q930R | LRR | VUS | No | NA | NA | NA | NA | NA | NA | NA | NA | NA | NA | NA | 0 | 0 | 0 | 0 |
| R931K | LRR | N/A | No | NA | NA | NA | NA | NA | NA | NA | NA | NA | NA | NA | 0 | 0 | 0 | 0 |
| D941Y | LRR | VUS | No | NA | NA | NA | NA | NA | NA | NA | NA | NA | NA | 40688659:G:T | 1 | 1.71E-05 | 0 | 0 |
| D944V | LRR | VUS | No | 40294867:A:T | 2 | 16434 | 1.22E-04 | 1 | 15100 | 6.62E-05 |  |  |  | 40688669:A:T | 2 | 3.42E-05 | 1 | 1.74E-04 |
| R948Q | LRR | VUS | No | NA | NA | NA | NA | NA | NA | NA | NA | NA | NA | NA | 0 | 0 | 0 | 0 |
| S954T | LRR | N/A | No | NA | NA | NA | NA | NA | NA | NA | NA | NA | NA | NA | 0 | 0 | 0 | 0 |
| S958L | LRR | VUS | No | NA | NA | NA | NA | NA | NA | NA | NA | NA | NA | 40688711:C:T | 3 | 5.13E-05 | 0 | 0 |
| R969K | LRR | N/A | No | 40295454:G:A | 1 | 16496 | 6.06E-05 | 0 | 15116 | 0 |  |  |  | 40689256:G:A | 2 | 3.42E-05 | 0 | 0 |
| D972G | LRR | VUS | No | 40295463:A:G | 3 | 16504 | 1.82E-04 | 1 | 15116 | 6.62E-05 | 1 | 15720 | 6.36E-05 | 40689265:A:G | 3 | 5.13E-05 | 0 | 0 |
| S973G | LRR | VUS | No | only S973N | NA | NA | NA | NA | NA | NA | NA | NA | NA | NA | 0 | 0 | 0 | 0 |
| S973N | LRR | VUS | No | 40295466:G:A | 2 | 16502 | 1.21E-04 | 0 | 15118 | 0 |  |  |  | 40689268:G:A | 2 | 3.42E-05 | 1 | 1.74E-04 |
| R981K | LRR | VUS | Yes | NA | NA | NA | NA | NA | NA | NA | NA | NA | NA | NA | 0 | 0 | 0 | 0 |
| I984V | LRR | N/A | No | 40295498:A:G | 1 | 16500 | 6.06E-05 | 0 | 15118 | 0 |  |  |  | NA | 0 | 0 | 0 | 0 |
| S1007T | LRR | VUS | No | NA | NA | NA | NA | NA | NA | NA | NA | NA | NA | NA | 0 | 0 | 0 | 0 |
| K1015R | LRR | N/A | No | 40295592:A:G | 2 | 16500 | 1.21E-04 | 0 | 15116 | 0 |  |  |  | NA | 0 | 0 | 0 | 0 |
| H1019R | LRR | VUS | Yes | NA | NA | NA | NA | NA | NA | NA | NA | NA | NA | NA | 0 | 0 | 0 | 0 |
| Q1029H | LRR | VUS | No | 40295635:G:T | 0 | 16498 | 0 | 1 | 15116 | 6.62E-05 | 1 | 15720 | 6.36E-05 | 40689437:G:T | 2 | 3.42E-05 | 0 | 0 |
| R1067Q | LRR | LP | Yes | NA | NA | NA | NA | NA | NA | NA | NA | NA | NA | 40692148:G:A | 3 | 5.13E-05 | 0 | 0 |
| N1089S | LRR | VUS | No | NA | NA | NA | NA | NA | NA | NA | NA | NA | NA | NA | 0 | 0 | 0 | 0 |
| S1096C | LRR | VUS | No | 40298433:C:G | 0 | 16498 | 0 | 1 | 15116 | 6.62E-05 | 2 | 15720 | 1.27E-04 | 40692235:C:G | 5 | 8.56E-05 | 1 | 1.74E-04 |
| P1099S | LRR | N/A | No | NA | NA | NA | NA | NA | NA | NA | NA | NA | NA | NA | 0 | 0 | 0 | 0 |
| Q1111H | LRR | N/A | No | 40298479:G:T | 0 | 16494 | 0 | 3 | 15118 | 1.98E-04 | 1 | 15720 | 6.36E-05 | 40692281:G:T | 179 | 3.06E-03 | 2 | 3.48E-04 |
| I1122V | LRR | VUS | No | NA | NA | NA | NA | NA | NA | NA | NA | NA | NA | NA | 0 | 0 | 0 | 0 |
| L1128M | LRR | VUS | No | NA | NA | NA | NA | NA | NA | NA | NA | NA | NA | NA | 0 | 0 | 0 | 0 |
| K1132E | LRR | N/A | No | NA | NA | NA | NA | NA | NA | NA | NA | NA | NA | NA | 0 | 0 | 0 | 0 |
| K1138E | LRR | N/A | No | 40299173:A:G |  |  |  |  |  |  | 1 | 15720 | 6.36E-05 | 40692975:A:G | 1 | 1.71E-05 | 0 | 0 |
| I1141V | LRR | N/A | No | NA | NA | NA | NA | NA | NA | NA | NA | NA | NA | NA | 0 | 0 | 0 | 0 |
| A1151T | LRR | VUS | No | 40299212:G:A | 3 | 16494 | 1.82E-04 | 0 | 15116 | 0 | 2 | 15720 | 1.27E-04 | 40693014:G:A | 7 | 1.20E-04 | 0 | 0 |
| C1152F | LRR | VUS | No | NA | NA | NA | NA | NA | NA | NA | NA | NA | NA | NA | 0 | 0 | 0 | 0 |
| S1159R | LRR | VUS | No | NA | NA | NA | NA | NA | NA | NA | NA | NA | NA | 40693040:T:G | 1 | 1.71E-05 | 0 | 0 |
| A1160D | LRR | N/A | No | NA | NA | NA | NA | NA | NA | NA | NA | NA | NA | NA | 0 | 0 | 0 | 0 |
| L1165P | LRR | VUS | No | NA | NA | NA | NA | NA | NA | NA | NA | NA | NA | NA | 0 | 0 | 0 | 0 |
| S1181Y | LRR | VUS | No | NA | NA | NA | NA | NA | NA | NA | NA | NA | NA | NA | 0 | 0 | 0 | 0 |
| I1192V | LRR | VUS | No | 40302866:A:G | 2 | 16266 | 1.23E-04 | 0 | 15066 | 0 |  |  |  | 40696668:A:G | 6 | 1.03E-04 | 0 | 0 |
| I1192M | LRR | VUS | No | only I1192V | NA | NA | NA | NA | NA | NA | NA | NA | NA | NA | 0 | 0 | 0 | 0 |
| A1215T | LRR | VUS | No | 40304000:G:A | 4 | 16500 | 2.42E-04 | 1 | 15116 | 6.62E-05 | 2 | 15720 | 1.27E-04 | 40697802:G:A | 6 | 1.03E-04 | 0 | 0 |
| H1216R | LRR | VUS | No | NA | NA | NA | NA | NA | NA | NA | NA | NA | NA | NA | 0 | 0 | 0 | 0 |
| N1221K | LRR | LP | - | NA | NA | NA | NA | NA | NA | NA | NA | NA | NA | NA | 0 | 0 | 0 | 0 |
| S1228T | LRR | VUS | No | 40304040:G:C | 7 | 16498 | 4.24E-04 | 1 | 15114 | 6.62E-05 | 8 | 15720 | 5.09E-04 | 40697842:G:C | 15 | 2.57E-04 | 0 | 0 |
| P1262A | LRR | N/A | No | 40305791:C:G | 0 | 16490 | 0 | 3 | 15112 | 1.99E-04 | 2 | 15718 | 1.27E-04 | 40699593:C:G | 58 | 9.93E-04 | 1 | 1.74E-04 |
| T1271I | LRR | VUS | No | NA | NA | NA | NA | NA | NA | NA | NA | NA | NA | NA | 0 | 0 | 0 | 0 |
| L1281L | LRR | LB | - | NA | NA | NA | NA | NA | NA | NA | NA | NA | NA | NA | 0 | 0 | 0 | 0 |
| W1295R | LRR | VUS | No | NA | NA | NA | NA | NA | NA | NA | NA | NA | NA | NA | 0 | 0 | 0 | 0 |
| L1304F | LRR | VUS | No | NA | NA | NA | NA | NA | NA | NA | NA | NA | NA | NA | 0 | 0 | 0 | 0 |
| C1313* | LRR | N/A | No | 40305946:T:A | 0 | 16490 | 0 | 1 | 15114 | 6.62E-05 | NA | NA | NA | 40699748:T:A | 4 | 6.85E-05 | 0 | 0 |
| R1320S | LRR | VUS | No | NA | NA | NA | NA | NA | NA | NA | NA | NA | NA | 40702269:G:T | 17 | 2.91E-04 | 0 | 0 |
| R1325Q | LRR | P | Yes | 40308481:G:A | 16 | 16496 | 9.70E-04 | 8 | 15118 | 5.29E-04 | 4 | 15720 | 2.54E-04 | 40702283:G:A | 27 | 4.62E-04 | 0 | 0 |
| V1330M | LRR | N/A | No | NA | NA | NA | NA | NA | NA | NA | NA | NA | NA | NA | 0 | 0 | 0 | 0 |
| R1334Q | LRR | VUS | Yes | 40308508:G:A | 4 | 16498 | 2.42E-04 | 2 | 15112 | 1.32E-04 | 1 | 15720 | 6.36E-05 | 40702310:G:A | 3 | 5.13E-05 | 0 | 0 |
| I1339M | ROC | VUS | No | NA | NA | NA | NA | NA | NA | NA | NA | NA | NA | NA | 0 | 0 | 0 | 0 |
| V1340M | ROC | VUS | No | NA | NA | NA | NA | NA | NA | NA | NA | NA | NA | NA | 0 | 0 | 0 | 0 |
| K1347R | ROC | N/A | No | 40308547:A:G | 1 | 16498 | 6.06E-05 | 0 | 15118 | 0 |  |  |  | NA | 0 | 0 | 0 | 0 |
| T1348P | ROC | N/A | No | NA | NA | NA | NA | NA | NA | NA | NA | NA | NA | NA | 0 | 0 | 0 | 0 |
| Q1353K | ROC | VUS | Yes | 40308564:C:A | 1 | 16500 | 6.06E-05 | 2 | 15118 | 1.32E-04 | 1 | 15720 | 6.36E-05 | 40702366:C:A | 3 | 5.13E-05 | 0 | 0 |
| M1355K | ROC | N/A | No | NA | NA | NA | NA | NA | NA | NA | NA | NA | NA | NA | 0 | 0 | 0 | 0 |
| S1366N | ROC | N/A | No | NA | NA | NA | NA | NA | NA | NA | NA | NA | NA | NA | 0 | 0 | 0 | 0 |
| V1369A | ROC | VUS | No | NA | NA | NA | NA | NA | NA | NA | NA | NA | NA | NA | 0 | 0 | 0 | 0 |
| I1371V | ROC | LB | No | 40308618:A:G | 5 | 16504 | 3.03E-04 | 10 | 15118 | 6.61E-04 | 12 | 15720 | 7.63E-04 | 40702420:A:G | 33 | 5.65E-04 | 0 | 0 |
| V1373M | ROC | VUS | Yes | 40308624:G:A |  |  |  |  |  |  | 2 | 15720 | 1.27E-04 | 40702426:G:A | 3 | 5.13E-05 | 0 | 0 |
| Q1379R | ROC | VUS | No | NA | NA | NA | NA | NA | NA | NA | NA | NA | NA | NA | 0 | 0 | 0 | 0 |
| L1388I | ROC | VUS | Yes | NA | NA | NA | NA | NA | NA | NA | NA | NA | NA | 40702471:C:A | 3 | 5.13E-05 | 0 | 0 |
| V1389I | ROC | VUS | No | 40308672:G:A | 2 | 16474 | 1.21E-04 | 4 | 15108 | 2.65E-04 | 3 | 15720 | 1.91E-04 | 40702474:G:A | 6 | 1.03E-04 | 0 | 0 |
| A1396S | ROC | N/A | No | NA | NA | NA | NA | NA | NA | NA | NA | NA | NA | NA | 0 | 0 | 0 | 0 |
| R1398H | ROC | N/A | No | 40309109:G:A | 1099 | 16498 | 0.0666 | 1124 | 15114 | 0.0744 | 1123 | 15718 | 0.0714 | 40702911:G:A | 4635 | 7.93E-02 | 473 | 8.22E-02 |
| Y1402C | ROC | VUS | Yes | NA | NA | NA | NA | NA | NA | NA | NA | NA | NA | 40702923:A:G | 1 | 1.71E-05 | 0 | 0 |
| S1403R | LRR | LP | - | only S1403G and S1403N | NA | NA | NA | NA | NA | NA | NA | NA | NA | NA | 0 | 0 | 0 | 0 |
| H1405Q | ROC | VUS | No | NA | NA | NA | NA | NA | NA | NA | NA | NA | NA | NA | 0 | 0 | 0 | 0 |
| T1410M | ROC | N/A | No | 40309145:C:T | 1 | 16498 | 6.06E-05 | 1 | 15118 | 6.61E-05 | 2 | 15720 | 1.27E-04 | 40702947:C:T | 60 | 1.03E-03 | 0 | 0 |
| A1413T | ROC | VUS | Yes | NA | NA | NA | NA | NA | NA | NA | NA | NA | NA | NA | 0 | 0 | 0 | 0 |
| Y1415E | ROC | N/A | No | NA | NA | NA | NA | NA | NA | NA | NA | NA | NA | NA | 0 | 0 | 0 | 0 |
| A1417V | ROC | N/A | Yes | NA | NA | NA | NA | NA | NA | NA | NA | NA | NA | NA | 0 | 0 | 0 | 0 |
| A1417E | ROC | N/A | No | NA | NA | NA | NA | NA | NA | NA | NA | NA | NA | NA | 0 | 0 | 0 | 0 |
| Y1419C | ROC | N/A | No | NA | NA | NA | NA | NA | NA | NA | NA | NA | NA | NA | 0 | 0 | 0 | 0 |
| E1427G | ROC | VUS | No | NA | NA | NA | NA | NA | NA | NA | NA | NA | NA | NA | 0 | 0 | 0 | 0 |
| L1435E | ROC | N/A | No | NA | NA | NA | NA | NA | NA | NA | NA | NA | NA | NA | 0 | 0 | 0 | 0 |
| L1435F | ROC | N/A | Yes | NA | NA | NA | NA | NA | NA | NA | NA | NA | NA | NA | 0 | 0 | 0 | 0 |
| F1436S | ROC | N/A | Yes | NA | NA | NA | NA | NA | NA | NA | NA | NA | NA | NA | 0 | 0 | 0 | 0 |
| F1436L | ROC | VUS | Yes | NA | NA | NA | NA | NA | NA | NA | NA | NA | NA | NA | 0 | 0 | 0 | 0 |
| N1437H | ROC | P | Yes | 40309225:A:C |  |  |  |  |  |  | 1 | 15720 | 6.36E-05 | 40703027:A:C | 1 | 1.71E-05 | 0 | 0 |
| N1437D | ROC | LP | Yes | NA | NA | NA | NA | NA | NA | NA | NA | NA | NA | NA | 0 | 0 | 0 | 0 |
| N1437S | ROC | LP | No | 40309226:A:G | 1 | 16500 | 6.06E-05 | 0 | 15116 | 0 |  |  |  | 40703028:A:G | 3 | 5.13E-05 | 0 | 0 |
| I1438E | ROC | N/A | Yes | NA | NA | NA | NA | NA | NA | NA | NA | NA | NA | NA | 0 | 0 | 0 | 0 |
| I1438T | ROC | N/A | Yes | NA | NA | NA | NA | NA | NA | NA | NA | NA | NA | NA | 0 | 0 | 0 | 0 |
| I1438V | ROC | N/A | No | NA | NA | NA | NA | NA | NA | NA | NA | NA | NA | NA | 0 | 0 | 0 | 0 |
| K1439R | ROC | N/A | No | NA | NA | NA | NA | NA | NA | NA | NA | NA | NA | NA | 0 | 0 | 0 | 0 |
| A1440P | ROC | VUS | Yes | NA | NA | NA | NA | NA | NA | NA | NA | NA | NA | NA | 0 | 0 | 0 | 0 |
| R1441C | ROC | P | Yes | 40310434:C:T | 6 | 16500 | 3.64E-04 | 0 | 15116 | 0 | 16 | 15720 | 1.02E-03 | 40704236:C:T | 42 | 7.19E-04 | 0 | 0 |
| R1441G | ROC | P | Yes | 4031:0434:C:G | 6 | 16500 | 3.64E-04 | 0 | 15116 | 0 |  |  |  | 40704236:C:G | 6 | 1.03E-04 | 0 | 0 |
| R1441H | ROC | P | Yes | 40310435:G:A | 1 | 16496 | 6.06E-05 | 0 | 15110 | 0 |  |  |  | 40704237:G:A | 5 | 8.56E-05 | 0 | 0 |
| R1441S | ROC | P | Yes | NA | NA | NA | NA | NA | NA | NA | NA | NA | NA | NA | 0 | 0 | 0 | 0 |
| R1441R | ROC | LB | - | only R1441G/C/H | NA | NA | NA | NA | NA | NA | NA | NA | NA | NA | 0 | 0 | 0 | 0 |
| A1442P | ROC | P | Yes | NA | NA | NA | NA | NA | NA | NA | NA | NA | NA | NA | 0 | 0 | 0 | 0 |
| A1442S | ROC | N/A | No | NA | NA | NA | NA | NA | NA | NA | NA | NA | NA | NA | 0 | 0 | 0 | 0 |
| A1442T | ROC | N/A | No | NA | NA | NA | NA | NA | NA | NA | NA | NA | NA | NA | 0 | 0 | 0 | 0 |
| S1443A | ROC | N/A | No | NA | NA | NA | NA | NA | NA | NA | NA | NA | NA | 40704242:T:G | 2 | 3.42E-05 | 0 | 0 |
| S1443F | ROC | N/A | No | NA | NA | NA | NA | NA | NA | NA | NA | NA | NA | NA | 0 | 0 | 0 | 0 |
| S1445F | ROC | VUS | No | NA | NA | NA | NA | NA | NA | NA | NA | NA | NA | NA | 0 | 0 | 0 | 0 |
| S1445C | ROC | VUS | No | NA | NA | NA | NA | NA | NA | NA | NA | NA | NA | NA | 0 | 0 | 0 | 0 |
| P1446L | ROC | VUS | No | NA | NA | NA | NA | NA | NA | NA | NA | NA | NA | 40704252:C:T | 5 | 8.56E-05 | 0 | 0 |
| V1447M | ROC | P | Yes | only V1447A | NA | NA | NA | NA | NA | NA | NA | NA | NA | NA | 0 | 0 | 0 | 0 |
| V1447G | ROC | N/A | Yes | only V1447A |  |  |  |  |  |  |  |  |  | 40704255:T:G | 1 | 1.71E-05 | 0 | 0 |
| V1447E | ROC | N/A | No | only V1447A | NA | NA | NA | NA | NA | NA | NA | NA | NA | NA | 0 | 0 | 0 | 0 |
| V1447L | ROC | N/A | Yes | only V1447A | NA | NA | NA | NA | NA | NA | NA | NA | NA | NA | 0 | 0 | 0 | 0 |
| V1450I | ROC | VUS | No | 40310461:G:A |  |  |  |  |  |  | 1 | 15720 | 6.36E-05 | 40704263:G:A | 5 | 8.56E-05 | 0 | 0 |
| G1451S | ROC | VUS | No | NA | NA | NA | NA | NA | NA | NA | NA | NA | NA | NA | 0 | 0 | 0 | 0 |
| R1462H | ROC | N/A | No | 40310498:G:A | 0 | 16498 | 0 | 2 | 15118 | 1.32E-04 |  |  |  | NA | 0 | 0 | 0 | 0 |
| A1464G | ROC | VUS | No | NA | NA | NA | NA | NA | NA | NA | NA | NA | NA | 40704306:C:G | 1 | 1.71E-05 | 0 | 0 |
| K1468E | ROC | VUS | No | 40310515:A:G | 0 | 16500 | 0 | 1 | 15114 | 6.62E-05 |  |  |  | NA | 0 | 0 | 0 | 0 |
| P1480L | ROC | VUS | No | NA | NA | NA | NA | NA | NA | NA | NA | NA | NA | NA | 0 | 0 | 0 | 0 |
| A1481T | ROC | VUS | No | NA | NA | NA | NA | NA | NA | NA | NA | NA | NA | NA | 0 | 0 | 0 | 0 |
| R1483Q | ROC | VUS | No | 40310561:G:A |  |  |  |  |  |  | 1 | 15720 | 6.36E-05 | 40704363:G:A | 1 | 1.71E-05 | 0 | 0 |
| A1490V | ROC | VUS | No | NA | NA | NA | NA | NA | NA | NA | NA | NA | NA | NA | 0 | 0 | 0 | 0 |
| E1492K | ROC | VUS | Yes | NA | NA | NA | NA | NA | NA | NA | NA | NA | NA | NA | 0 | 0 | 0 | 0 |
| R1501W | ROC | VUS | No | NA | NA | NA | NA | NA | NA | NA | NA | NA | NA | 40704416:C:T | 1 | 1.71E-05 | 0 | 0 |
| S1508G | ROC | VUS | No | NA | NA | NA | NA | NA | NA | NA | NA | NA | NA | NA | 0 | 0 | 0 | 0 |
| S1508R | ROC | N/A | No | NA | NA | NA | NA | NA | NA | NA | NA | NA | NA | NA | 0 | 0 | 0 | 0 |
| R1514Q | COR-A | B | No | 40313976:G:A | 163 | 16490 | 9.88E-03 | 122 | 15114 | 8.07E-03 | 141 | 15720 | 8.97E-03 | 40707778:G:A | 426 | 7.29E-03 | 26 | 4.52E-03 |
| R1514G | COR-A | N/A | No | only R1514Q | NA | NA | NA | NA | NA | NA | NA | NA | NA | NA | 0 | 0 | 0 | 0 |
| G1520A | COR-A | VUS | No | NA | NA | NA | NA | NA | NA | NA | NA | NA | NA | NA | 0 | 0 | 0 | 0 |
| Y1527Y | COR-A | LB | - - | NA | NA | NA | NA | NA | NA | NA | NA | NA | NA | NA | 0 | 0 | 0 | 0 |
| R1538H | COR-A | N/A | No | 40314048:G:A | 0 | 16498 | 0 | 1 | 15116 | 6.62E-05 |  |  |  | NA | 0 | 0 | 0 | 0 |
| R1538C | COR-A | N/A | No | only R1538H |  |  |  |  |  |  |  |  |  | 40707849:C:T | 2 | 3.42E-05 | 0 | 0 |
| V1541M | COR-A | VUS | No | NA | NA | NA | NA | NA | NA | NA | NA | NA | NA | NA | 0 | 0 | 0 | 0 |
| P1542S | COR-A | N/A | No | 40314059:C:T | 461 | 16496 | 0.0279 | 465 | 15118 | 0.0308 | 441 | 15720 | 0.0281 | 40707861:C:T | 1423 | 2.44E-02 | 124 | 2.16E-02 |
| I1548V | COR-A | VUS | No | NA | NA | NA | NA | NA | NA | NA | NA | NA | NA | NA | 0 | 0 | 0 | 0 |
| R1550Q | COR-A | N/A | No | 40314084:G:A | 0 | 16502 | 0 | 1 | 15116 | 6.62E-05 | 2 | 15720 | 1.27e- | 40707886:G:A | 5 | 8.56E-05 | 0 | 0 |
| L1569I | COR-A | N/A | No | NA | NA | NA | NA | NA | NA | NA | NA | NA | NA | NA | 0 | 0 | 0 | 0 |
| V1573A | COR-A | N/A | No | NA | NA | NA | NA | NA | NA | NA | NA | NA | NA | NA | 0 | 0 | 0 | 0 |
| A1589S | COR-A | VUS | No | 40315238:G:T | 1 | 16500 | 6.06E-05 | 0 | 15116 | 0 |  |  |  | NA | 0 | 0 | 0 | 0 |
| V1613A | COR-A | VUS | No | NA | NA | NA | NA | NA | NA | NA | NA | NA | NA | NA | 0 | 0 | 0 | 0 |
| V1615M | COR-A | VUS | No | NA | NA | NA | NA | NA | NA | NA | NA | NA | NA | NA | 0 | 0 | 0 | 0 |
| E1616K | COR-A | VUS | No | NA | NA | NA | NA | NA | NA | NA | NA | NA | NA | NA | 0 | 0 | 0 | 0 |
| P1619L | COR-A | VUS | No | NA | NA | NA | NA | NA | NA | NA | NA | NA | NA | NA | 0 | 0 | 0 | 0 |
| K1620R | COR-A | VUS | No | NA | NA | NA | NA | NA | NA | NA | NA | NA | NA | NA | 0 | 0 | 0 | 0 |
| H1621R | COR-A | VUS | No | NA | NA | NA | NA | NA | NA | NA | NA | NA | NA | NA | 0 | 0 | 0 | 0 |
| H1621Q | COR-A | VUS | No | NA | NA | NA | NA | NA | NA | NA | NA | NA | NA | NA | 0 | 0 | 0 | 0 |
| H1621L | COR-A | N/A | No | NA | NA | NA | NA | NA | NA | NA | NA | NA | NA | NA | 0 | 0 | 0 | 0 |
| P1622L | COR-A | VUS | Yes | NA | NA | NA | NA | NA | NA | NA | NA | NA | NA | NA | 0 | 0 | 0 | 0 |
| P1622T | COR-A | VUS | Yes | NA | NA | NA | NA | NA | NA | NA | NA | NA | NA | NA | 0 | 0 | 0 | 0 |
| K1623E | COR-A | VUS | No | NA | NA | NA | NA | NA | NA | NA | NA | NA | NA | NA | 0 | 0 | 0 | 0 |
| I1626V | COR-A | VUS | No | NA | NA | NA | NA | NA | NA | NA | NA | NA | NA | 40713838:A:G | 1 | 1.71E-05 | 0 | 0 |
| S1627T | COR-A | VUS | No | 40320039:T:A | 2 | 16498 | 1.21E-04 | 1 | 15116 | 6.62E-05 |  |  |  | NA | 0 | 0 | 0 | 0 |
| R1628P | COR-A | N/A | No | 40320043:G:C | 8 | 16460 | 4.86E-04 | 5 | 15110 | 3.31E-04 | 4 | 15698 | 2.55E-04 | 40713845:G:C | 42 | 7.19E-04 | 0 | 0 |
| R1628C | COR-A | VUS | No | 40320042:C:T | 1 | 16500 | 6.06E-05 | 0 | 15118 | 0 |  |  |  | 40713844:C:T | 2 | 3.42E-05 | 0 | 0 |
| R1628H | COR-A | N/A | No | 40320043:G:A | 0 | 16460 | 0 | 1 | 15110 | 6.62E-05 | 1 | 15698 | 6.37E-05 | 40713845:G:A | 4 | 6.85E-05 | 1 | 1.74E-04 |
| Y1645S | COR-A | VUS | No | only T1645C | NA | NA | NA | NA | NA | NA | NA | NA | NA | NA | 0 | 0 | 0 | 0 |
| M1646T | COR-A | N/A | No | 40320097:T:C | 346 | 16494 | 0.021 | 252 | 15114 | 0.0167 | 326 | 15718 | 0.0207 | 40713899:T:C | 1024 | 1.75E-02 | 137 | 2.38E-02 |
| Q1648R | COR-A | VUS | No | 40320103:A:G | 4 | 16494 | 2.43E-04 | 4 | 15118 | 2.65E-04 | 2 | 15718 | 1.27E-04 | 40713905:A:G | 4 | 6.85E-05 | 0 | 0 |
| Y1649S | COR-A | VUS | No | NA | NA | NA | NA | NA | NA | NA | NA | NA | NA | NA | 0 | 0 | 0 | 0 |
| E1654C | COR-A | N/A | No | NA | NA | NA | NA | NA | NA | NA | NA | NA | NA | NA | 0 | 0 | 0 | 0 |
| Q1657* | COR-A | N/A | No | NA | NA | NA | NA | NA | NA | NA | NA | NA | NA | NA | 0 | 0 | 0 | 0 |
| I1658F | COR-A | LP | NA | NA | NA | NA | NA | NA | NA | NA | NA | NA | NA | NA | 0 | 0 | 0 | 0 |
| P1661T | COR-A | VUS | No | 40320141:C:A | 0 | 16478 | 0 | 1 | 15110 | 6.62E-05 |  |  |  | 40713943:C:A | 2 | 3.42E-05 | 0 | 0 |
| R1677S | COR-B | VUS | No | 40321049:G:T | 1 | 16496 | 6.06E-05 | 0 | 15116 | 0 |  |  |  | 40714851:G:T | 2 | 3.42E-05 | 0 | 0 |
| I1691T | COR-B | VUS | No | NA | NA | NA | NA | NA | NA | NA | NA | NA | NA | NA | 0 | 0 | 0 | 0 |
| R1693Q | COR-B | VUS | No | NA | NA | NA | NA | NA | NA | NA | NA | NA | NA | NA | 0 | 0 | 0 | 0 |
| P1698L | COR-B | N/A | No | 40321111:C:T |  |  |  |  |  |  | 1 | 15720 | 6.36E-05 | 40714913:C:T | 1 | 1.71E-05 | 0 | 0 |
| Y1699C | COR-B | P | Yes | 40321114:A:G | 1 | 16466 | 6.07E-05 | 0 | 15098 | 0 |  |  |  | NA | 0 | 0 | 0 | 0 |
| F1700L | COR-B | P | Yes | NA | NA | NA | NA | NA | NA | NA | NA | NA | NA | NA | 0 | 0 | 0 | 0 |
| M1702T | COR-B | N/A | Yes | 40321123:T:C |  |  |  |  |  |  | 1 | 15720 | 6.36E-05 | 40714925:T:C | 3 | 5.13E-05 | 0 | 0 |
| M1702V | COR-B | N/A | No | 40321122:A:G | 0 | 16484 | 0 | 1 | 15116 | 6.62E-05 |  |  |  | NA | 0 | 0 | 0 | 0 |
| R1707K | COR-B | VUS | No | NA | NA | NA | NA | NA | NA | NA | NA | NA | NA | NA | 0 | 0 | 0 | 0 |
| L1712F | COR-B | VUS | Yes | NA | NA | NA | NA | NA | NA | NA | NA | NA | NA | NA | 0 | 0 | 0 | 0 |
| R1725Q | COR-B | VUS | No | 40322038:G:A |  |  |  |  |  |  | 1 | 15720 | 6.36E-05 | 40715840:G:A | 1 | 1.71E-05 | 0 | 0 |
| R1728H | COR-B | LP | Yes | only R1728L | NA | NA | NA | NA | NA | NA | NA | NA | NA | NA | 0 | 0 | 0 | 0 |
| R1728L | COR-B | N/A | Yes | 40322047:G:T | 1 | 16494 | 6.06E-05 | 2 | 15114 | 1.32E-04 | 2 | 15720 | 1.27E-04 | 40715849:G:T | 10 | 1.71E-04 | 0 | 0 |
| S1752C | COR-B | VUS | No | NA | NA | NA | NA | NA | NA | NA | NA | NA | NA | NA | 0 | 0 | 0 | 0 |
| D1756G | COR-B | VUS | No | NA | NA | NA | NA | NA | NA | NA | NA | NA | NA | NA | 0 | 0 | 0 | 0 |
| D1756Y | COR-B | VUS | No | NA | NA | NA | NA | NA | NA | NA | NA | NA | NA | 40715932:G:T | 1 | 1.71E-05 | 0 | 0 |
| H1758P | COR-B | VUS | No | NA | NA | NA | NA | NA | NA | NA | NA | NA | NA | NA | 0 | 0 | 0 | 0 |
| P1759L | COR-B | N/A | No | NA | NA | NA | NA | NA | NA | NA | NA | NA | NA | NA | 0 | 0 | 0 | 0 |
| S1761R | COR-B | LP | Yes | 40322145:A:C | 2 | 16488 | 1.21E-04 | 0 | 15110 | 0 |  |  |  | 40715947:A:C | 1 | 1.71E-05 | 0 | 0 |
| S1761N | COR-B | N/A | No | only S1761R |  |  |  |  |  |  |  |  |  | NA | 0 | 0 | 0 | 0 |
| F1762S | COR-B | N/A | No | NA | NA | NA | NA | NA | NA | NA | NA | NA | NA | 40715951:T:C | 1 | 1.71E-05 | 0 | 0 |
| R1771T | COR-B | VUS | No | NA | NA | NA | NA | NA | NA | NA | NA | NA | NA | NA | 0 | 0 | 0 | 0 |
| C1774Y | COR-B | VUS | No | NA | NA | NA | NA | NA | NA | NA | NA | NA | NA | NA | 0 | 0 | 0 | 0 |
| L1795F | COR-B | P | Yes | 40322386:G:T | 9 | 16502 | 5.45E-04 | 0 | 15116 | 0 |  |  |  | 40716188:G:T | 11 | 1.88E-04 | 0 | 0 |
| E1797D | COR-B | N/A | No | NA | NA | NA | NA | NA | NA | NA | NA | NA | NA | NA | 0 | 0 | 0 | 0 |
| T1806I | COR-B | VUS | No | NA | NA | NA | NA | NA | NA | NA | NA | NA | NA | NA | 0 | 0 | 0 | 0 |
| G1819R | COR-B | VUS | No | NA | NA | NA | NA | NA | NA | NA | NA | NA | NA | NA | 0 | 0 | 0 | 0 |
| Q1823H | COR-B | N/A | No | only Q1823K | NA | NA | NA | NA | NA | NA | NA | NA | NA | NA | 0 | 0 | 0 | 0 |
| Q1823K | COR-B | VUS | No | 40322468:C:A | 2 | 16484 | 1.21E-04 | 0 | 15110 | 0 |  |  |  | 40716270:C:A | 3 | 5.13E-05 | 0 | 0 |
| L1827F | COR-B | N/A | No | NA | NA | NA | NA | NA | NA | NA | NA | NA | NA | NA | 0 | 0 | 0 | 0 |
| I1852V | COR-B | N/A | No | 40323204:A:G | 1 | 16496 | 6.06E-05 | 1 | 15116 | 6.62E-05 |  |  |  | NA | 0 | 0 | 0 | 0 |
| P1865R | COR-B | N/A | No | 40323244:C:G | 1 | 16500 | 6.06E-05 | 0 | 15118 | 0 | 1 | 15720 | 6.36E-05 | 40717046:C:G | 1 | 1.71E-05 | 1 | 1.74E-04 |
| M1869V | COR-B | VUS | No | only M1869T |  |  |  |  |  |  |  |  |  | NA | 0 | 0 | 0 | 0 |
| M1869T | COR-B | VUS | No | 40323256:T:C | 2 | 16498 | 1.21E-04 | 5 | 15116 | 3.31E-04 | 6 | 15720 | 3.82E-04 | 40717058:T:C | 26 | 4.45E-04 | 1 | 1.74E-04 |
| D1887N | KIN | VUS | Yes | NA | NA | NA | NA | NA | NA | NA | NA | NA | NA | NA | 0 | 0 | 0 | 0 |
| G1891A | KIN | VUS | Yes | NA | NA | NA | NA | NA | NA | NA | NA | NA | NA | NA | 0 | 0 | 0 | 0 |
| G1900E | KIN | N/A | Yes | NA | NA | NA | NA | NA | NA | NA | NA | NA | NA | NA | 0 | 0 | 0 | 0 |
| L1914I | KIN | VUS | Yes | NA | NA | NA | NA | NA | NA | NA | NA | NA | NA | NA | 0 | 0 | 0 | 0 |
| R1941H | KIN | VUS | No | 40335031:G:A | 7 | 16496 | 4.24E-04 | 1 | 15116 | 6.62E-05 | 2 | 15720 | 1.27E-04 | 40728833:G:A | 8 | 1.37E-04 | 1 | 1.74E-04 |
| S1954F^1^ | KIN | LP | No | NA | NA | NA | NA | NA | NA | NA | NA | NA | NA | NA | 0 | 0 | 0 | 0 |
| R1957G | KIN | N/A | No | only R1757C | NA | NA | NA | NA | NA | NA | NA | NA | NA | 40728880:C:G | 1 | 1.71E-05 | 0 | 0 |
| Q1961R | KIN | VUS | No | NA | NA | NA | NA | NA | NA | NA | NA | NA | NA | 40728893:A:G | 2 | 3.42E-05 | 0 | 0 |
| I1991V | KIN | VUS | No | NA | NA | NA | NA | NA | NA | NA | NA | NA | NA | NA | 0 | 0 | 0 | 0 |
| Y2006H | KIN | VUS | No | NA | NA | NA | NA | NA | NA | NA | NA | NA | NA | NA | 0 | 0 | 0 | 0 |
| A2010T | KIN | VUS | No | NA | NA | NA | NA | NA | NA | NA | NA | NA | NA | NA | 0 | 0 | 0 | 0 |
| I2012T | KIN | VUS | No | NA | NA | NA | NA | NA | NA | NA | NA | NA | NA | NA | 0 | 0 | 0 | 0 |
| Y2018Y | KIN | LB | - | NA | NA | NA | NA | NA | NA | NA | NA | NA | NA | 40734201:C:T | 7 | 1.20E-04 | 3 | 5.22E-04 |
| G2019S | KIN | P | Yes | 40340400:G:A | 89 | 16490 | 5.40E-03 | 7 | 15116 | 4.63E-04 | 186 | 15720 | 0.0118 | 40734202:G:A | 603 | 1.03E-02 | 314 | 5.46E-02 |
| I2020T | KIN | P | Yes | 40340404:T:C | 2 | 16498 | 1.21E-04 | 0 | 15118 | 0 |  |  |  | NA | 0 | 0 | 0 | 0 |
| I2020L | KIN | LP | No | NA | NA | NA | NA | NA | NA | NA | NA | NA | NA | NA | 0 | 0 | 0 | 0 |
| I2020S | KIN | LP | Yes | NA | NA | NA | NA | NA | NA | NA | NA | NA | NA | NA | 0 | 0 | 0 | 0 |
| T2031S | KIN | LP | Yes | NA | NA | NA | NA | NA | NA | NA | NA | NA | NA | NA | 0 | 0 | 0 | 0 |
| L2063* | KIN | VUS | No | 40346826:ACTACT:A | 2 | 16478 | 1.21E-04 | 5 | 15098 | 3.31E-04 | 2 | 15720 | 1.27E-04 | NA | 0 | 0 | 0 | 0 |
| G2071D | KIN | N/A | No | only G2071S | NA | NA | NA | NA | NA | NA | NA | NA | NA | NA | 0 | 0 | 0 | 0 |
| V2074I | KIN | VUS | No | 40346863:G:A | 0 | 16492 | 0 | 1 | 15116 | 6.62E-05 | 1 | 15720 | 6.36E-05 | 40740665:G:A | 1 | 1.71E-05 | 0 | 0 |
| N2081D | KIN | N/A | Yes | 40346884:A:G | 376 | 16488 | 0.0228 | 274 | 15108 | 0.0181 | 455 | 15718 | 0.0289 | 40740686:A:G | 1544 | 2.64E-02 | 303 | 5.27E-02 |
| D2084N | KIN | N/A | No | 40346893:G:A | 0 | 16486 | 0 | 1 | 15104 | 6.62E-05 |  |  |  | NA | 0 | 0 | 0 | 0 |
| E2108K | KIN | VUS | No | NA | NA | NA | NA | NA | NA | NA | NA | NA | NA | NA | 0 | 0 | 0 | 0 |
| N2133S | KIN | VUS | No | NA | NA | NA | NA | NA | NA | NA | NA | NA | NA | NA | 0 | 0 | 0 | 0 |
| C2139S | KIN | VUS | No | NA | NA | NA | NA | NA | NA | NA | NA | NA | NA | NA | 0 | 0 | 0 | 0 |
| T2141M | KIN | N/A | No | NA | NA | NA | NA | NA | NA | NA | NA | NA | NA | NA | 0 | 0 | 0 | 0 |
| R2143H | WD40 | N/A | No | 40351585:G:A | 0 | 16500 | 0 | 4 | 15116 | 2.65E-04 |  |  |  | 40745387:G:A | 13 | 2.23E-04 | 0 | 0 |
| R2143M | WD40 | N/A | No | only R2143H | NA | NA | NA | NA | NA | NA | NA | NA | NA | NA | 0 | 0 | 0 | 0 |
| V2150I | WD40 | VUS | No | NA | NA | NA | NA | NA | NA | NA | NA | NA | NA | 40745407:G:A | 1 | 1.71E-05 | 0 | 0 |
| W2168L | WD40 | VUS | No | only W2168C | NA | NA | NA | NA | NA | NA | NA | NA | NA | NA | 0 | 0 | 0 | 0 |
| D2175H | WD40 | VUS | No | 40351680:G:C | 1 | 16504 | 6.06E-05 | 0 | 15118 | 0 |  |  |  | NA | 0 | 0 | 0 | 0 |
| Y2189C | WD40 | VUS | No | 40351723:A:G | 4 | 16504 | 2.42E-04 | 2 | 15118 | 1.32E-04 | 4 | 15720 | 2.54E-04 | 40745525:A:G | 26 | 4.45E-04 | 2 | 3.48E-04 |
| S2213C | WD40 | N/A | No | NA | NA | NA | NA | NA | NA | NA | NA | NA | NA | NA | 0 | 0 | 0 | 0 |
| H2236R | WD40 | VUS | No | NA | NA | NA | NA | NA | NA | NA | NA | NA | NA | NA | 0 | 0 | 0 | 0 |
| N2251T | WD40 | VUS | No | NA | NA | NA | NA | NA | NA | NA | NA | NA | NA | 40748276:A:C | 1 | 1.71E-05 | 0 | 0 |
| K2255M | WD40 | N/A | No | 40354486:A:T | 0 | 16502 | 0 | 1 | 15118 | 6.61E-05 | 2 | 15720 | 1.27E-04 | 40748288:A:T | 4 | 6.85E-05 | 1 | 1.74E-04 |
| N2261K | WD40 | VUS | No | Only N2261S | NA | NA | NA | NA | NA | NA | NA | NA | NA | 40749929:T:A | 1 | 1.71E-05 | 0 | 0 |
| K2278E | WD40 | N/A | No | 40356176:A:G |  |  |  |  |  |  | 2 | 15714 | 1.27E-04 | 40749978:A:G | 2 | 3.42E-05 | 0 | 0 |
| G2294R | WD40 | VUS | No | 40359296:G:A | 0 | 16398 | 0 | 1 | 15084 | 6.63E-05 |  |  |  | NA | 0 | 0 | 0 | 0 |
| V2296F | WD40 | N/A | No | NA | NA | NA | NA | NA | NA | NA | NA | NA | NA | NA | 0 | 0 | 0 | 0 |
| N2308D | WD40 | VUS | No | only N2308S | NA | NA | NA | NA | NA | NA | NA | NA | NA | NA | 0 | 0 | 0 | 0 |
| T2310A | WD40 | N/A | No | 40359344:A:G | 1 | 16450 | 6.08E-05 | 2 | 15092 | 1.33E-04 | 2 | 15720 | 1.27E-04 | 40753146:A:G | 5 | 8.56E-05 | 0 | 0 |
| T2310M | WD40 | VUS | No | 40359345:C:T | 1 | 16236 | 6.16E-05 | 1 | 15064 | 6.64E-05 | 2 | 15720 | 1.27E-04 | 40753147:C:T | 5 | 8.56E-05 | 1 | 1.74E-04 |
| N2313S | WD40 | VUS | No | NA | NA | NA | NA | NA | NA | NA | NA | NA | NA | NA | 0 | 0 | 0 | 0 |
| I2323F | WD40 | VUS | No | NA | NA | NA | NA | NA | NA | NA | NA | NA | NA | NA | 0 | 0 | 0 | 0 |
| I2336V | WD40 | VUS | No | NA | NA | NA | NA | NA | NA | NA | NA | NA | NA | NA | 0 | 0 | 0 | 0 |
| S2350I | WD40 | VUS | No | NA | NA | NA | NA | NA | NA | NA | NA | NA | NA | NA | 0 | 0 | 0 | 0 |
| T2356I | WD40 | VUS | No | 40363440:C:T | 6 | 16474 | 3.64E-04 | 2 | 15110 | 1.32E-04 | 7 | 15720 | 4.45E-04 | 40757242:C:T | 19 | 3.25E-04 | 0 | 0 |
| G2385R | WD40 | N/A | Yes | 40363526:G:A | 2 | 16492 | 1.21E-04 | 1 | 15114 | 6.62E-05 |  |  |  | 40757328:G:A | 54 | 9.24E-04 | 0 | 0 |
| V2390A | WD40 | VUS | No | only V2390M | NA | NA | NA | NA | NA | NA | NA | NA | NA | 40757344:T:C | 1 | 1.71E-05 | 0 | 0 |
| V2390M | WD40 | VUS | No | 40363541:G:A | 0 | 16498 | 0 | 1 | 15118 | 6.61E-05 | 1 | 15720 | 6.36E-05 | 40757343:G:A | 3 | 5.13E-05 | 0 | 0 |
| H2391Q | WD40 | LB | No | NA | NA | NA | NA | NA | NA | NA | NA | NA | NA | NA | 0 | 0 | 0 | 0 |
| E2395K | WD40 | N/A | No | NA | NA | NA | NA | NA | NA | NA | NA | NA | NA | 40758645:G:A | 1 | 1.71E-05 | 1 | 1.74E-04 |
| M2397T | WD40 | N/A | No | 40364850:T:C | 10990 | 16482 | 0.667 | 10111 | 15088 | 0.67 | 10351 | 15714 | 0.659 | 40758652:T:C | 36692 | 6.28E-01 | 3533 | 6.14E-01 |
| T2423S | WD40 | VUS | No | NA | NA | NA | NA | NA | NA | NA | NA | NA | NA | NA | 0 | 0 | 0 | 0 |
| I2434V | WD40 | VUS | No | NA | NA | NA | NA | NA | NA | NA | NA | NA | NA | NA | 0 | 0 | 0 | 0 |
| L2436P | WD40 | N/A | No | NA | NA | NA | NA | NA | NA | NA | NA | NA | NA | NA | 0 | 0 | 0 | 0 |
| L2439I | WD40 | VUS | No | 40364975:C:A | 0 | 16502 | 0 | 2 | 15114 | 1.32E-04 | 1 | 15720 | 6.36E-05 | 40758777:C:A | 5 | 8.56E-05 | 0 | 0 |
| A2461V | WD40 | VUS | No | 40365042:C:T | 1 | 16504 | 6.06E-05 | 2 | 15116 | 1.32E-04 | 1 | 15720 | 6.36E-05 | 40758844:C:T | 6 | 1.03E-04 | 0 | 0 |
| L2463P | WD40 | VUS | No | NA | NA | NA | NA | NA | NA | NA | NA | NA | NA | NA | 0 | 0 | 0 | 0 |
| L2466H | WD40 | N/A | No | NA | NA | NA | NA | NA | NA | NA | NA | NA | NA | NA | 0 | 0 | 0 | 0 |
| K2467Q | WD40 | N/A | Yes | 40367014:A:C | 2 | 16476 | 1.21E-04 | 0 | 15102 | 0 |  |  |  | NA | 0 | 0 | 0 | 0 |
| Q2484K | WD40 | N/A | No | NA | NA | NA | NA | NA | NA | NA | NA | NA | NA | NA | 0 | 0 | 0 | 0 |
| Q2490H | WD40 | VUS | No | NA | NA | NA | NA | NA | NA | NA | NA | NA | NA | NA | 0 | 0 | 0 | 0 |
| T2494I | WD40 | VUS | No | NA | NA | NA | NA | NA | NA | NA | NA | NA | NA | NA | 0 | 0 | 0 | 0 |
| V2495I | WD40 | VUS | No | 40367664:G:A | 2 | 16422 | 1.22E-04 | 2 | 15078 | 1.33E-04 | 2 | 15720 | 1.27E-04 | 40761466:G:A | 15 | 2.57E-04 | 0 | 0 |
| T2524A | WD40 | VUS | No | NA | NA | NA | NA | NA | NA | NA | NA | NA | NA | NA | 0 | 0 | 0 | 0 |

Variants are annotated by protein domain, MDSGene/ACMG classification, and functional activation status. Allele count (AC), allele number (AN), and allele frequency (AF) are shown for PD cases and healthy controls (HC) from GP2 whole-genome sequencing (WGS) datasets, and for PD GENEration (PD cases only), including individuals of Ashkenazi Jewish (AJ) ancestry where indicated. Genomic coordinates are provided in hg19 and hg38. NA indicates data not available.

**Abbreviations:** AC, allele count; AF, allele frequency; AJ, Ashkenazi Jewish; AN, allele number; CES, clinical exome sequencing; HC, healthy controls; PD, Parkinson's disease; WGS, whole-genome sequencing; NA, not available.

**Supplementary Figure 1. Representative immunoblot analysis of LRRK2 kinase-activating variants under standardized overexpression conditions.**
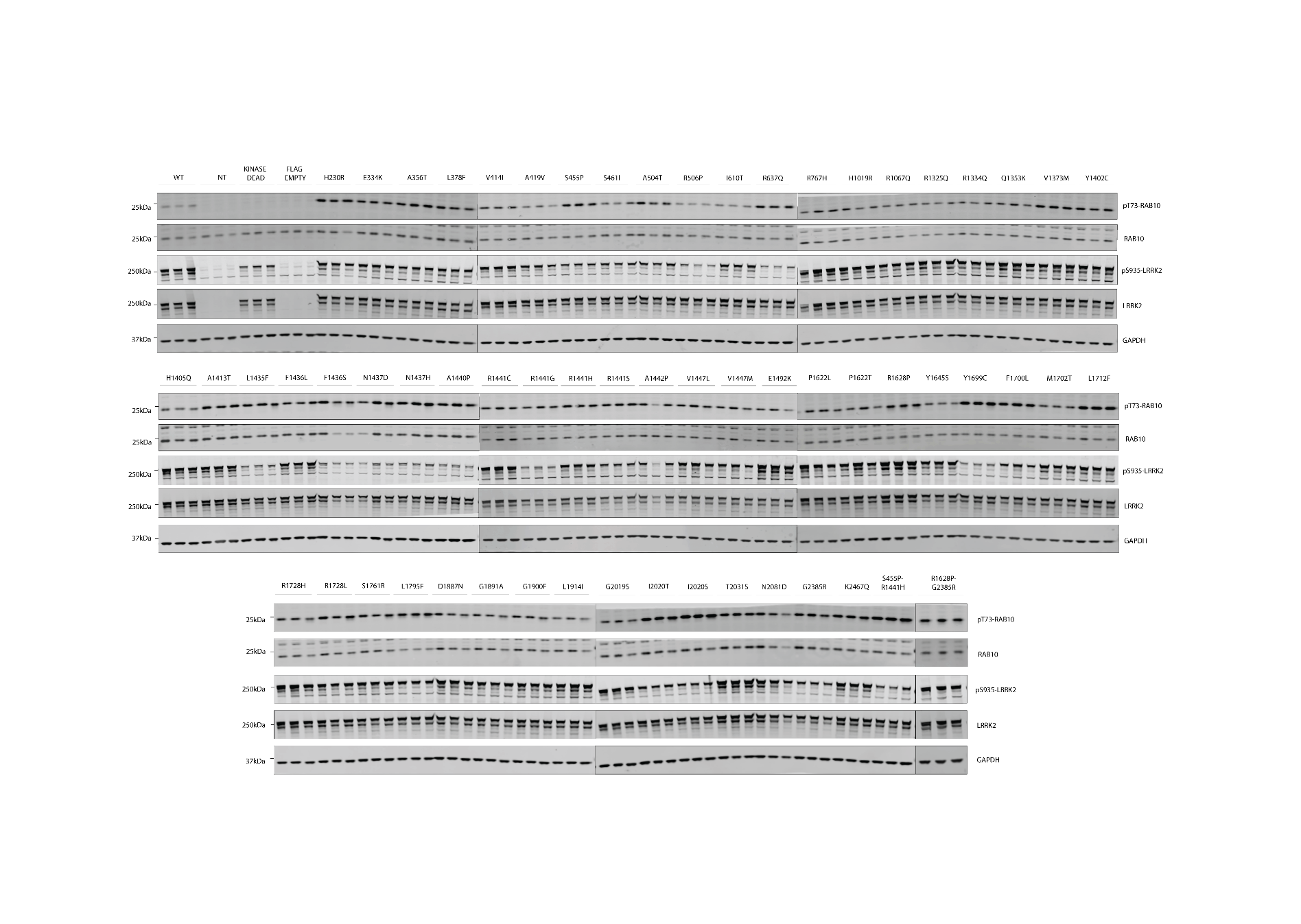

FLAG-tagged LRRK2 wild-type (WT), kinase-dead (KD; D2017A), and indicated activating variants (representing all activating LRRK2 variants identified to date across independent functional studies) were transiently expressed in HEK293 cells. Cells were lysed 24 h post-transfection and analysed by quantitative immunoblotting using the indicated antibodies. Each lane represents an independent biological replicate (separate transfections performed on different days). All variants expressed detectable LRRK2 protein, enabling normalization of pThr73-Rab10 phosphorylation to total LRRK2 levels. Quantification of pThr73-Rab10 phosphorylation is shown in Fig. 4B, whereas quantification of total LRRK2, Ser935 phosphorylation, and total Rab10 levels is provided in Supplementary Fig. 2. Several activating variants, particularly within the ROC-COR region, exhibited reduced Ser935 phosphorylation, consistent with previous reports linking increased LRRK2 kinase activity to decreased phosphorylation at 14-3-3 binding sites.

**Supplementary Figure 2: Quantification of total LRRK2, Ser935 phosphorylation, and total Rab10 levels in activating LRRK2 variants**.

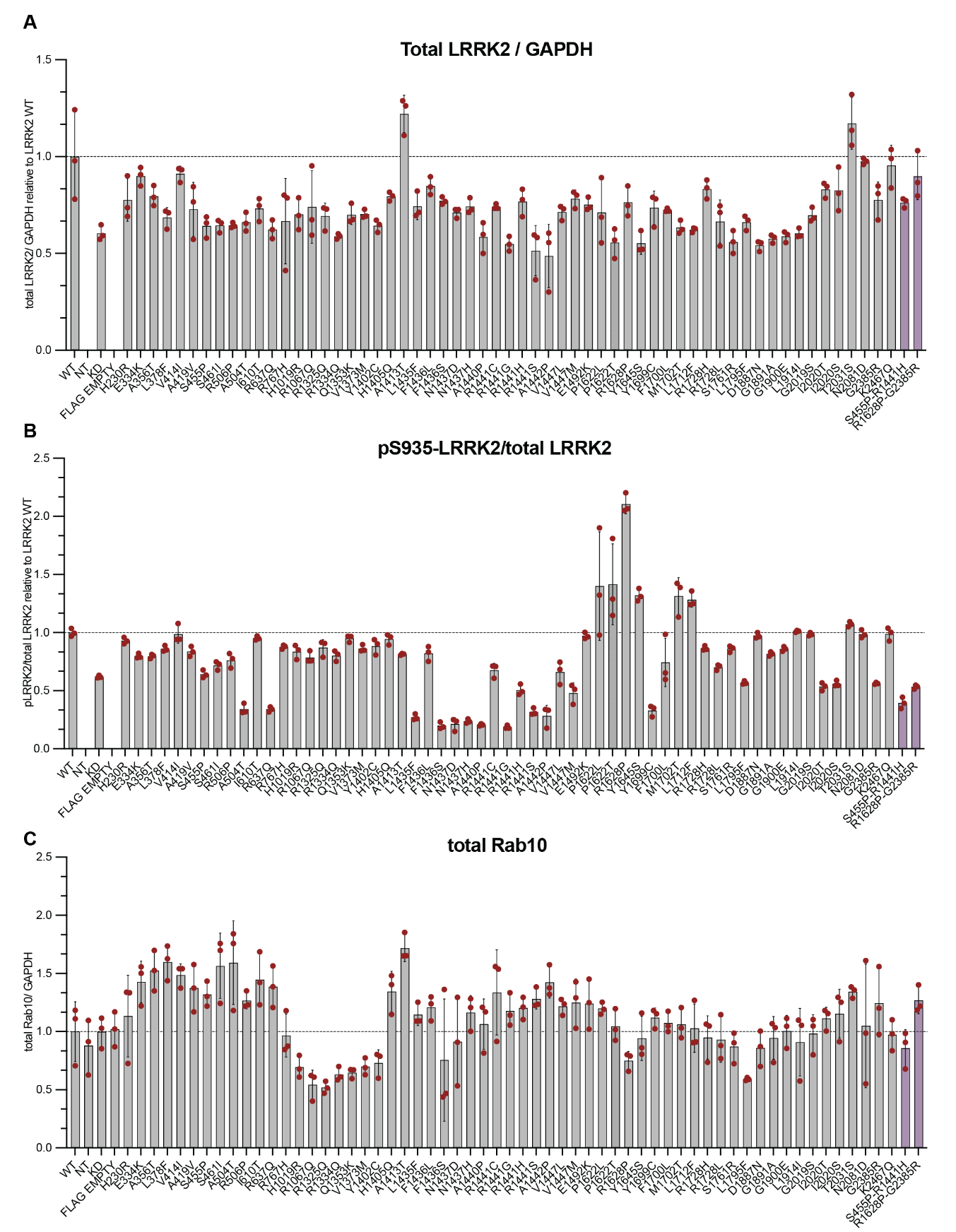

Data are presented as ratios of total LRRK2/GAPDH (A), pSer935-LRRK2/total LRRK2 (B), and total Rab10/GAPDH (C) for FLAG-tagged LRRK2 WT, KD (D2017A), and the indicated variants. Each data point represents an independent biological replicate.

**Figure 5-figure supplement 1. Dimerization, 14-3-3 binding, and evolutionary conservation of the four-part COR scaffold.**

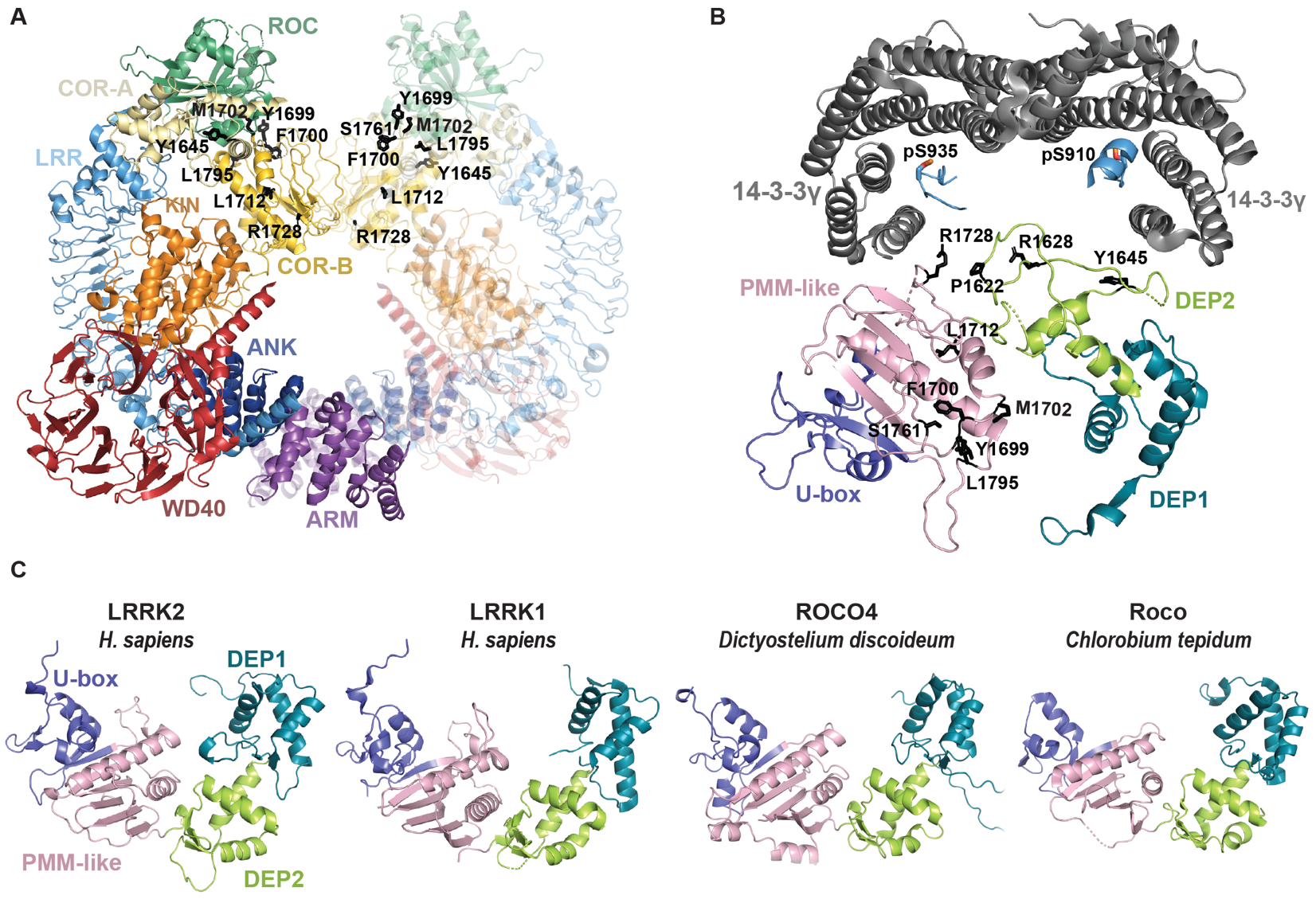

Panel **A** is based on PDB 8U8A. Panel **B** shows the 14-3-3γ-binding interface reported by Zhang and colleagues and is based on PDB 9CI3. Comparative evolutionary views additionally use human LRRK1 (PDB 8E04), Dictyostelium discoideum ROCO4 (AF3) and Chlorobaculum tepidum Roco (PDB 6HLU).

**(A)** Structure of the LRRK2 dimer with activating variants in the COR region shown as black sticks. This panel was used to assess whether activating COR variants might directly perturb the dimerization interface. Among the mapped variants, Arg1728 appears closest to the dimer interface, suggesting that direct disruption of dimerization may explain only a minority of activating substitutions. **(B)** Structure of 14-3-3γ bound to the COR region, with activating variants shown as black sticks. This panel illustrates the positions of activating variant sites relative to the 14-3-3-binding interface and supports the idea that variants affecting Pro1622, Arg1628, Tyr1645 and Arg1728 could weaken 14-3-3 binding, thereby favouring a more active LRRK2 conformation. **(C)** Structural comparison of the COR region from human LRRK2, human LRRK1, Dictyostelium discoideum Roco4, and Chlorobaculum tepidum Roco. In each case the COR region is partitioned into the four proposed subdomains DEP1, DEP2, PMM-like and U-box-like. This comparison illustrates the apparent conservation of a four-part COR scaffold from bacterial Roco proteins through early eukaryotes to human LRRK proteins.

**Figure 5-figure supplement 2. Structural comparisons and mechanistic hypotheses for the DEP1 and DEP2 subdomains.**

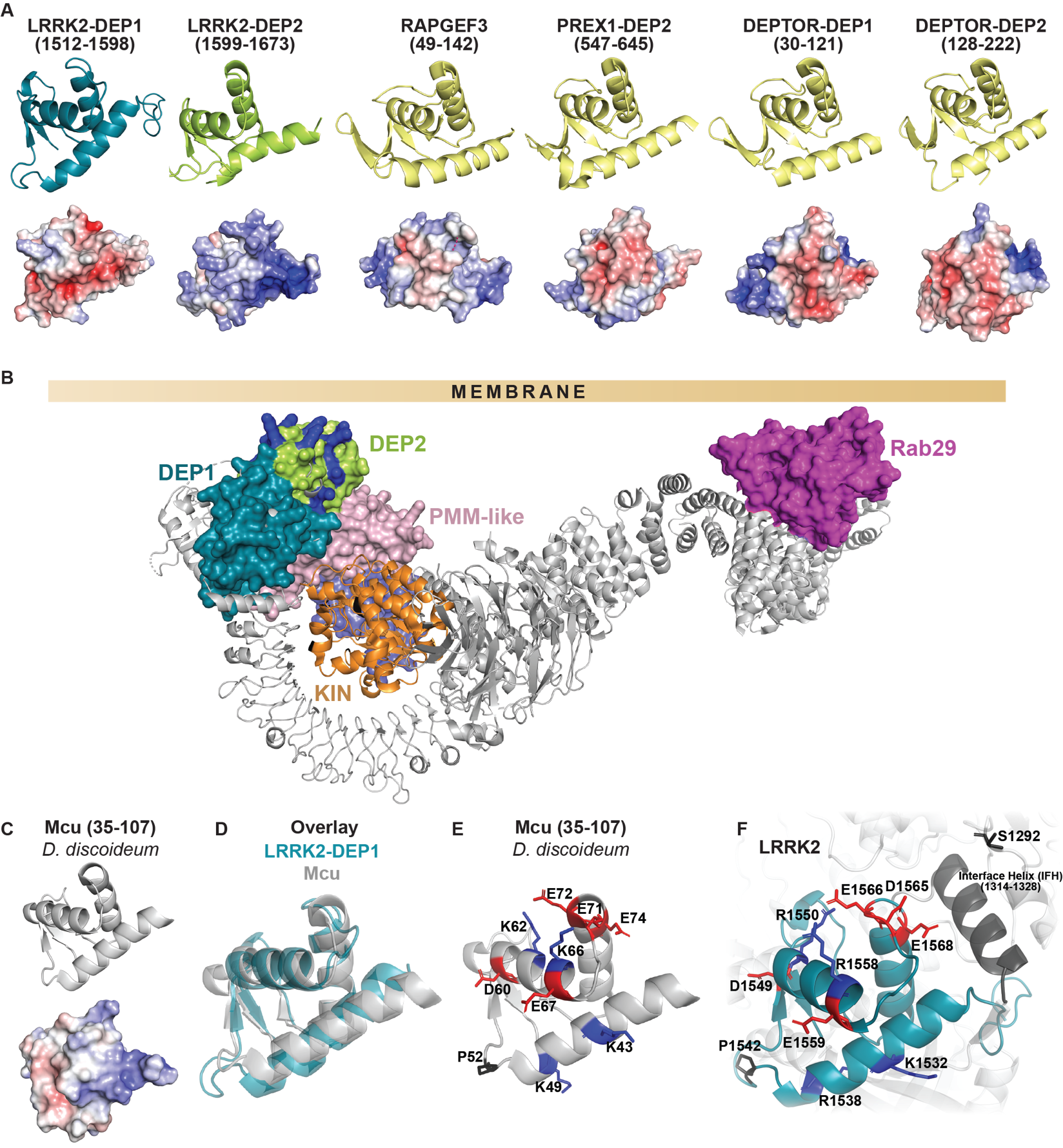
This figure uses structural comparisons with RAPGEF3 (PDB 6H7E), DEPTOR (PDB 7DKL), PREX1 (PDB 7RX9), and Mcu (Dictyostelium discoideum, PDB 5Z2H). Where indicated, AlphaFold models were also used. (A) Structural comparison of the proposed LRRK2 DEP1 and DEP2 subdomains with structurally similar DEP domains from other proteins, including RAPGEF3 (PDB 6H7E), PREX1 (PDB 7RX9), PREX2 where applicable, and DEPTOR (PDB 7DKL). Surface electrostatic representations are shown for the compared domains. This panel illustrates fold-level similarity of the LRRK2 subdomains to known DEP-domain-containing proteins and supports the interpretation of DEP1 and DEP2 as distinct structural modules. (B) Model of full-length LRRK2 recruited to the membrane by Rab29 through the armadillo domain. The overall orientation is informed by the Rab29-LRRK2 cryo-EM structure (PDB 8FO2). In this orientation, the DEP2 subdomain faces the membrane. Positively charged residues on DEP2 are highlighted (in dark blue), consistent with the possibility that they interact with negatively charged phospholipids and contribute to membrane association. This panel supports the hypothesis that DEP2 may act as a second membrane-binding element in LRRK2 in addition to Rab-mediated recruitment. **(D)** Structural overlay of the LRRK2 DEP1 subdomain with the Dictyostelium discoideum mitochondrial calcium transporter Mcu (PDB 5Z2H). This comparison identifies structural similarity between DEP1 and MCU (PMID: 32258880). **(E)** Structure of MCU with key charged residues highlighted. **(F)** Structure of the LRRK2 DEP1 subdomain with the corresponding charged residues highlighted. The conservation of these residues supports the hypothesis that DEP1 may contain a metal-responsive loop analogous to the calcium-binding feature of MCU, raising the possibility that metal binding could influence COR conformation and thereby regulate LRRK2 activity.

**Figure 5-figure supplement 3. Structural analogies of the PMM-like and U-box-like subdomains.**

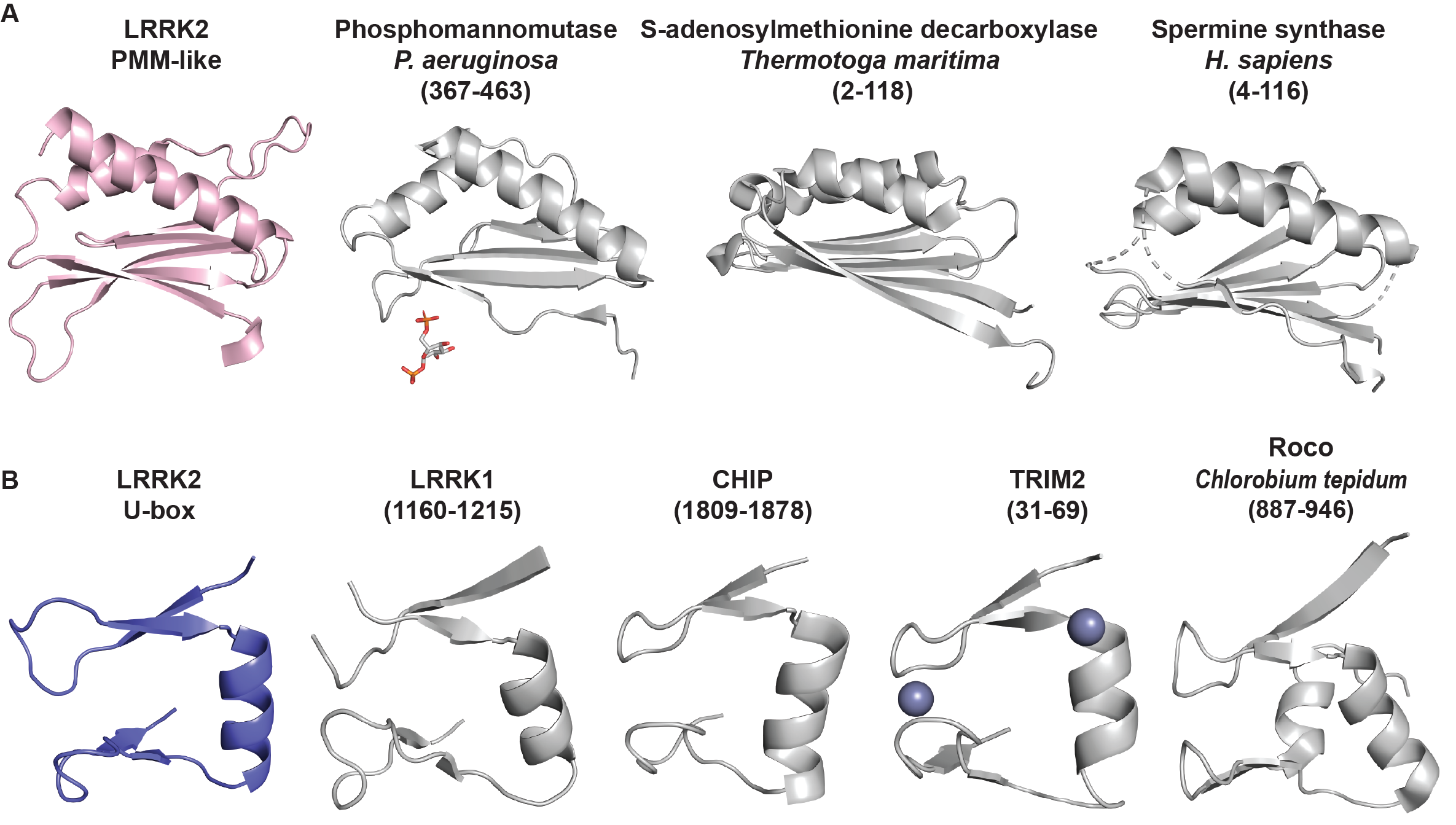

This figure uses structural comparisons with Pseudomonas aeruginosa phosphomannomutase (PDB 2FKM), Thermococcus kodakarensis phosphomannomutase/phosphoglucomutase-like protein (PDB 9IX8), Thermotoga maritima S-adenosylmethionine decarboxylase (PDB 1TMI), CHIP U-box (PDB 9DRY), TRIM2 RING (PDB 7ZJ3), human LRRK1 (PDB 8E04), and Chlorobaculum tepidum Roco (PDB 6HLU). **(A)** Isolated PMM-like subdomain of LRRK2 compared with structurally similar proteins or protein subdomains from Pseudomonas aeruginosa phosphomannomutase (PDB 2FKM), Thermococcus kodakarensis phosphomannomutase/phosphoglucomutase-like protein (PDB 9IX8), Thermotoga maritima S-adenosylmethionine decarboxylase (PDB 1TMI), and human spermine synthase. This comparison supports the interpretation of this region as a PMM-like module and raises the possibility that it may function as a small-molecule-responsive regulatory element. Notably, this is also the proposed COR subdomain that contains the highest number of activating patient variants. **(B)** Isolated U-box-like subdomain of LRRK2 compared with the corresponding region of LRRK1 (PDB 8E04), the U-box domain of CHIP (PDB 9DRY), the RING domain of TRIM2 (PDB 7ZJ3), and the corresponding region of Chlorobaculum tepidum Roco (PDB 6HLU). This panel illustrates structural similarity between the LRRK2 subdomain and U-box/RING-like folds, while also showing conservation of the underlying scaffold in bacterial Roco. These comparisons support the idea that the COR region includes an ancient structural module with similarity to later eukaryotic ubiquitin-related folds, without implying that the LRRK2 region is a bona fide canonical U-box domain.

**Supplementary Figure 3. LRRK2 dependent Rab10 phosphorylation in peripheral blood neutrophils of LRRK2 variant carriers and healthy controls.**

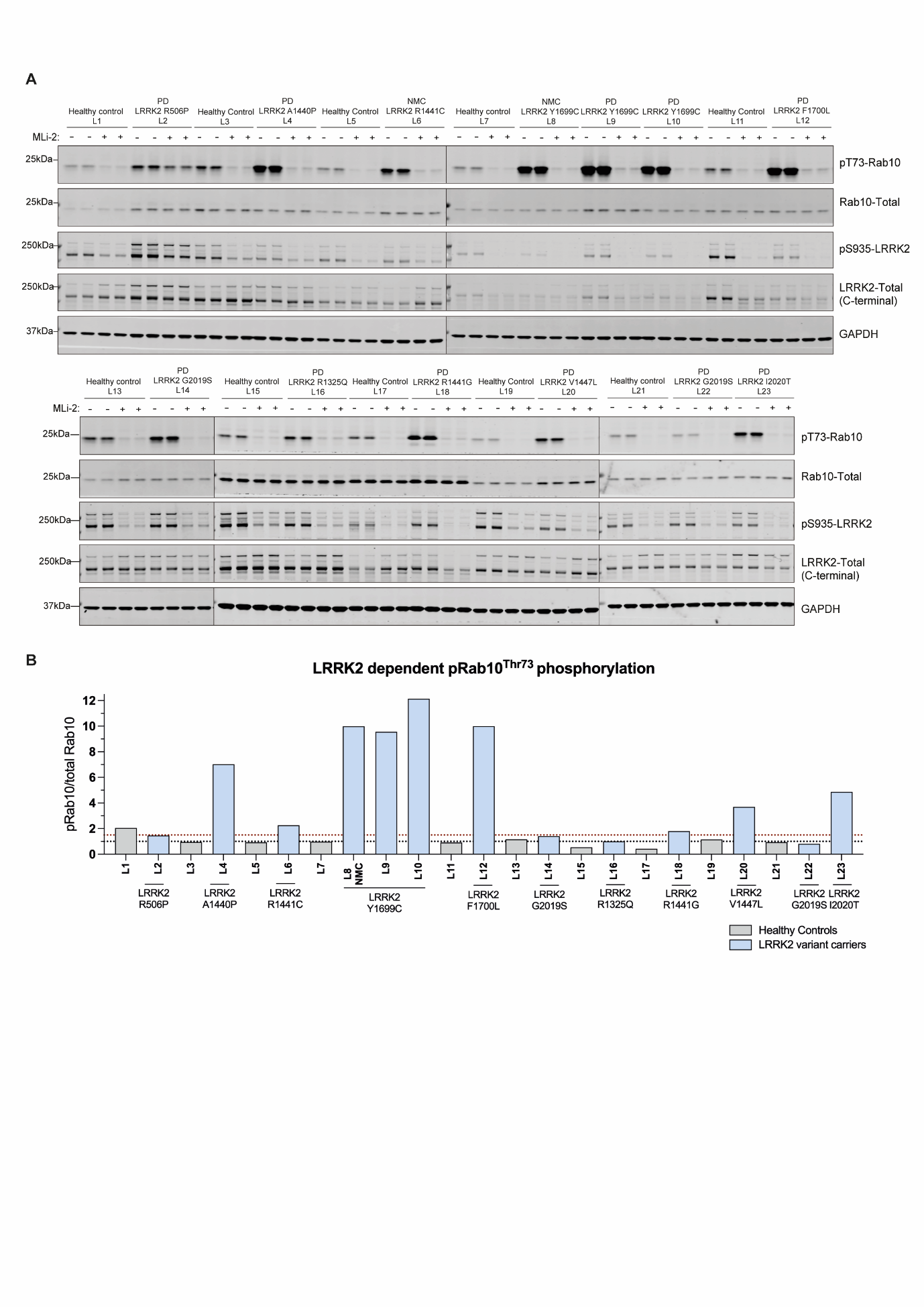

(A) and (B) Neutrophils were isolated from 10 mL of fresh peripheral blood from each participant and, where indicated, treated with 200 nM MLi-2 or vehicle control for 30 min prior to lysis. Equal amounts of protein (10 μg) were resolved by SDS-PAGE and analysed by immunoblotting using the indicated antibodies. Membranes were imaged using a LI-COR Odyssey CLx system. Quantified pRab10/total Rab10 ratios were normalized to the mean value of healthy controls (set to 1.0). Accordingly, a value of 1.5 corresponds to a 1.5-fold increase in LRRK2 kinase activity relative to healthy controls.

**Supplementary Figure 4. LRRK2 kinase activity of patient-derived variants assessed in HEK293 overexpression assays.**

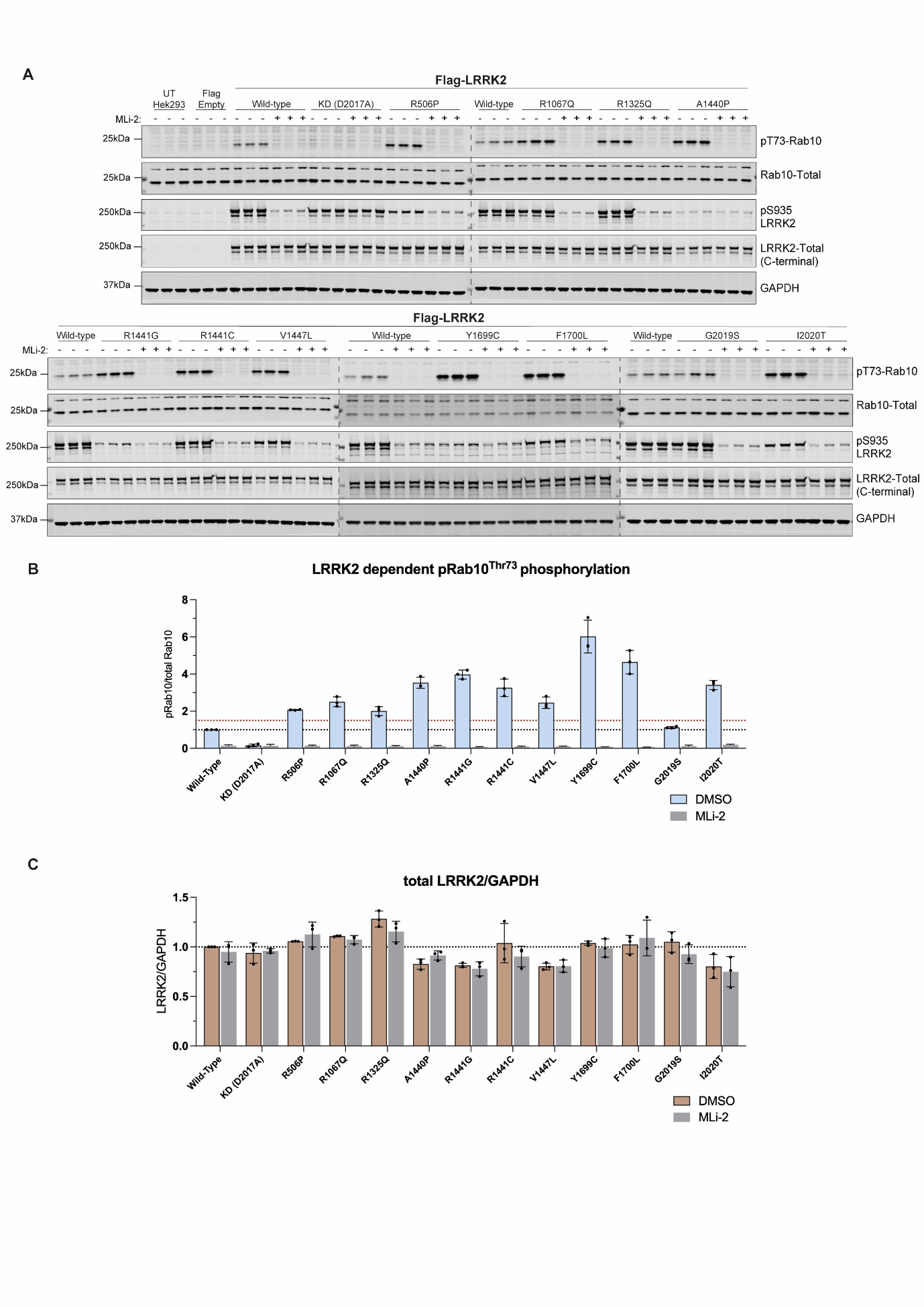

**(A)** FLAG-tagged LRRK2 wild-type (WT), kinase-dead (D2017A), and the indicated variants were transiently expressed in HEK293 cells. Approximately 20 h post-transfection, cells were treated with or without 200 nM MLi-2 for 90 min prior to lysis. Equal amounts of protein (20 μg) were analysed by immunoblotting using the indicated antibodies and imaged on a LI-COR Odyssey CLx system. Quantified results were normalized to WT LRRK2 (set to 1.0) and are presented as **(B)** pRab10/total Rab10 and **(C)** total LRRK2/GAPDH. The red dashed line at 1.5-fold pRab10/total Rab10 represents the threshold used to classify variants as kinase activating. Each data point represents an independent biological replicate.
